# New tuberculosis vaccines in India: Modelling the potential health and economic impacts of adolescent/adult vaccination with M72/AS01_E_ and BCG-revaccination

**DOI:** 10.1101/2023.02.24.23286406

**Authors:** Rebecca A Clark, Chathika K Weerasuriya, Allison Portnoy, Christinah Mukandavire, Matthew Quaife, Roel Bakker, Danny Scarponi, Rebecca C Harris, Kirankumar Rade, Sanjay Kumar Mattoo, Dheeraj Tumu, Nicolas A Menzies, Richard G White

## Abstract

**Background:** India had an estimated 2.9 million tuberculosis cases and 506 thousand deaths in 2021. Novel vaccines effective in adolescents and adults could reduce this burden. M72/AS01_E_ and BCG-revaccination have recently completed Phase IIb trials and estimates of their population-level impact are needed. We estimated the potential health and economic impact of M72/AS01_E_ and BCG-revaccination in India and investigated the impact of variation in vaccine characteristics and delivery strategies.

**Methods:** We developed an age-stratified compartmental tuberculosis transmission model for India calibrated to country-specific epidemiology. We projected baseline epidemiology to 2050 assuming no-new-vaccine introduction, and M72/AS01_E_ and BCG-revaccination scenarios over 2025–2050 exploring uncertainty in product characteristics (vaccine efficacy, mechanism of effect, infection status required for vaccine efficacy, duration of protection) and implementation (achieved vaccine coverage and ages targeted). We estimated reductions in tuberculosis cases and deaths by each scenario compared to no-new-vaccine introduction, as well as costs and cost-effectiveness from health-system and societal perspectives.

**Results:** M72/AS01_E_ scenarios were predicted to avert 40% more tuberculosis cases and deaths by 2050 compared to BCG-revaccination scenarios. Cost-effectiveness ratios for M72/AS01_E_ vaccines were around seven times higher than BCG-revaccination, but nearly all scenarios were cost-effective. The estimated average incremental cost was US$190 million for M72/AS01_E_ and US$23 million for BCG-revaccination per year. Sources of uncertainty included whether M72/AS01_E_ was efficacious in uninfected individuals at vaccination, and if BCG-revaccination could prevent disease.

**Conclusions:** M72/AS01_E_ and BCG-revaccination could be impactful and cost-effective in India. However, there is great uncertainty in impact, especially given unknowns surrounding mechanism of effect and infection status required for vaccine efficacy. Greater investment in vaccine development and delivery is needed to resolve these unknowns in vaccine product characteristics.

## Background

India has the largest global burden of tuberculosis. In 2021, there were an estimated 2.9 million cases and 506 thousand deaths–representing approximately 30% of the total globally.^1^ The COVID-19 pandemic has negatively impacted tuberculosis prevention and care in India, with increases in the number of deaths per year seen for the first time since 2007.^1,2^ Delays in diagnosis and treatment due to surveillance systems impacted by the pandemic (over 30% fewer notifications reported in 2021 than 2019) may lead to increases in the disease burden.^1,2^

Tuberculosis is a key focus for the Indian government. The National Strategic Plan to End Tuberculosis in India 2020–2025, developed by the National Tuberculosis Elimination Programme (NTEP), outlines ambitious goals for reducing *Mycobacterium tuberculosis* (*Mtb*) transmission, preventing tuberculosis disease, and addressing social determinants of health.^3^ Despite the COVID-19 pandemic, the NTEP has made progress toward these goals, including expanding molecular diagnostics, implementing tuberculosis-COVID bidirectional screening, and expanding policy on preventive therapy to include all household contacts of people diagnosed with pulmonary tuberculosis.^4^

The National Strategic Plan also calls for further development in tuberculosis vaccines, which has been a high priority for global organisations such as the World Health Organization (WHO). A recently completed WHO-commissioned study assessing the full value of tuberculosis vaccines made a strong case from the health and economic perspectives for continued investment,^5–9^ and previous work has demonstrated that novel vaccines or vaccination strategies will be needed to eliminate tuberculosis.^10,11^

Currently, sixteen candidates are in various phases throughout the vaccine pipeline, being trialled in a variety of ages and spanning prevention of disease, infection, and recurrence endpoints.^12^ A phase IIb trial of M72/AS01_E_ in adolescents and adults infected with *Mtb* demonstrated a prevention of disease efficacy of 49.7% (95% confidence interval: 2.1–74.2) after three years follow-up.^13^ However, M72/AS01_E_ would need a supportive Phase III trial for licensure, which is planned but likely to require years before results are available to inform policy.

Revaccination of uninfected adolescents with the Bacillus Calmette–Guérin (BCG) vaccine was assessed as a third parallel arm in a separate Phase IIb trial, and demonstrated an efficacy of 45.4% (6.4–68.1) against sustained infection,^14^ and an additional Phase IIb confirmation trial is underway to verify this finding, with results expected mid-2024.^15^ The original Chingleput BCG vaccination trial reported efficacy of 27% (−8–50) against disease in children and no efficacy in adults.^16^ A re-analysis of trial data restricted to participants with prior BCG vaccination and no tuberculosis disease at the time of vaccination showed a protective efficacy of 36% (11–54) against disease.^17^ As BCG is already-licensed, introducing BCG-revaccination may only require policy change, which could happen quickly.

India is arguably the most important country for global tuberculosis elimination, and policy makers require country-specific evidence of the anticipated health, cost, and budget impacts of specific vaccine candidates. As vaccines enter Phase III trials, it is important to predict how variation in vaccine profile and implementation will affect impact to maximise benefits and reduce delays between licensure and delivery. We estimated the potential health and economic impact of M72/AS01_E_ and BCG-revaccination in India and investigated the impact of variation in vaccine characteristics and delivery strategies.

## Methods

### Data

We obtained demographic data for India from the United Nations Population Division with estimates for single ages and years from 1900–2100.^18^ Tuberculosis disease and infection prevalence estimates were derived from the National TB Prevalence Survey in India 2019–2021.^19^ Incidence, notifications, and mortality estimates were obtained from WHO.^2^

### Structure

We adapted previous models and developed a compartmental dynamic model of tuberculosis in India.^5,11,20^ Our model was stratified by tuberculosis natural history and treatment, differences in access-to-care, vaccination, and age. We represented tuberculosis natural history by allowing for *Mtb* infection along a spectrum from uninfected to active clinical disease. We assumed a progressive loss of ability to reactivate following infection, with a monotonic decline in reactivation rates for subsequent latency compartments. Active disease was represented by both subclinical and clinical tuberculosis compartments to align with prevalence survey data.^19^ Anti-tuberculosis treatment was assumed to begin in 1960, and increase following a sigmoid curve to 2020. Due to the large contribution of private sector treatment in India, we incorporated differences in treatment mortality and completion probabilities between the public and private sector. Full model structure and parameters are in Supplementary Material sections 1–2.

### Calibration

The model was fit to 19 tuberculosis-related calibration targets: the incidence rate (all ages, children, and adults in 2000, 2020, and 2025), mortality rate (all ages in 2000, 2020, and 2025), notification rate (all ages, children, and adults in 2000 and 2020), disease prevalence (all ages, children, and adults in 2015 and 2021), infection prevalence (all ages in 2021), the proportion of incident cases with treatment history in 2020, the fraction of subclinical tuberculosis among active tuberculosis in 2020, and the prevalence ratio of active tuberculosis between access-to-care compartments in 2020 all assuming a uniform distribution between lower and upper bounds. We calibrated using the *hmer* R package^21^ to perform history matching with emulation followed by ABC-MCMC until we obtained 1000 parameter sets fitting all targets (further information in Supplementary Material section 3).

### Scenarios

#### i. No-new-vaccine baselines

Assuming the quality and coverage of services remains constant post 2020, we used the calibrated model to project baseline epidemiology to 2050 (the *Status Quo* no-new-vaccine baseline). We assumed that neonatal BCG vaccination would not be discontinued during the period of our analysis and was not explicitly modelled as its effect is implicitly included in country burden estimates.

As an alternative future scenario, we calibrated a *Strengthened Current Interventions* no-new-vaccine baseline. This baseline assumed scale-up of non-vaccine tuberculosis interventions between 2021 and 2035 to meet the target of a 50% reduction in tuberculosis incidence in 2035 compared to the 2015 estimates. This scale-up was included in the model by introducing multipliers on the rate of progression to disease and in the force of infection equation.

#### ii. Vaccine scenarios

Using the calibrated *Status Quo* no-new-vaccine model, we simulated Basecase scenarios over 2025–2050 for each product with characteristics informed a priori by clinical trial data and expert opinion.^13,14^ The Basecase M72/AS01_E_ scenario assumed a 50% efficacy prevention of disease vaccine with 10-years protection, efficacious with any infection status aside from active disease at vaccination. We assumed the vaccine would be introduced in 2030 routinely to those aged 15 (reaching 80% coverage) and as a campaign for ages 16–34 (reaching 70% coverage), with a repeat campaign in 2040. Based on expert advice, the vaccine price was $2.50 per dose, assuming two doses per course.

The Basecase BCG-revaccination scenario assumed a 45% efficacy vaccine to prevent infection with 10-years protection, and efficacious without infection at time of vaccination. We assumed the vaccine would be introduced in 2025 routinely to those aged 10 (reaching 80% coverage) and as a campaign for ages 11–18 (reaching 80% coverage) with repeat campaigns in 2035 and 2045. Based on the average estimated BCG price from UNICEF,^22^ the vaccine price was set at US$0.17 per dose, assuming one dose per course.

Vaccine introduction costs for both vaccine products were assumed to be US$2.40 (95% uncertainty interval = 1.20–4.80) per individual in the targeted age group based on vaccine introduction support policy from Gavi, the Vaccine Alliance.^23^ A further US$0.11 (0.06–0.22) supply costs and US$2.50 (1.00–5.00) delivery costs per dose were included,^24^ as well as US$0.94 (0.13–1.52) in patient and caregiver productivity losses per dose, to account for the time taken to receive vaccination.^25,26^ We assumed a 5% wastage rate.

Through consultation with vaccine and country-specific experts, we established specific M72/AS01_E_ and BCG-revaccination *Policy Scenarios* and *Vaccine Characteristic and Coverage Scenarios*. *Policy Scenarios* represented features of vaccination strategy under the control of decision-makers, which compared different age groups to target for vaccination. *Vaccine Characteristic and Coverage Scenarios* represented current uncertainties around vaccine performance and uptake, in which we varied unknowns in vaccine profile (such as efficacy, duration of protection, mechanism of effect) and achieved coverage, univariately from each Basecase scenario. We compared *Policy Scenarios* to identify the optimal implementation approach, and *Vaccine Characteristic and Coverage Scenarios* to quantify the impact of different sources of uncertainty. (Table 1).

**Table 1.**
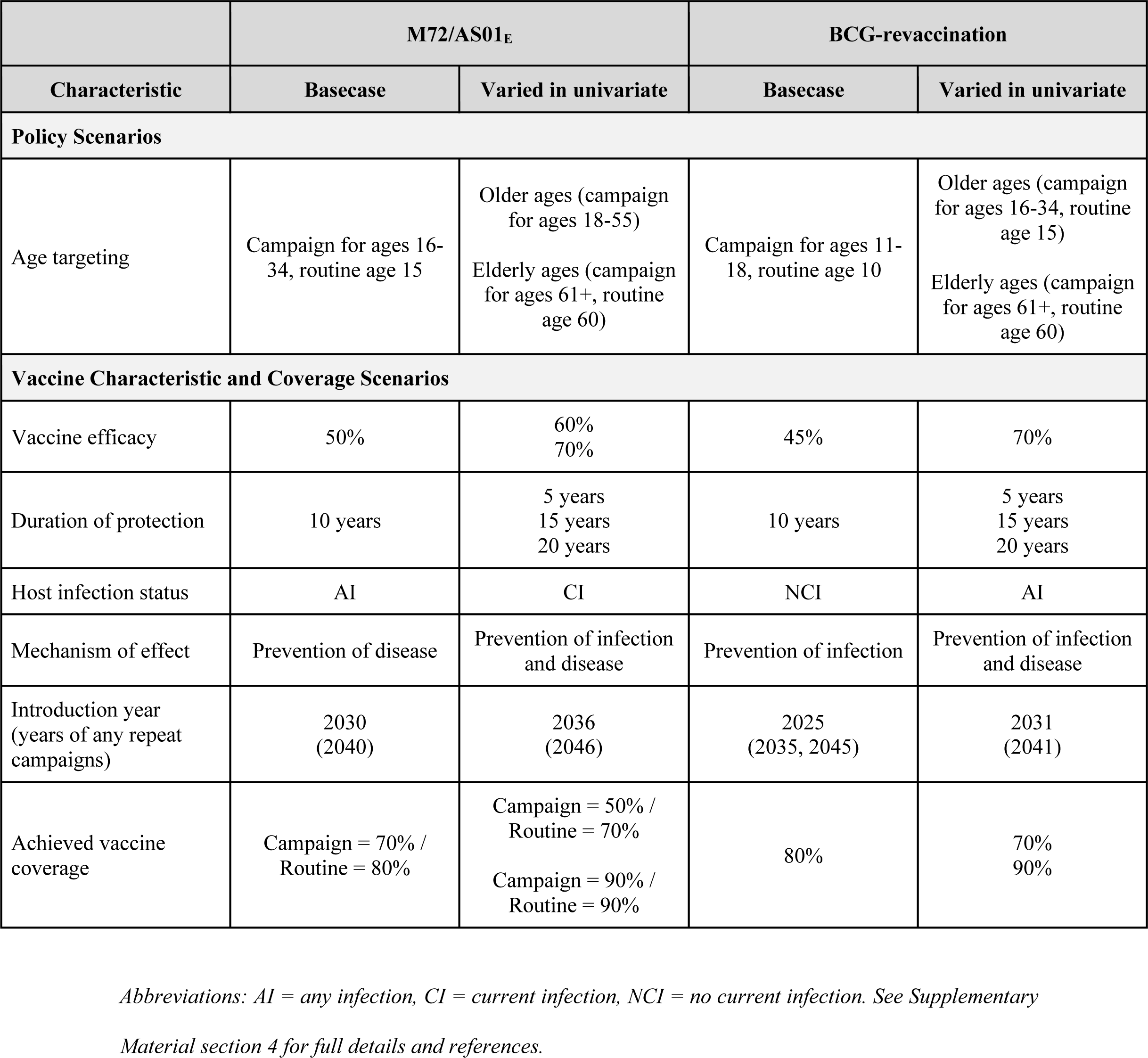
Assumed M72/AS01_E_ and BCG-revaccination scenarios

|  | M72/AS01<sub>E</sub> |  | BCG-revaccination |  |
| --- | --- | --- | --- | --- |
| Characteristic | Basecase | Varied in univariate | Basecase | Varied in univariate |
| **Policy Scenarios** |  |  |  |  |
| Age targeting | Campaign for ages 16-34, routine age 15 | Older ages (campaign for ages 18-55)<br>Elderly ages (campaign for ages 61+, routine age 60) | Campaign for ages 11-18, routine age 10 | Older ages (campaign for ages 16-34, routine age 15)<br>Elderly ages (campaign for ages 61+, routine age 60) |
| **Vaccine Characteristic and Coverage Scenarios** |  |  |  |  |
| Vaccine efficacy | 50% | 60%<br>70% | 45% | 70% |
| Duration of protection | 10 years | 5 years<br>15 years<br>20 years | 10 years | 5 years<br>15 years<br>20 years |
| Host infection status | AI | CI | NCI | AI |
| Mechanism of effect | Prevention of disease | Prevention of infection and disease | Prevention of infection | Prevention of infection and disease |
| Introduction year (years of any repeat campaigns) | 2030<br>(2040) | 2036<br>(2046) | 2025<br>(2035, 2045) | 2031<br>(2041) |
| Achieved vaccine coverage | Campaign = 70% /<br>Routine = 80% | Campaign = 50% /<br>Routine = 70%<br>Campaign = 90% /<br>Routine = 90% | 80% | 70%<br>90% |

*Abbreviations: AI = any infection, CI = current infection, NCI = no current infection. See Supplementary*
*Material section 4 for full details and references.*

### Outcomes

We estimated the cumulative number of tuberculosis cases and deaths averted between vaccine introduction and 2050 for each scenario compared to the predicted numbers in both no-new-vaccine baselines.

For each vaccine product, we conducted cost-effectiveness analyses for the *Policy Scenarios* indicated in Table 1, discounting both costs and health outcomes to 2025 (when vaccination began) at 3% per year as per guidelines.^27^ We calculated the difference in total disability-adjusted life years (DALYs) from vaccine introduction to 2050, using the disability weight for tuberculosis disease from the Global Burden of Disease 2019 study,^28^ and country- and age-specific life expectancy estimates from the United Nations Development Programme assuming no post-tuberculosis morbidity or mortality.^29^ We calculated incremental cost-effectiveness ratios (ICERs) as the ratio of mean incremental costs to mean incremental benefits in DALYs averted, and 95% uncertainty intervals from the health-system perspective for each efficient strategy for the analytic period 2025–2050. Higher cost-effectiveness ratios indicate greater spending is needed to achieve health improvements, such that the intervention is less likely to be cost-effective. We measured cost-effectiveness by 2050 against three India-specific cost thresholds: 1x gross domestic product (GDP) per capita (US$1,927.71),^26^ and country-level opportunity cost thresholds defined by Ochalek et al (country-level upper [US$363] and lower [US$264] bounds).^30^

To investigate how the consequences of vaccine introduction (versus no vaccination) changed based on the vaccine product characteristics, we examined the difference in ICERs for *Vaccine Characteristic and Coverage Scenarios* compared to the no-new-vaccine baseline assuming the vaccine was introduced using the delivery strategy from the most efficient *Policy Scenario* at the country-level lower bound.

We estimated the annual incremental costs of diagnosis, treatment, and vaccination for each scenario, as compared to the no-new-vaccine baseline in 2020 US dollars from health-system and societal perspectives.

## Results

The *Status Quo* baseline model fit all 19 calibration targets with at least 1000 parameter sets. Epidemiological projections from 2020–2050 are in Supplementary Material section 7. The *Status Quo* baseline predicted 72.2 (63.3–79.7) million incident tuberculosis cases and 13.8 (12.9–15.2) million tuberculosis deaths between 2025–2050. Assuming current non-vaccine tuberculosis interventions would be strengthened such that the incidence rate in 2035 was 50% of the incidence rate in 2015, the *Strengthened Current Interventions* baseline predicted 36.0 (28.9–66.4) million incident cases and 7.6 (6.1–13.2) million deaths between 2025–2050.

With the *Status Quo* no-new-vaccine baseline, we found a 50% efficacy M72/AS01_E_ prevention of disease vaccine, efficacious with any infection status, introduced in 2030 routinely to 15-year-olds and as a campaign for ages 16–34, could avert approximately 12.7 (11.0–14.6) million cases and 2.0 (1.8–2.4) million deaths between 2030–2050 (Figure 1). With a 70% efficacy vaccine, the number of averted cases and deaths by 2050 could be increased by 32–35% but delaying introduction of a vaccine until 2036 could lead to 5.2 million more cases and 968 thousand more deaths compared to the Basecase before 2050 (Figure 1). If the vaccine was only efficacious with current infection at vaccination, 5.8 million fewer cases and 900 thousand fewer deaths could be averted compared to the Basecase.

**Figure 1.**
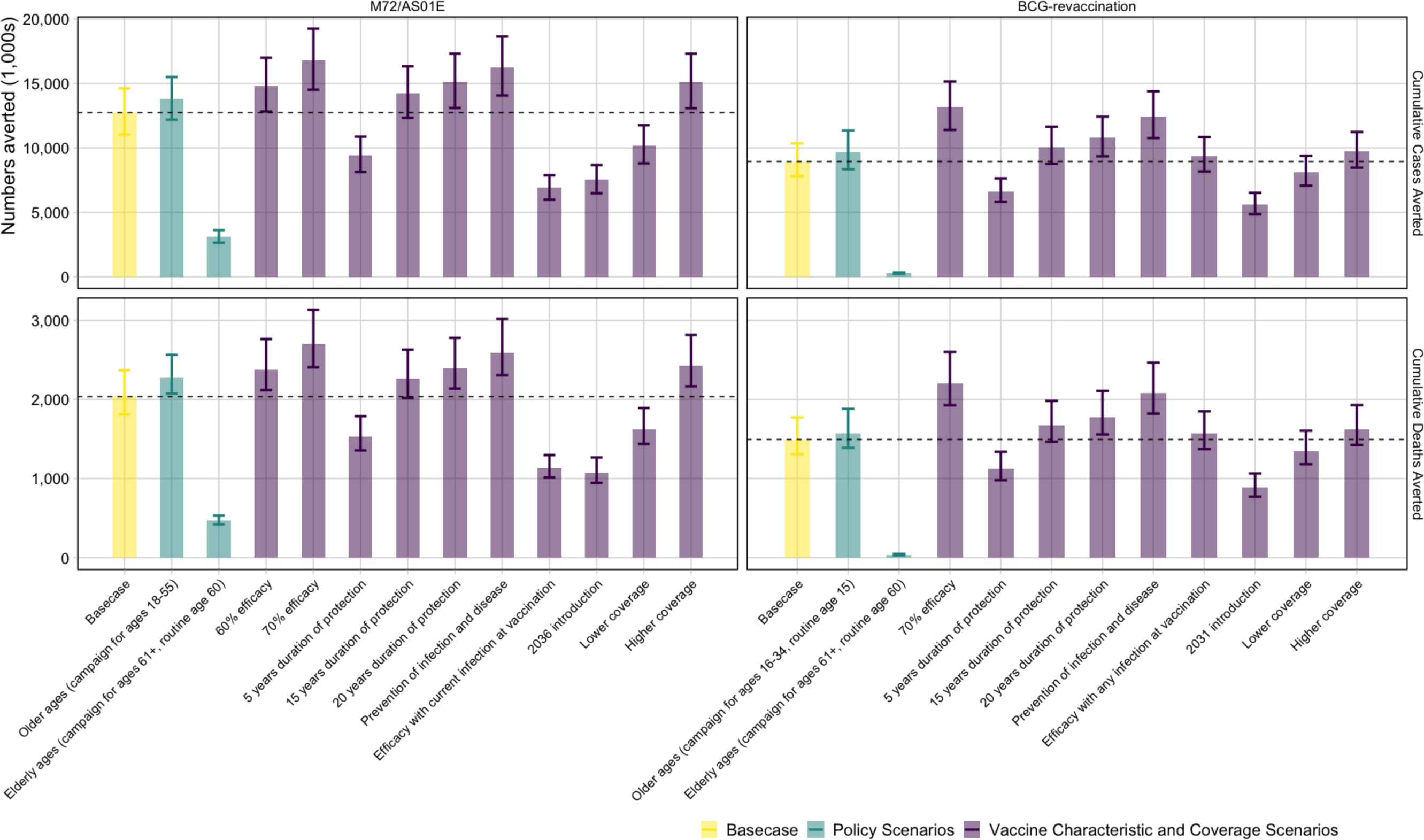
Cumulative cases and deaths averted (in 1,000s) by 2050 from M72/AS01_E_ and BCG-revaccination scenarios. *The top of the bar is the median estimate of the number averted for each scenario compared to the estimated number predicted by 2050 with the no-new-vaccine baseline with 95% uncertainty range. The horizontal line is the median value of the Basecase for each vaccine. The cases and deaths averted by each scenario are compared to 72.2 (63.3–79.7) million incident tuberculosis cases and 13.8 (12.9–15.2) million tuberculosis deaths predicted by the Status Quo baseline between 2025–2050*.

A 45% efficacy prevention of infection BCG vaccine, efficacious in those with no current infection, introduced in 2025 as routine vaccination of 10-year-olds and a campaign for ages 11– 18 could avert 9.0 (7.8–10.4) million cases and 1.5 (1.3–1.8) million deaths (Figure 1). If the vaccine prevented infection and disease, 3.4 million more cases and 600 thousand more deaths could be averted by 2050 compared to the Basecase. Fewer numbers could be averted compared to the Basecase with reduced duration of protection, later introduction, lower coverage, or only delivering the vaccine to ages 60 years and older (Figure 1).

Comparing the two products, even with a later introduction year for M72/AS01_E_, we found a higher health impact from M72/AS01_E_ vaccines compared to BCG-revaccination. The Basecase M72/AS01_E_ scenario was predicted to avert around 40% more tuberculosis cases and deaths before 2050 than the Basecase BCG-revaccination scenario.

With the *Strengthened Current Interventions* baseline, the Basecase M72/AS01_E_ scenario could avert 3.0 (1.1–11.3) million tuberculosis cases and 0.51 (0.19-1.9) million tuberculosis deaths between 2025–2050, averting 8.3% of the median total cases and 6.7% of the median total deaths predicted to occur during the same period. The Basecase BCG-revaccination scenario could avert 1.9 (0.42–8.0) million cases and 0.34 (0.08-1.4) million deaths between 2025–2050, or 5.3% of the median total tuberculosis cases and 4.5% of the median total tuberculosis deaths predicted to occur during the same period. Health impact values for all scenarios of both vaccines are in Supplementary Material section 8.

Cost-effectiveness analysis is shown in Table 2 and Figure 2 for the *Policy Scenarios* for each vaccine product. For M72/AS01_E_, delivering the vaccine routinely to those age 60 and as a campaign for ages 61+ (Elderly Ages) was not efficient and removed from consideration. Scenarios delivering the vaccine routinely to age 15 and as a campaign for ages 16–34 (Basecase) and delivering the vaccine as a campaign for ages 18–55 (Older Ages) were considered efficient and displayed on the efficiency frontier in Figure 2. The Basecase scenario was optimal at both country-level thresholds (ICER = US$145 per DALY averted), and the Older Ages scenario was optimal at 1x GDP threshold (ICER = US$1,120 per DALY averted). The incremental cost of the Basecase M72/AS01_E_ scenario was US$5.3 billion, with vaccination averting 36.9 million of the 4.0 billion DALYs predicted by the no-new-vaccine baseline between 2025–2050.

**Figure 2.**
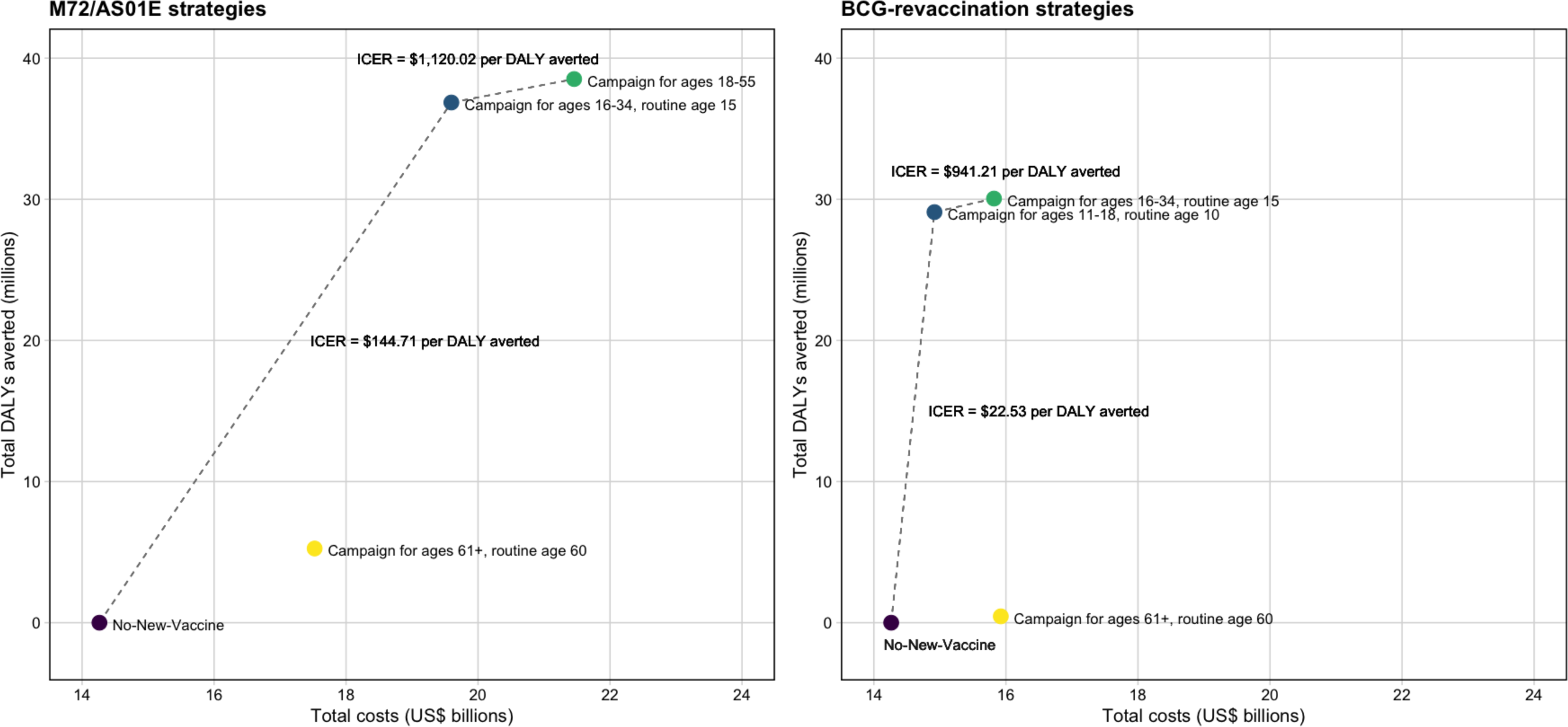
Efficiency frontiers (discounted total costs [US$ billions] per disability-adjusted life year (DALY) averted) for *Policy Scenarios* for each vaccine product

**Table 2.**
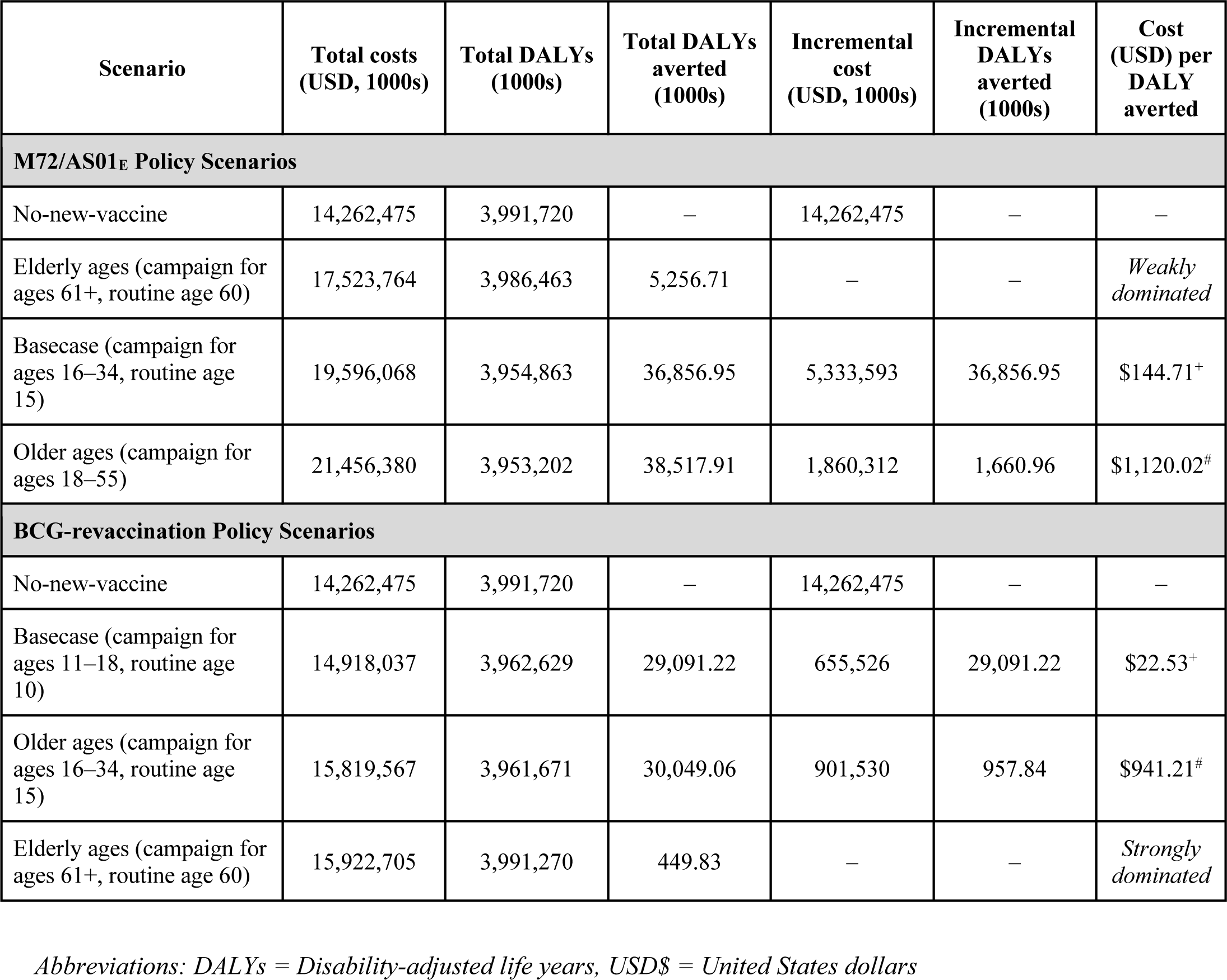
Cost-effectiveness analysis for M72/AS01_E_ and BCG-revaccination *Policy Scenarios*

For BCG-revaccination, delivering the vaccine routinely to those age 60 and as a campaign for ages 61+ (Elderly Ages) was dominated by other strategies and removed from consideration. Scenarios delivering the vaccine routinely to age 10 and as a campaign for ages 11–18 (Basecase) and delivering the vaccine routinely to those aged 15 and as a campaign for ages 16–34 (Older Ages) were considered efficient and displayed on the efficiency frontier in Figure 2. The Basecase scenario (ICER = US$23 per DALY averted) was optimal at both country-level thresholds and the Older Ages scenario (ICER = US$941 per DALY averted) was optimal at 1xGDP threshold. The incremental cost of the Basecase BCG-revaccination scenario was US$656 million, and this strategy averted 29.1 million of the 4.0 billion DALYs predicted by the no-new-vaccine baseline between 2025–2050.

Figure 3 displays the ICERs for each *Vaccine Characteristic and Coverage Scenario* compared to the no-new-vaccine baseline for each vaccine product. For every M72/AS01_E_ scenario shown in the figure, we assumed that the vaccine would be introduced routinely to those aged 15 and as a campaign to ages 16–34 (the most efficient strategy at the country-level lower bound from the cost-effectiveness analysis). Even with changes in the vaccine product characteristics, introducing an M72/AS01_E_ vaccine would be cost-effective compared to not implementing a vaccine (Figure 3). For every BCG-revaccination scenario, we assumed that the vaccine would be introduced routinely to those aged 10 and as a campaign to ages 11–18 (the most efficient strategy at the country-level lower bound from the cost-effectiveness analysis). Similarly, regardless of the resulting product characteristics, introducing BCG-revaccination to this age group would be cost-effective compared to not implementing a vaccine (Figure 3).

**Figure 3.**
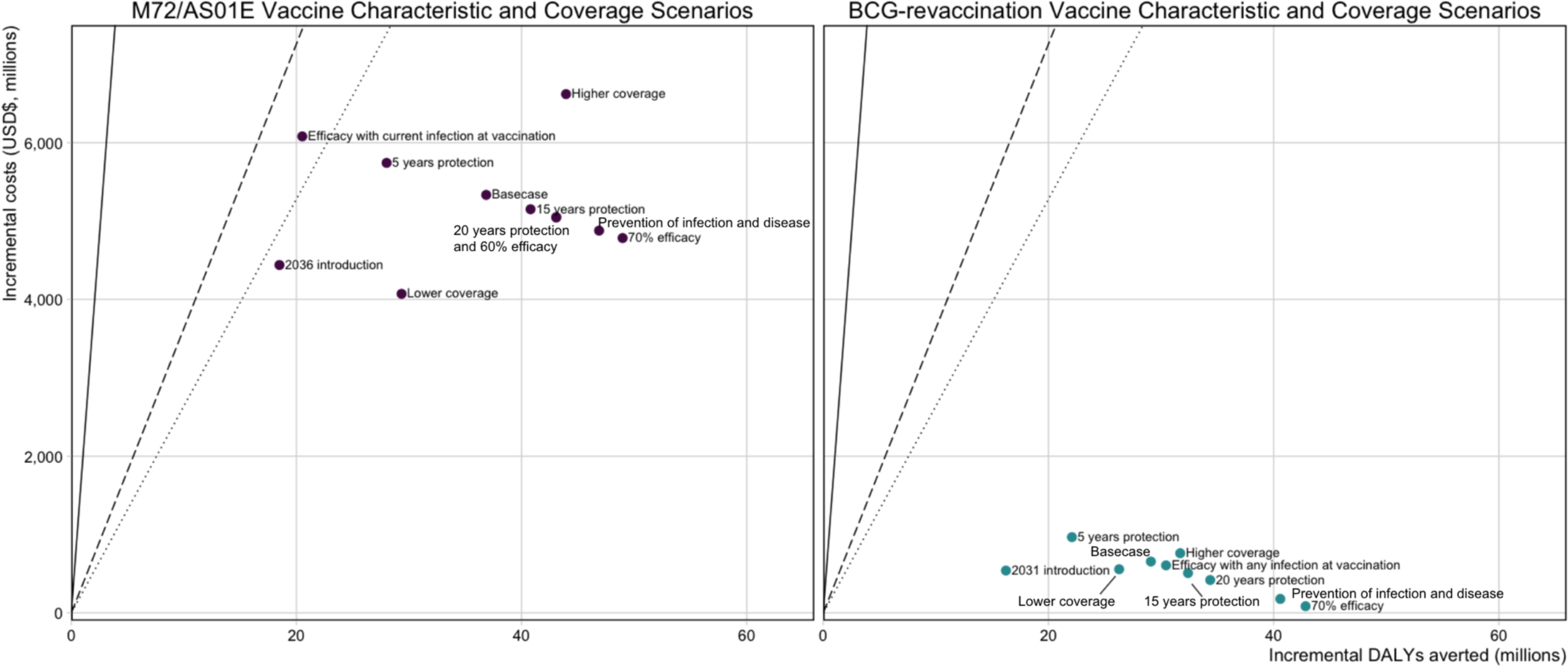
Comparison of ICERs for *Vaccine Characteristic and Coverage Scenarios* compared to the no-new-vaccine baseline for each vaccine product. The Basecase M72/AS01_E_ scenario assumes a 50% efficacy POD vaccine efficacious with any infection status at the time of vaccination, with 10 years duration of protection reaching 80% coverage for 15-year-olds and 70% coverage for those aged 16–34. Each M72/AS01_E_ scenario is delivered routinely to those aged 15 and as a campaign for those aged 16–34. The Basecase BCG-revaccination scenario assumes a 45% efficacy POI vaccine efficacious with no current infection at the time of vaccination, with 10 years duration of protection and reaching 80% coverage. Each BCG-revaccination scenario is delivered routinely to those aged 10 and as a campaign for those aged 11–18. The scenarios on the figure are labelled with the difference in product characteristics for that scenario compared to the Basecase. The 20 years protection and 60% efficacy scenarios for M72/AS01_E_ overlap and appear as one point on the figure.

From the health-system perspective, the annual average cost of vaccination in the Basecase M72/AS01_E_ scenario was approximately US$251 (170–368) million between 2025–2050. The annual average cost-savings in treatment and diagnostics were US$60 (49–74) million over 2025– 2050. The annual average cost of vaccination in the Basecase BCG-revaccination scenario was US$67 (29–122) million over 2025–2050. The annual average cost-savings in treatment and diagnostics were US$43 (35–55) million over 2025–2050. The average annual cost of vaccination in the Basecase M72/AS01_E_ scenario was almost four times greater than the average annual cost of vaccination with the Basecase BCG-revaccination scenario. Accounting for cost-savings, the average annual incremental program cost in the Basecase M72/AS01_E_ scenario (US$190 million) was over eight times greater than the average annual incremental program cost with the Basecase BCG-revaccination scenario (US$23 million).

Figure 4 demonstrates the distribution of costs and cost-savings per year from vaccine introduction to 2050 for the Basecase scenarios for both vaccine products. During the initial 5-year scale-up to maximum achieved coverage, the average vaccination cost for the Basecase M72/AS01_E_ scenario was US$638 million per year, compared to US$121 million per year for the Basecase BCG-revaccination scenario. The cost during the repeat campaign in 2040 for the Basecase M72/AS01_E_ vaccine was US$2.2 billion, compared to US$377 million and US$272 million, respectively, for the two repeat campaigns in 2035 and 2045 for the Basecase BCG-revaccination scenario. Full economic results are in Supplementary Material section 9.

**Figure 4.**
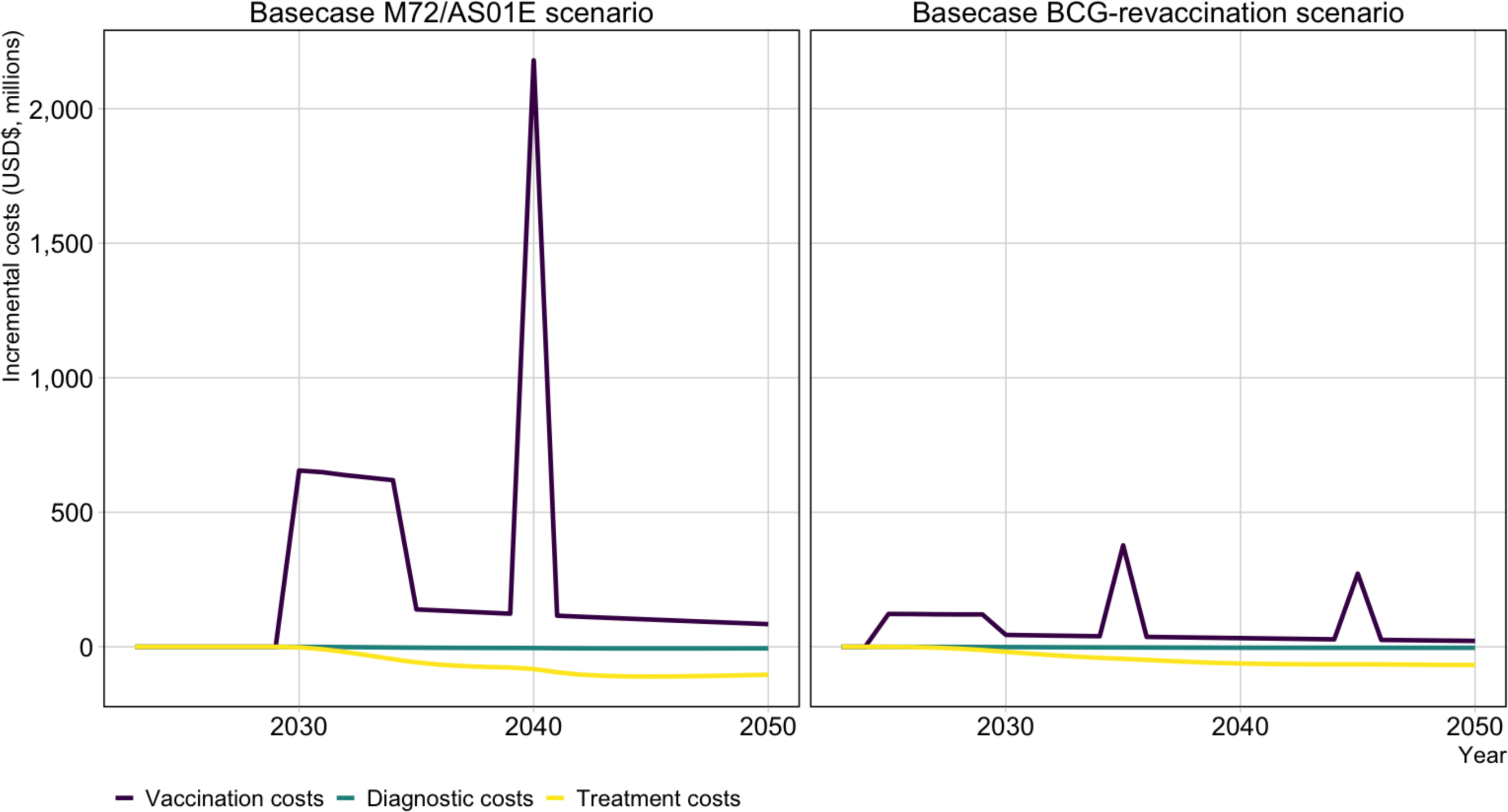
Incremental costs by year until 2050 for the Basecase M72/AS01_E_ and BCG-revaccination scenarios compared to the no-new-vaccine baseline. *Abbreviations: USD$ = United States dollars*.

## Discussion

We found that M72/AS01_E_ scenarios could avert approximately 12.7 (11.0–14.6) million cases and 2.0 (1.8–2.4) million deaths, and BCG-revaccination scenarios could avert approximately 9.0 (7.8–10.4) million cases and 1.5 (1.3–1.8) million deaths of the 72.2 (63.3–79.7) million cases and 13.8 (6.1–13.2) million deaths predicted by the *Status Quo* baseline between 2025 and 2050. Cost-effectiveness ratios for the Basecase M72/AS01_E_ scenario was around seven times higher than that for the Basecase BCG-revaccination scenario, but regardless of the realised product characteristics, nearly all *Vaccine Characteristic and Coverage Scenarios* were cost-effective at the most conservative country-level threshold compared to the no-new-vaccine baseline. The average annual cost of M72/AS01_E_ vaccination was four times greater than BCG-revaccination. Introducing the vaccine could lead to an annual incremental program cost of US$190 million for M72/AS01_E_ and US$23 million for BCG-revaccination, accounting for vaccination costs as well as savings in diagnostic and treatment costs.

Our modelling demonstrated a 40% greater health impact from M72/AS01_E_ compared to BCG-revaccination. The difference in impact was due to assumptions made on vaccine characteristics and delivery. Based on clinical trial data and expert opinion, we assumed the Basecase M72/AS01_E_ vaccine would prevent disease and be efficacious in everyone without active disease at vaccination. In contrast, based on trial data,^14,31^ we assumed the Basecase BCG-revaccination scenario would be efficacious only in people without infection at the time of vaccination, and would prevent infection. Therefore, M72/AS01_E_ would be effective in a larger proportion of the population compared to BCG-revaccination and have a more rapid impact on tuberculosis incidence. The effect of BCG-revaccination on disease will be delayed by the time between vaccination and infection in addition to the time from infection to disease. This is consistent with previous work showing more rapid impact on disease of a vaccine that prevents disease directly in those currently infected.^11^

As demonstrated in the National Tuberculosis Prevalence Survey, the highest tuberculosis prevalence estimates are found in older adolescents and adults.^19^ The Basecase scenario for M72/AS01_E_ delivered the vaccine routinely to those aged 15 and as a campaign for ages 16–34, as opposed to BCG-revaccination which was targeted routinely to those aged 10 and a campaign for ages 11–18. As the M72/AS01_E_ vaccine was targeted to an age group with a higher burden, we saw increased impact on burden.

We explored variation in decisions regarding delivery and the realised vaccine by evaluating *Policy Scenarios* and *Vaccine Characteristic and Coverage Scenarios* where we varied characteristics univariately from each Basecase, and found all uncertainties had the anticipated direction of effect. Both M72/AS01_E_ and BCG-revaccination were highly influenced by vaccine efficacy and duration of protection, with higher efficacies and longer durations of protection increasing health impact and cost-effectiveness. Key sources of uncertainty were whether M72/AS01_E_ was efficacious without infection at vaccination, and if BCG-revaccination was also able to prevent disease in adults, both of which are key areas of research. Given the uncertainty surrounding prevention of disease efficacy from BCG-revaccination, any roll out of BCG to adolescents and adults should be rigorously evaluated with a prevention of disease outcome.

M72/AS01_E_ scenarios were predicted to have higher vaccination costs per year compared to BCG-revaccination. The assumed M72/AS01_E_ vaccine price per course of US$5.00 (two doses for US$2.50 each) was almost 30 times the US$0.17 price per course of BCG-revaccination, in addition to duplicated delivery and supply costs necessary to deliver two doses of M72/AS01_E_ compared to one dose of BCG. These cost differences directly contribute to higher cost-effectiveness ratios and larger annual cost for M72/AS01_E_. Our analyses demonstrated that both vaccines could be cost-effective, aligning with previous cost-effectiveness analyses of tuberculosis analyses demonstrated that both vaccines could be vaccines.6,32 While vaccination could have a substantial budget impact, costs could be partially offset with diagnostic and treatment savings.

Comparing the ICERs for *Vaccine Characteristic and Coverage Scenarios*, we see that even if the product characteristics change from the Basecase scenario for each vaccine product, the decision remains the same. Introducing M72/AS01_E_ or BCG-revaccination would be a cost-effective intervention.

This work has limitations. We modelled the impact of specific M72/AS01_E_ and BCG-revaccination scenarios with characteristics based on clinical trial data and consultation with vaccine and country-specific experts, but it will be many years before the actual characteristics are known. To capture some uncertainty, we univariately varied efficacy, duration of protection, whether the vaccine prevents only infection or disease or both, and who the vaccine would be efficacious in. The majority of scenarios continued to demonstrate large potential health impact and cost-effectiveness. We were not investigating the separate question of determining the range of plausible conditions that M72/AS01_E_ would no longer be cost-effective or scenarios where BCG-revaccination would have a greater impact, which is an important area for future work to address.

The Basecase M72/AS01_E_ scenario assumed efficacy with any infection status at vaccination, implying that the vaccine would work in both those who were infected with *Mtb* and those who were uninfected. While the Phase IIb trial of M72/AS01_E_ only enrolled adults with a positive interferon-gamma release assay (IGRA) value, previous trials have indicated that an immune response is invoked in adolescents both with and without infection, and the Phase III trial will enrol IGRA positive and negative individuals aged 15–44 years. Therefore, the expected initial indicated population is everyone within these ages, and thus we aligned our primary assumption for host infection status with this. We evaluated a scenario assuming only current infection at vaccination and determined that efficacy in those who are uninfected at the time of vaccination is important to maximise health impact and cost-effectiveness. Investigating whether M72/AS01_E_ works in populations with any infection status is a key aspect for future research.

We modelled a small subset of age-targeted delivery scenarios, which may differ from the strategies India will choose. We evaluated alternatives informed by expert opinion and results from interviews with key decision-makers in India,^33^ but did not investigate targeting specific groups, such as healthcare workers, people completing tuberculosis treatment, or household contacts of people with tuberculosis, who could be at high risk of developing tuberculosis disease and may be prioritised for vaccination. This strategy has previously been suggested to have a high population-level impact per individual vaccinated,^34,35^ and is an important aspect for future country-level models to address to support decision making. To inform policy, we only modelled scenarios with vaccine efficacy greater than 50% for M72/AS01_E_ and greater than 45% for BCG-revaccination (aligning with the median estimates of protection from the Phase IIb trials). However, the true vaccine efficacy is currently unknown, and if our assumptions were too optimistic, we may have overestimated the health and economic impacts.

The burden of tuberculosis varies widely across India. From the recent National Tuberculosis Prevalence Survey, the prevalence per 100,000 population of pulmonary tuberculosis among adults ranged from 115 (47–184) in Kerala to 534 (365–704) in Delhi.^19^ Optimal delivery strategies may vary by state, given the vast differences in age composition, population size, and tuberculosis burden. Modelling specific regions to investigate the generalisability of national predictions is an important area of future research.

We ran cost-effectiveness analysis for each product on the mutually exclusive age-targeting *Policy Scenarios*. We selected the Basecase vaccine profile characteristics for each vaccine product as it incorporates the primary assumptions from experts in the field on the likely vaccine product characteristics, but we did not run cost-effective analysis for the age-targeting strategies with other vaccine characteristics.

Our work is a modelling exercise, and limitations associated with mathematical models apply. We developed our tuberculosis natural history structure incorporating recent advances in knowledge regarding the clinical course of disease, such as subclinical tuberculosis and a latency structure with a progressive loss in the ability to reactivate. If our assumptions around these novel aspects, particularly around interactions with vaccines, are incorrect, we may have over or underestimated the impact. While we used the best available data to inform calibration targets and natural history parameters, we were limited by what was available. We ensured that the modelled trends aligned with the most recent estimates of tuberculosis burden, as vaccines are not anticipated to be introduced until at least 2025. However, with only one estimate of whole-country disease prevalence and one estimate of whole-county infection prevalence in India, we were restricted with what we could infer about these measures over time, which highlights the need for more regularly collected data on disease prevalence and infection. We made decisions on natural history parameter ranges based on the most recent literature available, but this still resulted in wide prior ranges for some parameters. Further data collection into these areas would improve model estimates.

We projected the no-new-vaccine baseline as *Status Quo*, where we assume that the rate and quality of services remained constant from 2020 onwards, and the resulting trends in burden from 2020–2050 follows a slight decline. Given the commitment of the Indian government to improvements in tuberculosis care, prevention, and ending the tuberculosis epidemic, our model could be overestimating the burden of tuberculosis. Therefore, our health and economic impacts may be overestimated. We ran a sensitivity analysis for the Basecase scenario for each vaccine product using the *Strengthened Current Interventions* no-new-vaccine baseline. We found that vaccines would still have a positive health impact and would be cost-effective even if the incidence rate was declining faster than assumed in our primary scenario. We demonstrated that vaccines could also be an impactful and cost-effective investment for the Indian government if future tuberculosis burden is much lower.

The results from this study could be used to inform policy makers considering novel tuberculosis vaccine introduction. We have demonstrated that both BCG-revaccination and M72/AS01_E_ could have a positive health impact and would be cost-effective if delivered, given our current assumptions. We evaluated uncertainty surrounding vaccine characteristics, and found that even if characteristics were changed, we would still see positive health impact and cost-effectiveness.

The decision for how to take these results forward to country-level introduction lies with the policy maker, and how they are able to allocate their available budget. While we made some comparisons between products, the results of our study assume a reality where only one vaccine product is introduced. However, it is likely that both vaccine products could be introduced into the population, and the resulting health benefit could be increased. BCG is already licenced and recommended by the WHO for infants, and therefore BCG-revaccination of older adolescents and adults could be introduced earlier than M72/AS01_E_ through a policy change. Resources may need to be spent on epidemiological studies investigating population characteristics, such as the infection prevalence, to determine where a vaccine effective in those who are uninfected will have the most impact. M72/AS01_E_ is still a vaccine candidate and forward progression depends on results from the Phase III trial which has yet to start. More uncertainty surrounding costs and product characteristics exist, but overall M72/AS01_E_ predicted an increased health impact compared to BCG-revaccination.

## Conclusions

We propose it is inadvisable to focus solely on one or two vaccine candidates to address the tuberculosis burden. While promising results have been seen from recent trials, it will be years before we can verify these characteristics, and therefore we need a wide selection of options for the greatest likelihood of mitigating tuberculosis burden. We need to continue investment in all candidates currently in the pipeline, and support the development of new candidates, to increase the probability of success.

Our modelling suggests that M72/AS01_E_ and BCG-revaccination may substantially reduce the tuberculosis burden in India over future decades and would be cost-effective regardless of the realised product characteristics. We informed vaccine characteristics using clinical trial data but found variability in the vaccine profile as a crucial source of uncertainty. We cannot solely rely on M72/AS01_E_ and BCG-revaccination in case the realised characteristics differ considerably from expectations. Investment in multiple vaccine developments and delivery should be increased to raise the probability of success.

## List of Abbreviations

BCG: Bacillus Calmette–Guérin
DALY: Disability-adjusted life years
ICER: Incremental cost-effectiveness ratio
*Mtb*: *Mycobacterium tuberculosis*
NTEP: National Tuberculosis Elimination Programme
WHO: World Health Organization

## Data Availability

Epidemiologic data used are available from the World Health Organization Global TB Report CSV files to download (https://www.who.int/teams/global-tuberculosis-programme/data) and summarised in the Supplementary Material. Population estimates and projections are available from the United Nations Department of Economic and Social Affairs World Population Prospects 2019 (https://population.un.org/wpp/Download/Standard/Population/). Analytic code will be made available at https://doi.org/10.5281/zenodo.6421372 immediately following publication indefinitely for anyone who wishes to access the data for any purpose.

## Declarations

### Ethics approval and consent to participate

Not applicable

### Consent for publication

Not applicable

### Availability of data and materials

Epidemiologic data used are available from the *World Health Organization Global TB Report CSV files to download* (https://www.who.int/teams/global-tuberculosis-programme/data) and summarised in the Supplementary Material. Population estimates and projections are available from the *United Nations Department of Economic and Social Affairs World Population Prospects 2019* (https://population.un.org/wpp/Download/Standard/Population/). Analytic code will be made available at https://doi.org/10.5281/zenodo.6421372 immediately following publication indefinitely for anyone who wishes to access the data for any purpose.

### Authors’ contributions

Conception: RCH, RGW, NAM, CKW

Data acquisition and preparation: RAC, AP, CM, MQ, CKW Data analysis: RAC, RGW, CM, CKW, RB

Interpretation of results: RAC, RGW, CKW, AP, NAM, CM

Manuscript drafting and revisions: RAC, RGW, AP, NAM, CKW, MQ, DT, KR, CM, RB, SKM, RCH, DS

All authors had the opportunity to access and verify the data and were responsible for the decision to submit the manuscript for publication.

## Acknowledgements

We thank the Bill & Melinda Gates Foundation for providing funding (INV-001754) to undertake this research.

