## Supplementary Material for "New tuberculosis vaccines in India: Modelling the potential health and economic impacts of adolescent/adult vaccination with M72/AS01_E_ and BCG-revaccination"

Rebecca A. Clark et. al

**Table of Contents**


**SUPPLEMENTARY METHODS** **3**

**1. Model structure and equations** **3**

1.1 Natural history model structure 3

1.2 Natural history model equations 4

**2. Natural history** **5**

2.1 Natural history parameter values and data sources 5

2.2 Operationalising age varying parameters 7

2.3 Treatment initiation and outcomes 8

**3. Model simulation and calibration** **9**

3.1 Model simulation 9

3.2 Model calibration 9

3.3 Incorporating the COVID-19 pandemic 11

3.4 Modifications to calibration targets 13

3.4.1 Tuberculosis prevalence targets 13

3.4.2 Tuberculosis infection prevalence 15

3.4.3 Tuberculosis case notifications 15

3.4.4 Proportion of previously treated incident cases 17

**4. Policy scenarios** **23**

4.1 No-new-vaccine baseline 23

4.2 Vaccine delivery scenarios 23

4.2.1 Classifying tuberculosis vaccines 23

4.2.2 M72/AS01$_E$ and BCG-revaccination scenarios 24

4.2.3 Vaccine delivery assumptions 24

4.3 Vaccine model structure 26

4.3.1 No Current Infection vaccines 26

4.3.2 Current Infection vaccines 27

4.3.3 Any Infection vaccines 28

**5. Economic analysis methods** **30**

5.1 Calculation of disability-adjusted life years 30

5.2 Tuberculosis-related cost model 30

5.3 Vaccine introduction 30
5.4 Cost-effectiveness analysis and willingness-to-pay thresholds 31
5.5 Total costs from the health-system and societal perspectives 31
**6. Health impact outcomes 32**
**SUPPLEMENTARY RESULTS 33**
**7. No-new-vaccine baselines 33**
7.1 *Status Quo* no-new-vaccine baseline calibration 33
7.2 Posteriors distributions for model parameters 39
7.3 *Strengthened Current Interventions* no-new-vaccine baseline calibration 40
**8. Health impact results 41**
8.1 M72/AS01E scenarios 41
8.2 BCG-revaccination scenarios 44
**9. Economic results 47**
9.1 M72/AS01E scenarios 47
9.2 BCG-revaccination scenarios 53
9.3 Economic results with the *Strengthened Current interventions* no-new-vaccine baseline 59
**References 60**

#### SUPPLEMENTARY METHODS

#### 1. Model structure and equations

We created an age-stratified compartmental differential equation model of tuberculosis in India, including dimensions for age, tuberculosis natural history, vaccination, and access-to-care. The age and access-to-care structures are identical to those included in Clark et al.[1] Minor modifications from the Clark et al. natural history structure are described below. The vaccination structure is in section 4.3.

##### 1.1 Natural history model structure

A natural history structure with eight compartments in Figure S1.1 was created by adapting features of previous models and has been described previously.[1] The latency structure in this model demonstrates a progressive loss of ability to reactivate, with the reactivation rate in the Latent-Fast compartment greater than in Latent-Slow and greater still than in Latent-Zero, where we assume the rate of reactivation is 0. We do not explicitly have a self-clearance compartment. We assume that those in Latent-Fast can only fast progress to subclinical disease or continue to remain latent and transition to Latent-Slow. There is no direct transition between Latent-Fast and Latent-Zero.

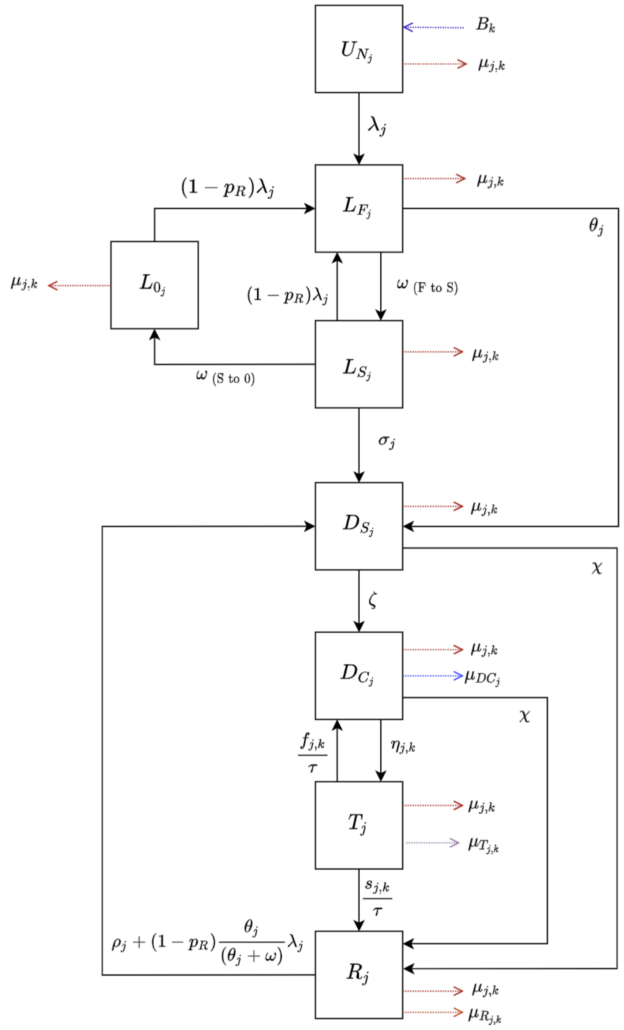


**Figure S1.1 Tuberculosis natural history model structure**

*Abbreviations: $U_N$ = Uninfected-Naive; $L_F$ = Latent-Fast; $L_S$ = Latent-Slow; $L_0$ = Latent-Zero, $D_S$ = Subclinical Disease; $D_C$ = Clinical Disease; T = On-Treatment; R = Recovered. Subscript j represents parameters that vary by age, and subscript k represents parameters that vary over time.*

#### 1.2 Natural history model equations

| | |
|---|---|
| $Age\ j = 0$ | $Age\ j \neq 0$ |
| $\frac{dU_{N_j}}{dt} = B_k - (\lambda_j + \mu_{j,k})U_{N_j}$ | $\frac{dU_{N_j}}{dt} = -(\lambda_j + \mu_{j,k})U_{N_j}$ |

$$\frac{dL_{F_j}}{dt} = \lambda_j U_{N_i} + (1 - p_R)\lambda_j L_{0_i} + [(1 - p_R)\lambda_j]L_{S_i} - (\omega_{FS} + \theta_j + \mu_{j,k})L_{F_i}$$

$$\frac{dL_{S_j}}{dt} = \omega_{FS} L_{F_i} - (\omega_{S0} + \sigma_j + (1 - p_R)\lambda_j + \mu_{j,k})L_{S_i}$$

$$\frac{dL_{0_j}}{dt} = \omega_{S0} L_{S_i} - [(1 - p_R)\lambda_j + \mu_{j,k}]L_{0_i}$$

$$\frac{dD_{S_j}}{dt} = \theta_j L_{F_j} + \sigma_j L_{S_j} + [\rho_j + (1 - p_R)\frac{\theta_j}{\theta_i + \omega}\lambda_j]R_j - (\chi + \zeta + \mu_{j,k})D_{S_j}$$

$$\frac{dD_{C_j}}{dt} = \zeta D_{S_i} + \frac{f_{j,k}}{\tau}T_j - (\chi + \eta_{j,k} + \mu_{DC_i} + \mu_{j,k})D_{C_i}$$

$$\frac{dT_j}{dt} = \eta_{j,k} D_{C_j} - \left(\frac{s_{j,k} + f_{j,k}}{\tau} + \mu_{T_{j,k}} + \mu_{j,k}\right)T_j$$

$$\frac{dR_j}{dt} = \frac{s_{j,k}}{\tau}T_j + (D_{S_j} + D_{C_j})\chi - [\rho_j + (1 - p_R)\frac{\theta_j}{\theta_i + \omega}\lambda_j + \mu_{R_j} + \mu_{j,k}]R_j$$

#### 2. Natural history

##### 2.1 Natural history parameter values and data sources

Parameters used in the natural history model structure are provided in Table S2.1 below, along with their definitions, sources, and information on whether the parameter is fixed or varied (as well as whether they are varied by age or time) during calibration. Further details about how the age varying parameters are implemented are provided in section 2.2, and further details on parameters related to treatment are provided in section 2.3. The parameter ranges provided for the tuberculosis natural history parameters are priors fitted during calibration in a Bayesian analysis. We assume that all values within the prior range are equally likely. The prior ranges were pre-specified based on literature review and were reviewed as new data became available.

**Table S2.1 India national model parameter values and sources**

| Description | Units | Symbol | Prior | Fixed or Varying During Calibration | Age Varying | Time Varying | Source |
|---|---|---|---|---|---|---|---|
| ***Births and deaths (excluding on-treatment mortality)*** | | | | | | | |
| Birth rate | Per year | $B_k$ | United Nations World Population Prospects population estimates and projections | Fixed | No | Yes | [2] |
| Background mortality rate | Per year | $\mu_{j,k}$ | Calculated in the model from United Nations population estimates and projections | Fixed | Yes, age specific mortality rates from demographic dataset | Yes | [2] |
| Mortality rate for clinical tuberculosis disease | Per person per year | $\mu_{DC_j}$ | (0–0.178) | Varying | Yes, value for children is greater than value for adults | No | [3] |
| Mortality rate post-tuberculosis disease | Per person per year | $\mu_{R_j}$ | $0.22\mu_{j,k}$ | Fixed relationship | Yes because $\mu_{j,k}$ varies | Yes because $\mu_{j,k}$ varies | [4] |
| ***Natural History*** | | | | | | | |
| Force of infection | Per year | $\lambda_j$ | Fitted | Fixed Equation | Yes, age specific contact rates[5] | No | *Calculated* |
| Probability of transmission per infectious contact | - | $p_T$ | (0–0.0068) | Varying | No | No | *Assumed* |

| | | | | | | | |
|---|---|---|---|---|---|---|---|
| Fraction of total tuberculosis that is extrapulmonary | - | $ep$ | 0.222 | Fixed | No | No | [6,7] |
| Infectiousness of subclinical relative to clinical tuberculosis | - | $r$ | 0.83 | Fixed | No | No | [8] |
| Rate of fast progression to disease, by age | Per person per year | $\theta_j$ | (0.0696–0.111) | Varying | Yes; Retain if value for children is **less** than value for adults. | No | [9] |
| Rate from $L_F$ to $L_S$ | Per person per year | $\omega_{FS}$ | 0.5 | Fixed | No | No | *Defined* |
| Rate of reactivation from $L_S$, by age | Per person per year | $\sigma$ | (0.000135–0.00113) | Varying | Yes; Retain if value for children is **less** than value for adults. | No | [9] |
| Rate from $L_S$ to $L_0$ | Per person per year | $\omega_{S0}$ | (0.0254–0.0467) | Varying | No | No | [9] |
| Rate of progression from $D_S$ to $D_C$ | Per person per year | $\zeta$ | (0–12) | Varying | No | No | *Assumed* |
| Rate of natural cure from $D_C$ and $D_S$ | Per person per year | $\chi$ | (0.10–0.25) | Varying | No | No | [10,11] |
| Rate of relapse from R, by age | Per person per year | $\rho_j$ | (0.0001–0.07) | Varying | Yes; Retain if value for children is **less** than value for adults. | No | [12–14] |
| ***Protection Parameters*** | | | | | | | |
| Protection from reinfection $L_S$, $L_F$, $L_0$, R | - | $p_R$ | (0.60–0.85) | Varying | No | No | [10,11,15–17] |
| Access-to-care parameter | - | $p_E$ | (0–1) | Varying | No | No | *Assumed* |

#### 2.2 Operationalising age varying parameters

We assume that aspects of tuberculosis natural history and mortality vary by age. This is implemented by stratifying certain natural history parameters by age and applying age-specific prior ranges and relative constraints during calibration.[18] The following table describes the method used to operationalise the age varying differences in parameters between adults, defined as all ages greater than and equal to 15, and children, defined as all ages less than 15. For the rate per year of reactivation, relapse, and fast progression to tuberculosis disease, we assume that the rate for children is less than that for adults. For mortality rates, we assume the opposite: the rate for children is higher than that for adults.

**Table S2.2 How age varying parameters are operationalized**

| Parameter | Range | Age Varying Description | Age Scaling Parameter | Adults ($\theta_{A15}$) | Children ($\theta_{A0}$) |
|---|---|---|---|---|---|
| $\theta_j$ Rate per year of fast progression | $(0.0696–0.111)$ | Retain if value for children is **less** than value for adults | Sample $j_1$ from $(0–1)$ | Sample $\theta_{A15}$ from $(0.0696–0.111)$ | $\max(0.0696, \theta_{A15} \times j_1)$ |
| $\sigma_j$ Rate per year of reactivation | $(0.000135–0.00113)$ | Retain if value for children is **less** than value for adults | Sample $j_2$ from $(0–1)$ | Sample $\sigma_{A15}$ from $(0.000135–0.00113)$ | $\max(0.000135, \sigma_{A15} \times j_2)$ |
| $\rho_j$ Rate per year of relapse | $(0.01–0.07)$ | Retain if value for children is **less** than value for adults | Sample $j_3$ from $(0–1)$ | Sample $\rho_{A15}$ from $(0.01–0.07)$ | $\max(0.01, \rho_{A15} \times j_3)$ |
| $\mu_{DC_j}$ Clinical TB mortality rate per year | $(0–0.178)$ | Retain if value for children is **greater** than value for adults | Sample $S_{Age}$ from $(0–1)$ | $\mu_{DC_{A0}} \times S_{Age}$ | Sample $\mu_{DC_{A0}}$ from $(0–0.178)$ |
| $\mu_{T_j} = \frac{\kappa_j}{\tau}$ On-treatment mortality rate per year | $(0–0.270)$ | Retain if value for children is **greater** than value for adults | Sample $S_{Age}$ from $(0–1)$ | $\frac{\kappa_{A0}}{\tau} \times S_{Age}$ | Sample $\kappa_{A0}$ from $(0–0.135)$ |

##### 2.3 Treatment initiation and outcomes

Steps for calculating treatment initiation, treatment completion, non-completion, and mortality rates are described in the Supplementary Material for Clark et. al.[1] We assume the SFR is the ratio between treatment completions to the sum of treatment completions and non-completions. In India, $\mathrm{SFR} = 0.96$. The data used to calculate the on-treatment outcomes was obtained from the WHO. However, as the private sector accounts for a substantial portion of treatments in India, and not all of the treatments conducted in the private sector are reported, we make adjustments to the on-treatment completion and non-completion fractions from Table S2.3 as described below and in Table S2.4.

**Table S2.3 Calculating treatment outcome parameter values for adults and children**

| Parameter | Adults | Children |
|---|---|---|
| $\kappa_j$<br>On-treatment mortality fraction | $\kappa_{A0} \times S_{Age}$ | Sample $\kappa_{A0}$ from $(0–0.135)$ |
| $s_j$<br>On-treatment completion fraction | $(1 - \kappa_{A15})\mathrm{SFR}$ | $(1 - \kappa_{A0})\mathrm{SFR}$ |
| $f_j$<br>On-treatment non-completion fraction | $(1 - \kappa_{A15})(1 - \mathrm{SFR})$ | $(1 - \kappa_{A0})(1 - \mathrm{SFR})$ |

We assume that the total number of treatments is composed of the treatments that are reported and the treatments that are not reported. We assume that the on-treatment mortality fraction is the same in the public and private sector, but want to adjust the treatment completion and non-completion rates to account for differences between those reported and those not reported as in Table S2.4.

**Table S2.4 Calculation of treatment outcomes for India by year**

| Description | Symbol | Year $(k)$ | | | | | | | | |
|---|---|---|---|---|---|---|---|---|---|---|
| | | ≤2012 | 2013 | 2014 | 2015 | 2016 | 2017 | 2018 | 2019 | ≥2020 |
| Fraction of total treatments reported | $f_{T,k}$ | 0.60 | 0.63 | 0.68 | 0.67 | 0.73 | 0.77 | 0.80 | 0.83 | 0.87 |
| On-treatment mortality rate | $\frac{\kappa_j}{\tau}$ | $\frac{\kappa_j}{\tau}$<br>Sample $\kappa_{A0}$ from $(0, 0.135)$, then $\kappa_{A15} = \kappa_{A0} \times S_{Age}$ | | | | | | | | |
| On-treatment completion rate | $\frac{s_j}{\tau}$ | $\frac{f_{T,k}0.96(1-\kappa_j)+(1-f_{T,k})0.40}{\tau}$ | | | | | | | | |
| On-treatment non-completion rate | $\frac{f_j}{\tau}$ | $\frac{f_{T,k}0.04(1-\kappa_j)+(1-f_{T,k})(0.60-\kappa_j)}{\tau}$ | | | | | | | | |

We assume that 60% of the total treatment occurs in the public sector and the remaining 40% occurs in the private sector. We assume that all treatments not reported are from the private sector, that the treatment completion rate in the private sector is 40%, and that there is no reporting bias (in that they were equally likely to not report treatment completions or non-completions or deaths). Before 2012, only the treatment conducted in the public sector was reported, but since then, treatment in the private sector has begun to be reported.[19]

#### 3. Model simulation and calibration

##### 3.1 Model simulation

We specified a system of ordinary differential equations defining the derivatives with respect to time of a set of state variables, to simulate the country-specific tuberculosis epidemic between 1900 and 2050. We initialised the simulation by distributing the population between the eight tuberculosis natural history states using a fitted parameter representing the proportion of the population uninfected at the start of the simulation. For each year of the simulation (1900–2050), our models are designed to exactly match the age and country-specific UN population estimates and projections. Forty percent of the population was assigned to the low access-to-care stratum and the remaining sixty percent of the population was assigned to the high access-to-care stratum.

##### 3.2 Model calibration

For this India modelling analysis, we followed the same modelling approach as in Clark et al.[1]

Broadly, this was as follows:

1. Construct a mechanistic model
2. Calibrate the model by identifying areas of the input parameter space where the output of the mechanistic model was consistent with the historical epidemiologic data
3. Use the calibrated model to simulate and predict future tuberculosis epidemiology and model new vaccines

In the context of this analysis, step 1 was achieved by creating the compartment differential equation model as specified in Section 1. For step 2, we independently calibrated a model by identifying areas of the parameter space that made the output of the model match the corresponding calibration targets (Table S3.1 below). Further details on the sources for the calibration targets and any additional modifications are in the subsequent sections.

The model was fitted to the calibration targets using history matching with emulation, a method that allows us to explore high-dimensional parameter spaces efficiently and robustly.[20–23] History matching progresses as a series of iterations, called waves, where implausible areas of the parameter space, i.e., areas that are unable to give a match between the model output (e.g., the predicted incidence rate by the model) and the empirical data (e.g., the incidence rate calibration target from the WHO data), are found and discarded. In order to identify implausible parameter sets, emulators, which are statistical approximations of model outputs that are built using a modest number of model runs, are used. Emulators provide an estimate of the value of the model at any parameter set of interest, with the advantage that they are orders of magnitude faster than the model.

History matching with emulation, implemented through the *hmer* package in R,[24,25] considerably reduced the size of the parameter space to investigate. Rejection sampling was then performed on the reduced space to identify at least 1000 parameter sets that matched all targets.

If we were unable to find at least 1000 fully fitted parameter sets using history matching with emulation, we subsequently used an Approximate Bayesian Computation using Markov Chain Monte Carlo method (ABC-MCMC). ABC-MCMC was conducted using the *easyABC* package in R, modified by the Sebastian Funk, Gwenan Knight, and the Tuberculosis Modelling group at LSHTM for adaptive sampling and to accept seeded parameter values.[26,27] We used parameter sets with the maximum number of targets fitted using history matching with emulation as starting seeds for multiple MCMC chains per country, with the ABC-MCMC algorithm continuously adapting using the last 1000 points, a burn in of 1000 samples, and the noise factor set to 0.0001.

Once we had obtained 1000 parameter sets that produced output consistent with the calibration targets, we used those parameter sets with the mechanistic model to simulate the future (step 3) for each country as the *Status Quo* no-new-vaccine baseline, where we assumed that current trends and quality of non-vaccine tuberculosis services continued into the future at the same rate.

As an alternative future, we calibrated a *Strengthened Current Interventions* no-new-vaccine baseline. This baseline assumed a scale up in other non-vaccine tuberculosis interventions between 2021 and 2035 in order to meet the target of a 50% reduction in tuberculosis incidence in 2035 compared to the 2015 estimates (an incidence rate of 108.5 (64–164 per 100,000 population). This scale-up was introduced in the model by introducing parameters (sampled between 0 and 1) which act as multipliers on the rate of progression to disease and in the force of infection equation.

The process of generating fits for the *Strengthened Current Interventions* no-new-vaccine baseline while capturing uncertainty was as follows:

1. Obtain 1000 full fits from the *Status Quo* baseline
2. Subset the 1000 *Status Quo* full fits to 100 by:
    a. Ranking the 1000 *Status Quo* fits from smallest to largest tuberculosis incidence rate in 2020
    b. Retaining every 10th parameter set
3. Use emulation on each of the 100 parameter sets
4. Obtain 100 "groups" of fully fitting parameter sets (one group for each original parameter)
5. Subset each group of fully fitting parameter sets to 10 by:
    a. Ranking the parameter sets in each group from smallest to largest tuberculosis incidence rate in 2035
    b. Retaining every nth parameter set to obtain 10 across the range
6. Obtain 1000 full fits for the *Strengthened Current Interventions* baseline by combining the 10 parameter sets from each of the 100 emulation sets

##### Table S3.1 India national model calibration targets

| Calibration Targets | Year | Age (years) | Estimate | Lower | Upper |
|---|---|---|---|---|---|
| Tuberculosis incidence rate (per 100,000 population/year) | 2000[28] | All | 289 | 149 | 473 |
| | 2020[28] | All | 188 | 129 | 257 |
| | | 0-14 | 91 | 56 | 126 |
| | | ≥15 | 224 | 138 | 310 |
| | 2025[29] | All | 212 | 145 | 293 |
| Tuberculosis mortality rate (per 100,000 population/year) | 2000[6] | All | 67 | 57 | 79 |
| | 2020[6] | All | 37 | 34 | 40 |
| | 2025[29] | All | 36 | 33 | 39 |
| Tuberculosis case notification rate (per 100,000 population/year) | 2000[2,7] | All | 177 | 142 | 212 |
| | 2020[2,7] | All | 136 | 109 | 163 |
| | | 0-14 | 33 | 26 | 40 |
| | | ≥15 | 173 | 138 | 208 |
| Active tuberculosis prevalence (per 100,000 population) | 2015[30,31] | All | 315 | 210 | 529 |
| | 2021[32] | All | 312 | 218 | 406 |
| | 2021[32] | ≥15 | 394 | 276 | 512 |
| Tuberculosis infection prevalence proportion | 2021[32] | All | 0.314 | 0.114 | 0.514 |
| Access-to-care tuberculosis prevalence ratio | 2020[33] | All | 0.427 | 0.327 | 0.527 |
| Subclinical tuberculosis prevalence ratio | 2020[34] | All | 0.504 | 0.361 | 0.797 |
| Proportion of incident tuberculosis cases having previously been treated | 2020 | All | 0.191 | 0.139 | 0.241 |

#### 3.3 Incorporating the COVID-19 pandemic

It will be a number of years before the full implications of disruptions to tuberculosis prevention and care during the COVID-19 pandemic are realised. The WHO provided estimates for the impact on the tuberculosis incidence and mortality rates between 2020–2025 relative to January 2020, shown in Figure S3.2.[29]

In order to ensure that the model is appropriately representing the future trends in incidence and mortality, we calibrated to the projected incidence and mortality for 2025, which is estimated as a 10% increase in both mortality and incidence in 2025 compared to January 2020. To implement this, we calculated a 10% increase to the incidence and mortality rates estimated by the WHO in 2019, shown in Table S3.2, and calibrated to both for 2025.

*A–Impact on tuberculosis incidence rate* *B–Impact on tuberculosis mortality rate*

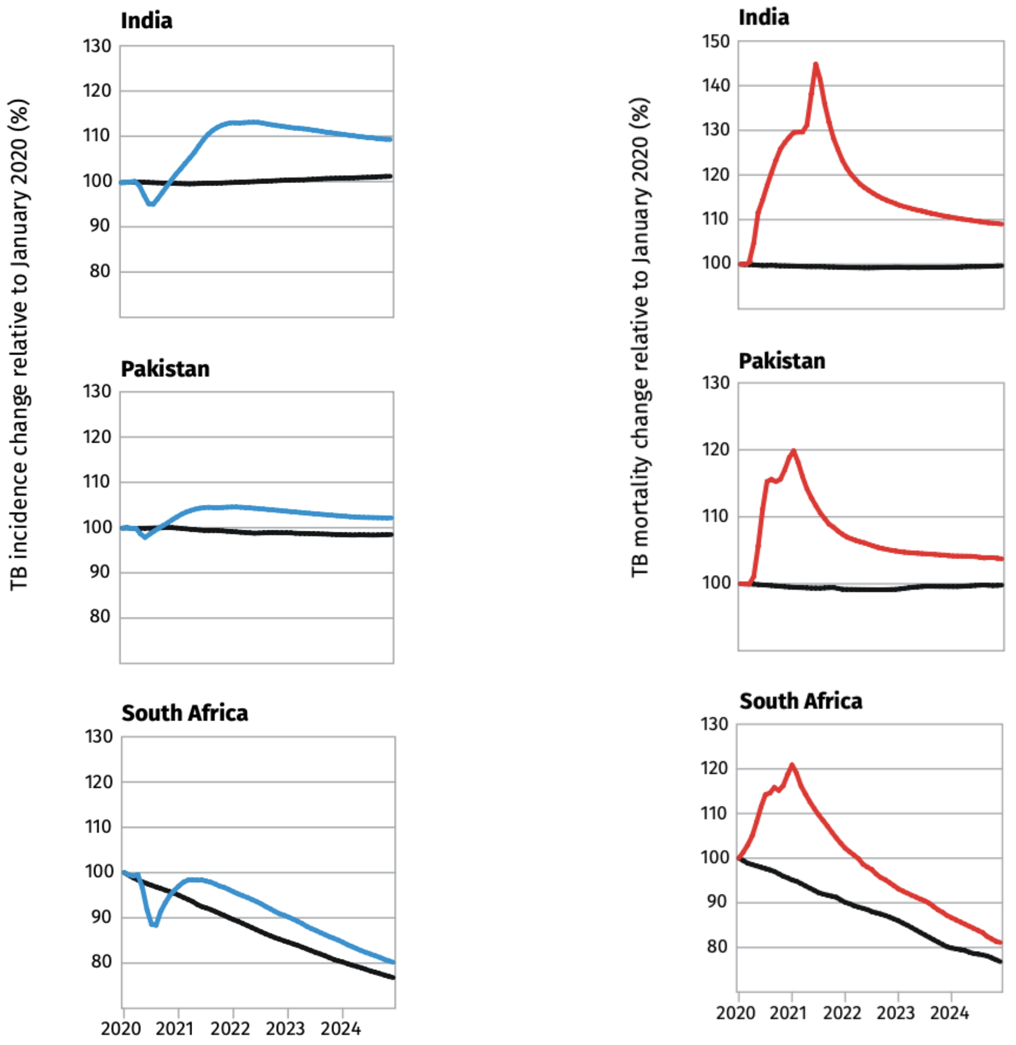


**Figure S3.2** **The estimated impact of the COVID-19 pandemic on the (A) tuberculosis incidence and (B) mortality rates from the WHO Global Tuberculosis Report 2021[29]**

**Table S3.2** **Incidence and mortality rate targets for all ages for 2025**

| **Year** | **Incidence rate (per 100,000)** | **Mortality rate (per 100,000)** |
|---|---|---|
| 2019 | 193<br>(126–260) | 33<br>(30–35) |
| 2025<br>(10% increase from 2019) | 212<br>(145–293) | 36<br>(33–39) |

##### 3.4 Modifications to calibration targets

###### 3.4.1 Tuberculosis prevalence targets

***i. Adjusting the 2015 target bounds***

We obtained an estimate for the tuberculosis prevalence in 2015 from "Estimating tuberculosis incidence from primary survey data: a mathematical modelling approach" by Pandey et al 2017.[30] In it, they estimate the prevalence of smear-positive cases across all ages in India, as well as the proportion of cases that are smear-positive. The mean and 95% confidence intervals for the estimates of these are 159.38 (122.9–196.59) and 0.63 (0.43–0.93) respectively.[30] Cited sources within the paper suggest that these quantities have been modelled as lognormal (smear-positive prevalence) and beta (smear-positive proportion) distributed.[30] The total prevalence, therefore, can be determined as

$$\text{Pulmonary tuberculosis prevalence} = \frac{\text{Smear-positive prevalence}}{\text{Smear-positive proportion}}$$

If we assume that the mean estimate for the proportion of cases that are smear-positive is accurate, then we simply quotient the smear-positive bounds by this value (0.63). This gives

$$\text{Pulmonary tuberculosis prevalence} \ = \ 253.0 \ (195.1\text{–}312.0)$$

Since we have confidence intervals and a knowledge of the underlying distributions, we can attempt to determine the hyperparameters of the distributions. Once we have these, we can sample repeatedly from the quotient of the two distributions to get an estimate for its confidence interval. We sample from the numerator's distribution, sample from the denominator's distribution, and quotient them to represent a sample from the (unknown) prevalence distribution. Given enough samples, we can obtain a reasonable estimate of the confidence interval. Since the lognormal distribution has a closed-form, we can simply solve for the hyperparameters.

$$\text{Smear-positive prevalence} \sim \text{lognorm}(\mu = 5.046,\ \sigma^2 = 0.01436)$$

The beta-distribution is less straightforward, but we can use maximum likelihood estimation to find feasible parameter values. Doing so gives

$$\text{Smear}-\text{positive proportion} \sim \text{beta}(\alpha = 10, \beta = 6)$$

Then we perform monte-carlo sampling to generate a representative sample from our quotient distribution, from which we obtain a 95% confidence interval.

$$\text{Pulmonary tuberculosis prevalence} = \ 253.0 \ (169.3\text{–}424.7)$$

***ii. Adjusting for extrapulmonary tuberculosis***

In our model we are representing everyone with tuberculosis, which includes both pulmonary (PTB) and extrapulmonary tuberculosis (EPTB). EPTB is not infectious but is included in the WHO estimates of yearly incidence and mortality rates. The 2021 prevalence estimates from the National Tuberculosis Prevalence Survey did not adjust for EPTB in the estimate provided for adults, and neither did the 2015 study which estimated the tuberculosis prevalence from subnational surveys. Therefore, we want to adjust the PTB prevalence estimates and range by the amount of EPTB in order to estimate the total TB prevalence. To estimate the proportion of EPTB, we used the average of the proportion of incident extrapulmonary tuberculosis cases from 2013–2020 (Table S3.3).

**Table S3.3 Number of incident tuberculosis cases by year in India**

| Year | New EPTB cases | Relapse EPTB cases | Total incident EPTB cases | Total incident cases | Proportion incident EPTB cases |
|---|---|---|---|---|---|
| 2013 | 226,557 | – | 226,557 | 1,243,905 | 0.18 |
| 2014 | 275,502 | – | 275,502 | 1,609,547 | 0.17 |
| 2015 | 298,831 | – | 298,831 | 1,667,136 | 0.18 |
| 2016 | 281,162 | – | 281,162 | 1,763,876 | 0.16 |
| 2017 | 276,786 | 3,067 | 279,853 | 1,649,694 | 0.17 |
| 2018 | 380,904 | 5,622 | 386,526 | 1,908,683 | 0.20 |
| 2019 | 476051 | 3,862 | 479,913 | 2,162,323 | 0.22 |
| 2020 | 471,000 | 4,034 | 475,034 | 1,629,301 | 0.29 |

*Abbreviations: EPTB = extrapulmonary tuberculosis*

From the National tuberculosis prevalence survey India 2019–2021, the prevalence of microbiologically confirmed pulmonary tuberculosis among population aged ≥ 15 years in India was estimated at 316 (290–342) [This adjusted prevalence was estimated using a robust standard errors model with imputation and inverse probability weighting].[32] Averaging the proportion of incident EPTB cases column in Table S3.3 and dividing the estimates and bounds on the pulmonary tuberculosis prevalence estimates by (1- average of proportion of incident EPTB cases) we obtain the following as the estimates of the tuberculosis prevalence per 100,000 population:

$$\text{All ages tuberculosis prevalence in 2015} = 315.1\ (210.8\text{–}529.0)$$

$$\text{Adult tuberculosis prevalence in 2021} = 393.6\ (361.2\text{–}426.0)$$

##### *iii. Adjusting the 2021 target bounds*

The National Tuberculosis Prevalence Survey India 2019–2021 reports estimates for the prevalence of all forms of tuberculosis among all age groups in India (312.0 [286.0–337.0] per 100,000 population) and the prevalence of microbiologically confirmed pulmonary tuberculosis among adults aged ≥ 15 years in India (316.0 [290.0–342.0] per 100,000 population). As described in the previous section, we adjusted the estimate of the prevalence of pulmonary tuberculosis in adults for EPTB, giving a revised estimate for the prevalence of all forms of tuberculosis disease in adults of 393.6 (361.2–426.0). We subsequently increased the upper and lower bounds on the all age and adult targets by 30%, leading to estimates of 312.0 (218.4–405.6) and 393.6 (275.5–511.7) respectively. Rationale for adjusting the bounds on the targets is described below.

*Rationale 1: Impact of the Covid-19 pandemic*

Some state groups started and completed the survey before the COVID-19 pandemic, others during, others after the major pandemic waves had completed.[32] Depending on the impact of COVID-19 measures on tuberculosis, this could bias the estimates of the region either up or down, and bias the overall estimate of the tuberculosis prevalence for India, particularly as Delhi (the region with the highest estimated tuberculosis prevalence) started and completed the survey before the pandemic.[32]

*Rationale 2: Differences between planned surveyed clusters and actual surveyed clusters*

The National Tuberculosis Prevalence Survey India 2019–2021 compares the number of pulmonary tuberculosis cases notified at the state group level in 2019, 2020 and 2021 between those clusters who were surveyed and those who were not surveyed.[32] Although no statistically significant differences were observed between the surveyed and not-surveyed clusters, there are qualitative differences between the number of notifications of pulmonary tuberculosis between groups, where non-surveyed clusters consistently have a lower number of notifications.[32]

##### 3.4.2 Tuberculosis infection prevalence

###### *i. Adjusting the 2021 target bounds*

The National Tuberculosis Prevalence Survey India 2019–2021 reports an estimate for the prevalence of tuberculosis infection in India among adults of 0.314 (0.272–0.353). We adjusted the bounds to give a revised estimate of 0.314 (0.114–0.514), with rationale described below.

*Rationale 1: Oversampling from Gujarat with no adjustment*

Of the 55 clusters where IGRA testing was done, 31 were in Gujarat and 24 were in the remaining 19 state groups. Gujarat had the lowest estimated tuberculosis prevalence per 100,000 population.[32] If we assume that prevalence of tuberculosis infection is correlated with prevalence of tuberculosis disease, then we would anticipate that the tuberculosis infection prevalence estimates from Gujarat would be commensurately low. As more than half of the clusters were from Gujarat, and there is no indication of adjustment for oversampling from this region, it is possible that the reported country-level tuberculosis infection prevalence is an underestimate. If our assumption that prevalence of infection correlates with prevalence of disease was incorrect, the tuberculosis infection prevalence estimates may actually be overestimated. As such, we have adjusted the bounds to account for oversampling with no adjustment, but retained the central estimate, resulting in a calibration target of 0.314 (0.114–0.514).

##### 3.4.3 Tuberculosis case notifications

###### *i. Adjusting to account for the private sector contribution to reported case notifications*

Treatment in India can occur in the public or private sector. While this varies by state, it is estimated that 60% of treatment is performed in the public sector, and the remaining 40% in the private sector. According to the WHO Global TB Report 2019, reported case notifications only included notifications from the public sector before 2013.[19] From 2013-2020, reported case notifications began to include the private sector (Figure S3.4). By 2020, approximately 31% of the total reported notifications were from the private sector.[19]

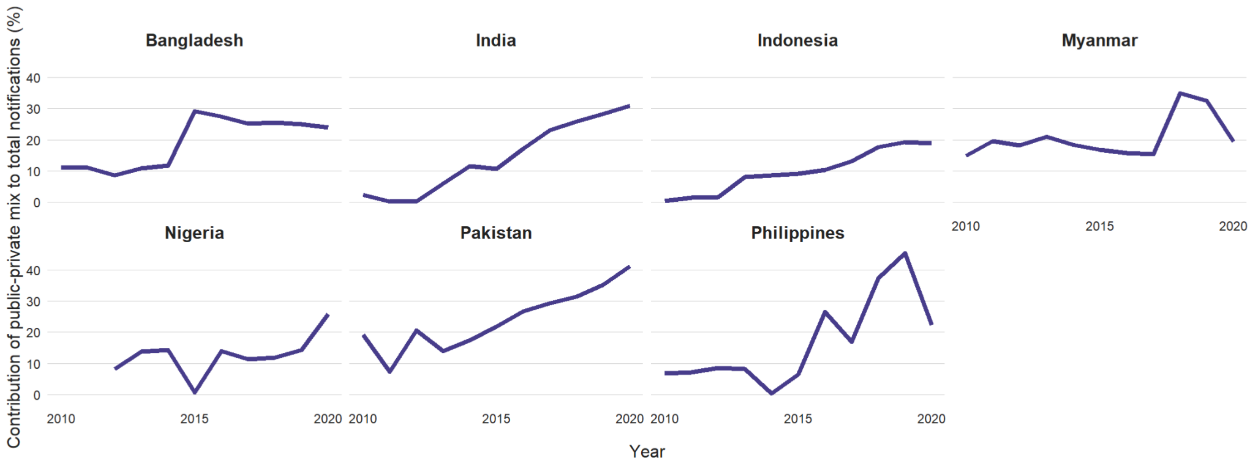


**Figure S3.4 Contribution of the private sector to reported case notifications from WHO Global TB Report 2021[29]**

The model represents case notifications as the number of tuberculosis treatment initiations. We want to calibrate the model to the *true* number of treatment initiations, as this is what the model will represent. Therefore, this involves adjusting the WHO reported case notifications to reflect underreporting from the private sector. To do this, we must calculate the fraction of total cases notifications (treatment initiations) that are reported, while accounting for both the private and public sector.

$\mathrm{RN}_{\mathrm{Total}}$ = Total Reported Notifications
$\mathrm{RN}_{\mathrm{Public}}$ = Public Sector Reported Notifications
$\mathrm{RN}_{\mathrm{Private}}$ = Private Sector Reported Notifications
$\mathrm{RN}_{\mathrm{Total}} = \mathrm{RN}_{\mathrm{Public}} + \mathrm{RN}_{\mathrm{Private}}$

Let: $f_{\mathrm{Private}} = \frac{\mathrm{RN}_{\mathrm{Private}}}{\mathrm{RN}_{\mathrm{Total}}}$ be the fraction of reported notifications that come from the private sector

Using the percent contribution of the private sector to the reported treatments, and the assumption that *all* treatments occurring in the public sector are reported, we can calculate the fraction of total notifications that are actually reported.

$\mathrm{N}_{\mathrm{Total}}$ = Total Notifications
$\mathrm{N}_{\mathrm{Public}}$ = Public Sector Notifications
$\mathrm{N}_{\mathrm{Private}}$ = Private Sector Notifications
$\mathrm{N}_{\mathrm{Total}} = \mathrm{N}_{\mathrm{Public}} + \mathrm{N}_{\mathrm{Private}}$

Let: $r_{f,k} = \frac{\mathrm{RN}_{\mathrm{Private}}}{\mathrm{N}_{\mathrm{Private}}}$ be the fraction of total private sector notifications reported in year $k$

$$\mathrm{RN}_{\mathrm{Total}} = \mathrm{N}_{\mathrm{Public}} + r_{f,k} \times \mathrm{N}_{\mathrm{Private}} \quad (1)$$

$$f_{\mathrm{Private}} \times \mathrm{RN}_{\mathrm{Total}} = r_{f,k} \times \mathrm{N}_{\mathrm{Private}} \quad (2)$$

$$\mathrm{RN}_{\mathrm{Total}} = \frac{r_{f,k} \times \mathrm{N}_{\mathrm{Private}}}{f_{\mathrm{Private}}} \quad (3)$$

Sub (3) into (1)

$$\frac{r_{f,k} \times \mathrm{N}_{\mathrm{Private}}}{f_{\mathrm{Private}}} = \mathrm{N}_{\mathrm{Public}} + r_{f,k} \times \mathrm{N}_{\mathrm{Private}}$$

Solve for $r_{f,k}$

$$r_{f,k} = \frac{\mathrm{N}_{\mathrm{Public}}}{\mathrm{N}_{\mathrm{Private}}(\frac{1}{f_{\mathrm{Private}}} - 1)}$$

$$\therefore r_{f,k} = \frac{60}{40(\frac{1}{f_{\mathrm{Private}}} - 1)}$$

Note: This calculation is valid for $f_{\mathrm{Private}} > 0$

We want to calculate $f_T$ = fraction of total notifications reported

$$f_T = \frac{\mathrm{RN}_{\mathrm{Total}}}{\mathrm{N}_{\mathrm{Total}}}$$

$$f_T = \frac{\mathrm{RN}_{\mathrm{Public}} + \mathrm{RN}_{\mathrm{Private}}}{\mathrm{N}_{\mathrm{Total}}}$$

Assume:

$$\mathrm{RN}_{\mathrm{Public}} = \mathrm{N}_{\mathrm{Public}} = 60$$

$$N_{\mathrm{Total}} = 100$$

$$f_T = \frac{60 + r_{f,k} \times \mathrm{N}_{\mathrm{Private}}}{100}$$

Using the derived equation, we can calculate the fraction of total notifications reported from 2013–2020 (Table S3.4).

**Table S3.4 The fraction of tuberculosis treatment notifications in India from the private sector and overall**

| Year (k) | Fraction of reported notifications that came from the private sector $f_{\text{Private},k}$ | Fraction of total possible private sector notifications that were reported $r_{f,k}$ | Fraction of total notifications reported $f_{T,k}$ |
|---|---|---|---|
| ≤ 2012 | 0 | 0 | 0.60 |
| 2013 | 0.05 | 0.08 | 0.63 |
| 2014 | 0.12 | 0.20 | 0.68 |
| 2015 | 0.10 | 0.17 | 0.67 |
| 2016 | 0.18 | 0.33 | 0.73 |
| 2017 | 0.22 | 0.42 | 0.77 |
| 2018 | 0.25 | 0.50 | 0.80 |
| 2019 | 0.28 | 0.58 | 0.83 |
| 2020 | 0.31 | 0.674 | 0.87 |

To adjust the WHO reported case notification estimates for underreporting, we divide the estimates by the fraction of total treatments reported ($f_T$), and assume 20% upper and lower uncertainty bounds. The reported and adjusted estimates of case notifications are provided in Table S3.5 and assume 20% upper and lower uncertainty bounds. The reported and adjusted estimates of case notifications are provided in Table S3.5.

**Table S3.5 The WHO reported and adjusted tuberculosis case notification targets for India**

| Year | WHO reported case notification value | Adjusted case notification value | Low bound (Adjusted value × 0.8) | High bound (Adjusted value × 1.2) |
|---|---|---|---|---|
| 2000 | 106 | 177 | 142 | 212 |
| 2020 | 118 | 136 | 109 | 163 |

##### 3.4.4 Proportion of previously treated incident cases

***i. Adjusting the proportion retreated bounds***

The proportion retreated target is included to ensure that the disease tuberculosis incidence is derived from the correct source (i.e., to ensure that we do not overestimate the amount of incidence that is coming from fast-progression or reactivation without treatment).

The data available from the WHO are:

1. Number of case notifications (i.e., the notified treatment initiations), per year
2. Number of case notifications who are people who have been previously treated, per year

By dividing the number of notifications who are people who have been previously treated (2) by the total number of notifications (1), we get the proportion of notifications that have been previously treated.

$$= \frac{\text{Number of notifications who are people who have been previously treated per year}}{\text{Number of notifications per year}}$$

$$= \text{Proportion of notifications that have been previously treated}$$

We assume that at equilibrium, the proportion of notifications who have been previously treated will be equal to the proportion of incident disease cases who have been treated previously.

The estimate of the proportion of notifications that have been previously treated for India from the WHO dataset is 10.0% (4.3–14.7). However, country specific estimates may be subject to recall bias as they rely on patients to accurately report previous treatment. Additionally, studies have shown that approximately 11% of patients recorded as "new" have had some form of previous tuberculosis treatment.[35] Therefore, we adjusted the estimates from the WHO dataset, and calibrated to a target of 19.1% (13.9–24.1).

##### *ii. Calculating the proportion retreated target in the model*

The subsequent pages describe the methods used to calculate the proportion retreated target.

**Definition 1:** The number of notifications, per year is the flow from Dc and T = $\eta \times D_C$

**Definition 2:** Being "previously treated" implies that an individual arrived in the R compartment via the T compartment.

**Definition 3:** For an individual to count as a notification of a person who was previously treated (Definition 2), they must flow from T → R → Ds → Dc → T

Looking at the total number of notifications broken down to their origins, we see that:

$$\begin{aligned}&\text{Total number of notifications per year} =\\&\text{Number of notifications of people who were in Ls per year}\\&\qquad + \text{Number of notifications of people who were in Lf per year}\\&\qquad + \text{Number of notifications of people who were in R per year}\end{aligned}$$

The "Notifications of people who were in R" term is further broken down into:

- People who entered R from T, per year
- People who entered R from Dc, per year
- People who entered R from Ds, per year

We can rewrite the total number of notifications per year equation as:

$$\begin{aligned}&\text{Total number of notifications per year} =\\&\text{Number of notifications of people who were in Ls per year}\\&+ \text{Number of notifications of people who were in Lf per year}\\&+ \text{Number of notifications of people who were in R having entered R from Ds per year}\\&+ \text{Number of notifications of people who were in R having entered R from Dc per year}\\&+ \text{Number of notifications of people who were in R having entered R from T per year}\end{aligned}$$

Recall now what we are looking to calibrate to:

$$= \frac{\text{Number of notifications who are people who have been previously treated per year}}{\text{Number of notifications per year}}$$

The denominator is directly available from the model: the total number of notifications ($\eta \times D_C$). Using definitions 2 and 3 above, the "number of notifications who are people who have been previously treated per year" = "notifications of people who were in R having entered R from T per year". Therefore, we can redefine our calibration target as:

$$= \frac{\text{Number of notifications of people who were in R having entered R from T per year}}{\text{Total number of notifications per year}}$$

We do not have **notifications** disaggregated by source, but we do have **incidence** disaggregated by source. Incident cases are defined as the flow into Ds, which can be from R, from Ls, or from Lf.

$$\begin{aligned}&\text{Total number of incident cases per year} = \\ &\text{Total number of incident cases of people from Lf per year} \\ &+ \text{Total number of incident cases of people from Ls per year} \\ &+ \text{Total number of incident cases of people from R per year}\end{aligned}$$

We obtain output on all of these flows, so we can calculate the proportion of incident cases from each pathway (Lf, Ls, and R) easily by just dividing the total number of incident cases of people from Lf, Ls or R by the total number of incident cases.

The proportions of incidence from each pathway are:

$$\frac{\text{RD}}{(\text{RD} + \text{LsD} + \text{LfD})} \qquad \frac{\text{LfD}}{(\text{RD} + \text{LsD} + \text{LfD})} \qquad \frac{\text{LsD}}{(\text{RD} + \text{LsD} + \text{LfD})}$$

*RD = flow from R to Ds = total number of incident cases of people from R, per year*
*LsD = flow from Ls to Ds = total number of incident cases of people from Ls, per year*
*LfD = flow from Lf to Ds = total number of incident cases of people from Lf, per year*
*RD + LsD + LfD = total number of incident cases, per year*

Similarly, we can disaggregate the flow from R to Ds further into how the people in R entered R.

$$\begin{aligned}&\text{Total number of incident cases of people from R per year} = \\ &\text{Total number of incident cases of people from R who entered R from T per year} \\ &+ \text{Total number of incident cases of people from R who entered R from Dc per year} \\ &+ \text{Total number of incident cases of people from R who entered R from Ds per year}\end{aligned}$$

Again, we don't have information on the disaggregated numbers of incident cases from R based on how they entered R, but we do have information on the entry to R.

$$\begin{aligned}\text{Total number entering into R per year} = \;&\text{Total number entering R from T per year} \\ &+ \text{Total number entering R from Ds per year} \\ &+ \text{Total number entering R from Dc per year}\end{aligned}$$

The proportion of the total flow into R from each of T, Ds, and Dc per year is:

$$\frac{\text{TR}}{(\text{TR+ DsR + DcR})} \qquad \frac{\text{DsR}}{(\text{TR+ DsR + DcR})} \qquad \frac{\text{DcR}}{(\text{TR+ DsR + DcR})}$$

*TR = flow from T to R = total number entering R from T* per year
*DcR = flow from Dc to R = total number entering R from Dc* per year
*DsR = flow from Ds to R = total number entering R from Ds* per year
*TR + DsR + DcR = total number entering R* per year

If we assume that the flows **INTO** R from each of T, Ds, and Dc are in the same proportions as the flows **OUT** of R, then we can disaggregate the outflow from R (which is the number of incident cases of people from R, per year, we called RD in the equation above) into incident cases of people from R who entered R from each of T, Ds, and Dc, per year by multiplying RD by the proportion from each of T, Ds, and Dc

Number of incident cases of people from R who entered R from T, per year
= (Number of incident cases of people from R, per year)(Proportion of flow out of R that is from people who entered R from T, per year) etc.

We can rewrite RD in terms of the disaggregated pathways from T, Ds, and Dc:

$$\mathrm{RD} = \mathrm{RD}\times\frac{\mathrm{TR}}{(\mathrm{TR}+\mathrm{DsR}+\mathrm{DcR})} + \mathrm{RD}\times\frac{\mathrm{DsR}}{(\mathrm{TR}+\mathrm{DsR}+\mathrm{DcR})} + \mathrm{RD}\times\frac{\mathrm{DcR}}{(\mathrm{TR}+\mathrm{DsR}+\mathrm{DcR})}$$

Subbing in the expression for RD above into the equation for the proportion of incident cases from R, we obtain:

$$\frac{\mathrm{RD}\times\frac{\mathrm{TR}}{(\mathrm{TR}+\mathrm{DsR}+\mathrm{DcR})} + \mathrm{RD}\times\frac{\mathrm{DsR}}{(\mathrm{TR}+\mathrm{DsR}+\mathrm{DcR})} + \mathrm{RD}\times\frac{\mathrm{DcR}}{(\mathrm{TR}+\mathrm{DsR}+\mathrm{DcR})}}{(\mathrm{RD}+\mathrm{LsD}+\mathrm{LfD})}$$

Factor, simplify and rewrite:

$$= \frac{\mathrm{RD}}{(\mathrm{RD}+\mathrm{LsD}+\mathrm{LfD})} * \left(\frac{\mathrm{TR}}{(\mathrm{TR}+\mathrm{DsR}+\mathrm{DcR})} + \frac{\mathrm{DsR}}{(\mathrm{TR}+\mathrm{DsR}+\mathrm{DcR})} + \frac{\mathrm{DcR}}{(\mathrm{TR}+\mathrm{DsR}+\mathrm{DcR})}\right)$$

$$= \left[\frac{\mathrm{RD}}{(\mathrm{RD}+\mathrm{LsD}+\mathrm{LfD})}\right]\left[\frac{\mathrm{TR}}{(\mathrm{TR}+\mathrm{DsR}+\mathrm{DcR})}\right] + \left[\frac{\mathrm{RD}}{(\mathrm{RD}+\mathrm{LsD}+\mathrm{LfD})}\right]\left[\frac{\mathrm{DsR}}{(\mathrm{TR}+\mathrm{DsR}+\mathrm{DcR})}\right] + \left[\frac{\mathrm{RD}}{(\mathrm{RD}+\mathrm{LsD}+\mathrm{LfD})}\right]\left[\frac{\mathrm{DcR}}{(\mathrm{TR}+\mathrm{DsR}+\mathrm{DcR})}\right]$$

Proportion of incident cases from R =
(Proportion of incident cases from R who entered R from T)
+ (Proportion of incident cases from R who entered R from Dc)
+ (Proportion of incident cases from R who entered R from Ds)

The assumption we make here is that (at equilibrium) these proportions of incident cases will be equivalent for flows entering Ds (incident cases), entering Dc (progression from subclinical to clinical disease) and entering T (treatment initiation / case notifications).

$$\frac{\mathrm{RD}}{(\mathrm{RD}+\mathrm{LsD}+\mathrm{LfD})} \qquad \frac{\mathrm{LfD}}{(\mathrm{RD}+\mathrm{LsD}+\mathrm{LfD})} \qquad \frac{\mathrm{LsD}}{(\mathrm{RD}+\mathrm{LsD}+\mathrm{LfD})}$$

$$\left[\frac{\mathrm{RD}}{(\mathrm{RD}+\mathrm{LsD}+\mathrm{LfD})}\right]\left[\frac{\mathrm{TR}}{(\mathrm{TR}+\mathrm{DsR}+\mathrm{DcR})}\right] \quad \left[\frac{\mathrm{RD}}{(\mathrm{RD}+\mathrm{LsD}+\mathrm{LfD})}\right]\left[\frac{\mathrm{DsR}}{(\mathrm{TR}+\mathrm{DsR}+\mathrm{DcR})}\right]$$

$$\left[\frac{\mathrm{RD}}{(\mathrm{RD}+\mathrm{LsD}+\mathrm{LfD})}\right]\left[\frac{\mathrm{DcR}}{(\mathrm{TR}+\mathrm{DsR}+\mathrm{DcR})}\right]$$

Therefore, the proportion of notifications of people who were in R having entered R from T will be the same as the proportion of incidence from people who were in R having entered R from T.

Going back to the calibration target once again:

$$= \frac{\text{Number of notifications of people who were in R having entered R from T per year}}{\text{Total number of notifications per year}}$$

Although we do not know the number of notifications of people who were in R having entered R from T per year, this is equal to the proportion of notifications of people who were in R having entered R from T multiplied by the total number of notifications per year

$$= \frac{\text{Total number of notifications per year}\times\text{Proportion of notifications of people who were in R having entered R from T}}{\text{Total number of notifications per year}}$$

We can cancel out the total number of notifications as it is in both the numerator and denominator.

$$= \frac{\text{Total number of notifications per year} \times \text{Proportion of notifications of people who were in R having entered R from T}}{\text{Total number of notifications per year}}$$

$$= \text{Proportion of notifications of people who were in R having entered R from T}$$

This value is calculated as the proportion of notifications of people who were in R multiplied by the proportion of the entry into R that came from T

$$= [\frac{\text{RD}}{(\text{RD + LsD + LfD})}][\frac{\text{TR}}{(\text{TR + DcR + DsR})}]$$

However, there may be some people who recently entered R from Dc or Ds, but who had also previously had treatment. Therefore, the previous equation is revised as:

$$= [\frac{\text{RD}}{(\text{RD + LsD + LfD})}][\frac{\text{TR + (DcR + DsR)} \times \text{(proportion of those in (DcR + DsR) who have been treated previously}}{(\text{TR + DcR + DsR})}]$$

We assume that the proportion of those in (DcR + DsR) who have been treated previously is the same as the proportion of those in DsR who have been treated previously. We can then set the value:

$$\text{proportion of those in (DcR + DsR) who have been treated previously} =$$

$$[\frac{\text{RD}}{(\text{RD + LsD + LfD})}][\frac{\text{TR + (DcR + DsR)} \times \text{(proportion of those in (DcR + DsR) who have been treated previously}}{(\text{TR + DcR + DsR})}]$$

If we substitute in the term, we can see that the same term is repeated again and again. Let

$$\text{A} = \frac{\text{RD}}{(\text{RD + LsD + LfD})(\text{TR + DcR + DsR})}$$

Then we can rewrite the above as:

$$\text{A[TR + (Ds + Dc)[A[TR + (Ds+Dc)[A[...]]]}$$

Let $\text{B} = \text{TR}$ and $\text{C} = (\text{Ds + Dc})$, substitute and expand:

$$\text{A[B + C(A[B + C(A[...]))}$$

$$\text{AB + AC(AB + AC(AB[...]))}$$

Let $\text{X} = \text{AB}$ and $\text{Y} = \text{AC}$, substitute and expand:

$$= \text{X + Y(X + Y(X[...]))}$$

$$= \text{X} + \text{XY} + \text{XY}^2 + \text{XY}^3 ...$$

$$= \text{X} (1 + \text{Y} + \text{Y}^2 + \text{Y}^3 ...)$$

$$= \frac{X}{(1-Y)}$$

Substituting back in for $X, Y, A, B$ , and$C$ we obtain:

$$= \frac{\text{AB}}{\text{(1-AC)}}$$

$$= \frac{\frac{\text{RD}}{(\text{RD + LsD + LfD})(\text{TR + DcR + DsR})}\times \text{TR}}{(1-\frac{\text{RD}}{(\text{RD + LsD + LfD})(\text{TR + DcR + DsR})}\times(\text{DcR + DsR}))}$$

$$= \frac{\text{RD}\times\text{TR}}{(\text{RD + LsD + LfD})(\text{TR + DcR + DsR}) \text{ - } \text{RD}(\text{DcR + DsR})}$$

$$= \frac{\text{RD}\times\text{TR}}{\text{TR}(\text{RD + LsD + LfD}) + (\text{DcR + DsR})(\text{LsD + LfD})}$$

*RD = flow from R to Ds = total number of incident cases of people from R, per year*
*TR = flow from T to R = total number entering R from T, per year*
*RD + LsD + LfD = total number of incident cases, per year*

*DcR = flow from Dc to R = total number entering R from Dc, per year*
*DsR = flow from Ds to R = total number entering R from Ds, per year*
*LsD = flow from Ls to Ds = total number of incident cases of people from Ls, per year*
*LfD = flow from Lf to Ds = total number of incident cases of people from Lf, per year*

#### 4. Policy scenarios

##### 4.1 No-new-vaccine baseline

The primary no-new-vaccine simulated was the no-new-vaccine baseline, which assumed non-vaccine tuberculosis interventions continue at current levels into the future. As reported country-level data includes the high coverage levels of neonatal BCG vaccination, this was not explicitly modelled. We assumed that BCG vaccination would not be discontinued over the model time horizon.

##### 4.2 Vaccine delivery scenarios

Two recently completed phase 2 trials have demonstrated encouraging efficacy results. The M72/AS01$_E$ candidate vaccine is a subunit vaccine for which results from a completed Phase IIb trial were published at the end of 2019.[36] After three years of follow-up, the efficacy of M72/AS01$_E$ at preventing disease in latently infected adults from South Africa, Zambia, and Kenya was estimated at 49.7% (95% confidence interval = 2.1–74.2).[36] To confirm this finding, a larger, Phase III follow-up study is needed, which includes participants who are uninfected, adolescents, as well as those living with HIV to assess safety and immunogenicity in these populations. This is being planned.

BCG-revaccination (administering a second dose of BCG to those who were vaccinated neonatally) was previously implemented in many countries, however evidence did not support the effectiveness of this practice. Interest in BCG-revaccination has recently been renewed following results from a trial for the vaccine candidate, H4:IC31. BCG-revaccination was assessed as a third parallel arm alongside H4:IC31 and a placebo in a pre-infection population in South Africa, and although neither vaccine appeared efficacious at preventing infection, BCG-revaccination appeared efficacious at preventing sustained infection (defined as three consecutive positive tests after day 84 of the trial) with an efficacy of 45.4% (6.4–68.1).[37] A larger trial of BCG-revaccination versus placebo in 1800 healthy adolescents from across South Africa is now underway to verify this finding.

We evaluated introducing vaccines with M72/AS01$_E$ and BCG-revaccination characteristics compared to the no-new-vaccine baseline as described in the subsequent sections.

###### 4.2.1 Classifying tuberculosis vaccines

Before describing the specific characteristics for the vaccine scenarios that we investigated, we provide a brief overview on classifying tuberculosis vaccines (descriptions from Clark et al.[1]).

Tuberculosis vaccines are characterised on four key characteristics: the vaccine efficacy, the host infection status at the time of vaccination required for the vaccine to be efficacious, the mechanism of effect, and the duration of protection. Vaccine efficacy defines the magnitude of protection induced by the vaccine. Vaccine efficacy is assumed to be either “all or nothing”, where the vaccine offers full protection to a subset of individuals (equal to the vaccine efficacy) who were vaccinated, or “degree”, where the vaccine offers partial protection to all individuals who received the vaccine.

The host infection status at the time of vaccination required for the vaccine to be efficacious defines the *Mtb* infection status required of the population at the time they receive the vaccine for the vaccine to be efficacious. We divide the host infection status into No Current Infection (NCI), where the vaccine is efficacious in uninfected populations only, Current Infection (CI), meaning the vaccine is efficacious in populations with current infection with *Mtb* only, or Any Infection (AI) where the vaccine is efficacious in both pre- and post-infection populations.

The vaccine mechanism of effect type determines how the vaccine will offer protection. A prevention of infection (POI) vaccine protects individuals from initial or re-infection with *Mtb*, whereas a prevention of disease (POD) vaccine functions by preventing individuals who may be uninfected or infected with *Mtb* from progressing to active disease. A prevention of infection and disease vaccine (POI&D) prevents both infection and disease. Finally, the duration of protection represents the length of time following vaccination that individuals are protected.

##### 4.2.2 M72/AS01$_E$ and BCG-revaccination scenarios

For each vaccine product, we established one "Basecase" vaccine scenario based on clinical trial data and expert opinion. We then varied vaccine product and delivery scenarios as univariate scenario analyses from the Basecase scenario as described in Table S4.1.

**Table S4.1 M72/AS01$_E$ and BCG-revaccination scenarios evaluated in the analysis**

| Characteristic | M72/AS01$_E$ | | BCG-revaccination | |
|---|---|---|---|---|
| | **Basecase** | **Varied in univariate** | **Characteristic** | **Basecase** |
| **Policy scenarios** | | | | |
| Age targeting | Campaign for ages 16-34, routine age 15 | Older ages (campaign for ages 18-55)<br><br>Elderly ages (campaign for ages 61+, routine age 60) | Campaign for ages 11-18, routine age 10 | Older ages (campaign for ages 16-34, routine age 15)<br><br>Elderly ages (campaign for ages 61+, routine age 60) |
| **Vaccine characteristic and coverage scenarios** | | | | |
| Vaccine efficacy | 50% | 60%<br>70% | 45% | 70% |
| Duration of protection | 10 years | 5 years<br>15 years<br>20 years | 10 years | 5 years<br>15 years<br>20 years |
| Host infection status | AI | CI | NCI | AI |
| Mechanism of effect | Prevention of disease | Prevention of infection and disease | Prevention of infection | Prevention of infection and disease |
| Introduction year (years of any repeat campaigns) | 2030<br>(2040) | 2036<br>(2046) | 2025<br>(2035, 2045) | 2031<br>(2041) |
| Achieved vaccine coverage | Campaign = 70% / Routine = 80% | Campaign = 50% / Routine = 70%<br><br>Campaign = 90% / Routine = 90% | 80% | 70%<br>90% |

##### 4.2.3 Vaccine delivery assumptions

***Vaccine eligible population***

In our modelling, we assume that there is no pre-vaccination infection testing. Therefore, even if a vaccine is only effective when delivered to uninfected individuals at the time of vaccination, we assume that both uninfected and infected individuals will receive the vaccine, and only the uninfected individuals will receive protection. Our model structure allows for counting and tracking individuals who received the vaccine but do not receive any protection from it.

***Efficacy***
From trial data, the efficacy of M72/AS01E at preventing disease in latently infected adults was estimated at 49.7% (2.1–74.2).[36] Therefore, our Basecase vaccine efficacy was set at 50%, and based on expert opinion we evaluated 60% and 70% as scenario analyses. BCG-revaccination appeared efficacious at preventing *sustained* infection with an efficacy of 45.4% (6.4–68.1).[37] The Basecase efficacy was set to 45%, and 70% was evaluated in a scenario analysis.

***Protection from repeat vaccinations***
In the event that an individual who is currently protected with a vaccine receives another course, after consultation with an immunologist we have made some assumptions on the resulting level of vaccine protection:

BCG-revaccination: Based on expert advice, we assume that no additional protection is afforded if a second or third vaccine is administered while the individual is currently protected from the first.

M72/AS01E: Based on expert advice, we assume that overall vaccine protection increases if a second vaccine is administered while the individual is currently protected by a first vaccine. We assume that this protection increases by (1-current protection) times vaccine efficacy, as in Table S4.2.

**Table S4.2 Increase in protection for the number of vaccine courses**

| Number of vaccine courses currently protected by | Basecase | Efficacy variation 1 | Efficacy variation 2 |
|---|---|---|---|
| One | 50.0% | 60.0% | 70.0% |
| Two | 75.0% | 84.0% | 91.0% |

Note that the number of vaccine courses refers to the number of vaccine courses that the individual is *currently* protected by, not that they have ever a) received, or b) been protected by. For example, if someone receives one vaccine, then wanes, then receives another one, they would only be currently protected by one, <u>not two</u>, vaccines, and so the efficacy would be either 50%, 60%, or 70% depending on the scenario.

***Mechanism of effect***
We assume that a vaccine that protects against infection will work by reducing the rate of infection for both initial and re-infection, and that a vaccine that protects against progression to disease will work by reducing the rate of progression to subclinical disease. If the vaccine protects against both infection and disease we assume that it has the same efficacy against preventing disease as it does infection. For example, if the vaccine is defined as a prevention of infection and disease vaccine with 50% efficacy, it reduces the rate of infection by 50% and the rate of progression to disease by 50%.

***Introduction year***
The Basecase introduction years, 2025 and 2030 for BCG-revaccination and M72/AS01E respectively, were determined based on considering when new trial data would become available, as well as incorporating time for licensure and policy change. The introduction year considered in scenario analyses, 2031 and 2036 for BCG-revaccination and M72/AS01E respectively, was based on applying IAVI/Full Value Assessment of Tuberculosis Vaccines analyses from Shelly Malhotra and expert advice to the earliest possible introduction year.[1]

***Age targeting***
The Basecase age was informed by ages of trial participants and expert advice. Additional scenarios were informed by work conducted by Pelzer et. al and expert advice.[38]

#### 4.3 Vaccine model structure

Depending on the host infection status required at the time of vaccination for the vaccine to be efficacious, we implemented a different vaccine structure in the model to account for differences in Vaccinated Protected, Vaccinated Not Protected, and Vaccinated Waned. Each compartment in the vaccine structure is replicated for all tuberculosis natural history compartments, access-to-care strata, and ages.

##### 4.3.1 No Current Infection vaccines

A No Current Infection (NCI) vaccine requires an individual to be uninfected at the time of vaccination in order for the vaccine to be efficacious. Implementation in the TBVax model of an NCI vaccine with the possibility of two repeat vaccine courses is provided in Figure S4.1. For our purposes, we assume that the level of protection remains the same regardless of the number of vaccine courses received (i.e. level of protection in "Vaccinated Protected (one vaccine course)" is equal to "Vaccinated Protected (two vaccine courses)" etc.). Additionally, because the vaccine is only efficacious for NCI, and in this model once you leave $U_N$ (the state where the vaccine is effective) you never return, once you enter a "Vaccinated Not Protected" state you *never* have the opportunity to become "Vaccinated Protected" again.

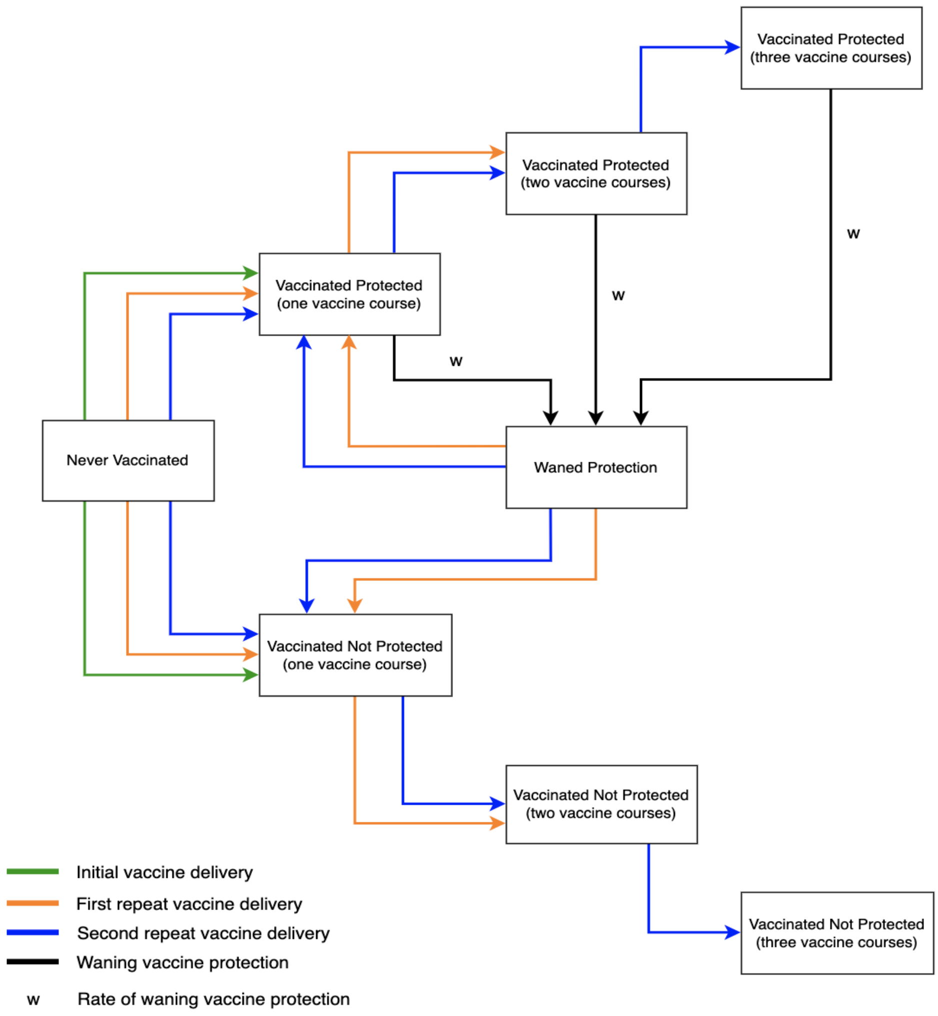


**Figure S4.1 Vaccine structure for a NCI vaccine**

##### 4.3.2 Current Infection vaccines

A Current Infection (CI) vaccine requires an individual to be infected at the time of vaccination in order for the vaccine to be efficacious. Implementation in the TBVax model of an CI vaccine with the possibility of two repeat vaccine courses is provided in Figure S4.2. For our purposes, we assume that the level of protection builds with each vaccine course, with efficacy values as in Table S4.2.

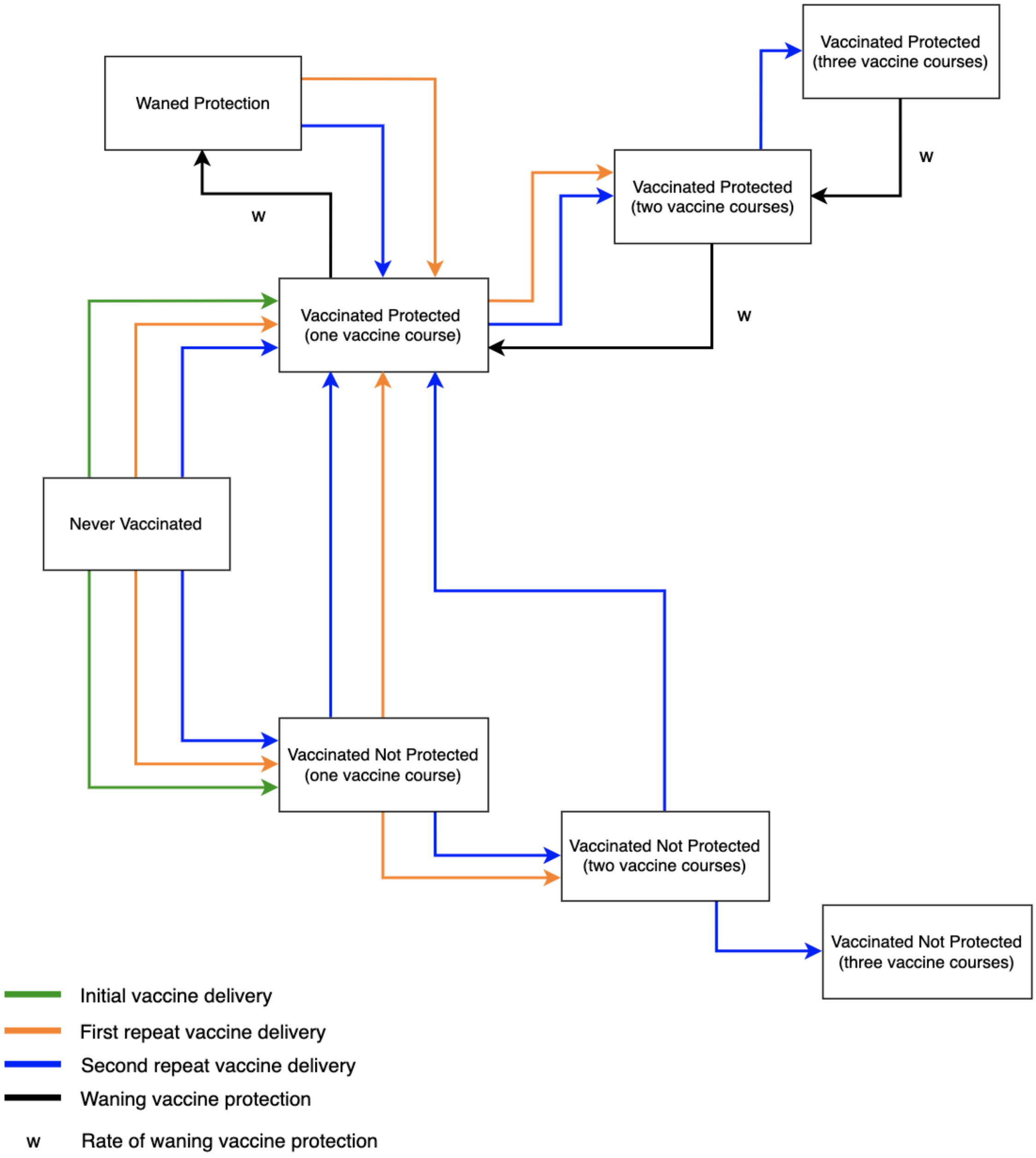


**Figure S4.2 Vaccine structure for a CI vaccine (where protection builds with each vaccine course)**

##### 4.3.3 Any Infection vaccines

An Any Current Infection (AI) vaccine will be efficacious with any infection status (aside from current active disease) at the time of vaccination. The "Vaccine Not Protected" compartments remain as we assume that individuals with subclinical disease may be accidentally vaccinated and would not receive protection from the vaccine. However, we do want to keep track of the number of vaccinations for cost purposes.

***AI-1 vaccines:*** With each vaccine course the level of protection remains the same (Figure S4.3). Waning occurs from any of the Vaccinated Protected compartments to the Waned Protection compartment.

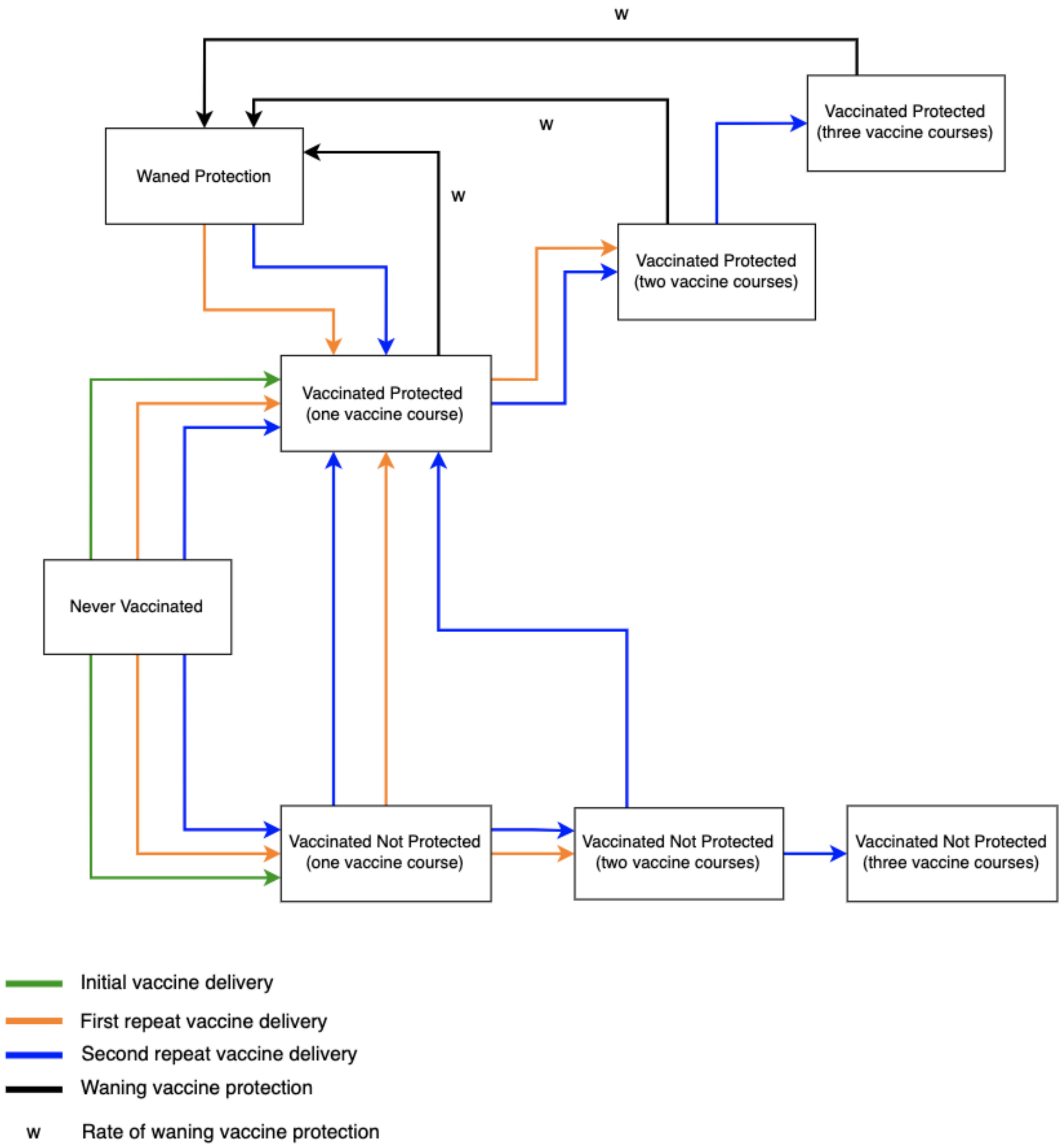


**Figure S4.3 Vaccine structure for an AI vaccine (where protection does not build with each vaccine course)**

***AI-2 vaccines:*** With each vaccine course the level of protection builds if the recipient is currently in a Vaccinated Protected compartment (Figure S4.4). This is the same structure as the CI vaccine with protection building (Figure S4.2). Waning occurs from any of the Vaccinated Protected compartments to the Vaccinated Protected compartment one level below, or to the Waned Protection compartment for those with only one course of protection.

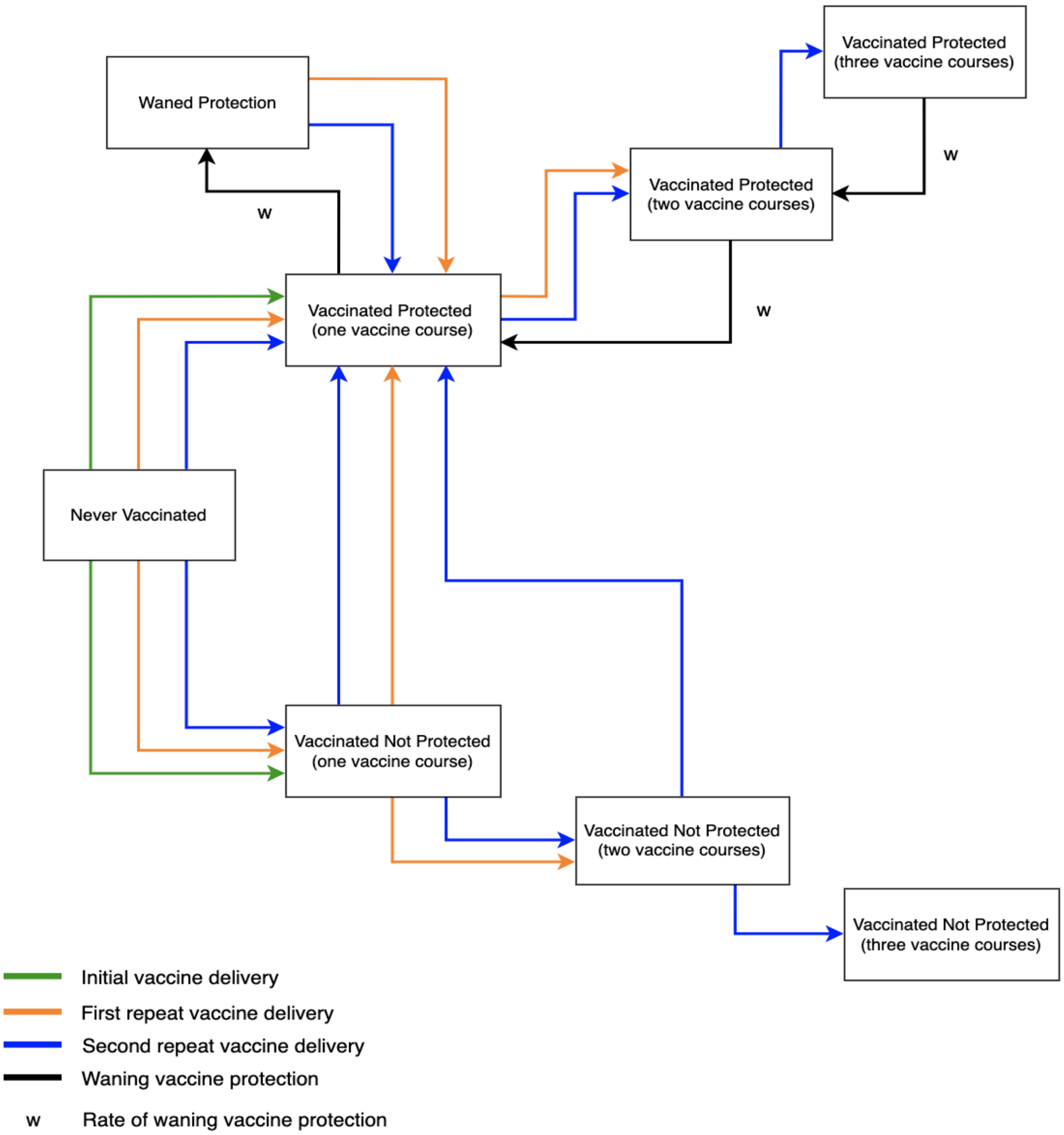


**Figure S4.4 Vaccine structure for an AI vaccine (where protection builds with each vaccine course)**

#### 5. Economic analysis methods

Before undertaking this work, we established an economic analysis plan, involving stakeholders and government officials to ensure we had incorporated all necessary information and planned to report on all key outcomes, to outline the methods used in this work. This is summarised below.

##### 5.1 Calculation of disability-adjusted life years

We calculated the difference in total disability-adjusted life years (DALYs) from vaccine introduction to 2050 for each scenario compared to the no-new-vaccine baseline. We used the disability weight for tuberculosis disease from the Global Burden of Disease 2019 study,[39] and country- and age-specific life expectancy estimates from the United Nations Development Programme.[40] To incorporate parameter uncertainty in years lost due to disability (YLD) weight estimates, we made 1000 draws from disability weight uncertainty ranges.

##### 5.2 Tuberculosis-related cost model

We estimated health system unit costs, patient costs and productivity losses based on a scoping review of published literature. For the tuberculosis programme, we obtained unit costs for drug-susceptible (DS) and drug-resistant (DR) tuberculosis treatment and diagnostic costs. Uncertainty in cost estimates is characterised through gamma distributions around plausible unit cost estimates in a probabilistic sensitivity analysis. There was considerable uncertainty in the cost of delivering a vaccine, including the price of vaccine compounds and programmatic delivery among adolescents. Based on expert opinion from funders, for the M72/AS01$_E$ vaccine we assume a \$2.50 per-dose vaccination price with two doses per course assumed in the Basecase. Based on the average estimated BCG price from 2020–2023 from UNICEF,[43] the vaccine price per dose for BCG-revaccination was set at \$0.17, with one dose assumed per course.

##### 5.3 Vaccine introduction

All cost inputs are given in Table S5.1.

Due to uncertainty in unit costs of vaccine supply and introduction among populations who may not typically receive large-scale mass vaccination, we make several assumptions around costs to supply and introduction of vaccines. Uncertainty in cost estimates is characterised through gamma distributions.

One-time vaccine introduction costs are included in years where there is a campaign and represent non-recurring costs such as establishing infrastructure and providing training for healthcare professionals. The costs were assumed to be \$2.40 (1.20–4.80) per individual in the targeted age group (as opposed to the actual number of recipients) based on the vaccine introduction support policy of Gavi, the Vaccine Alliance.[44] Vaccine delivery was assumed to be \$2.50 (1.00–5.00) per dose, with a further \$0.11 (0.06–0.22) supply costs per dose.[45] The cost of recipient vaccination time was \$0.94 (0.13–1.52), which was calculated by multiplying a wage proxy of GDP per capita for India by an estimate of the time required for vaccination.[46,47] We assume a 5% wastage rate.

For each year in the five-year scale up, the vaccination cost is calculated as:
Vaccination cost = (one time introduction costs) × (targeted age group population size) × 0.2 + (number of people vaccinated) × (number of doses) × (vaccine price + vaccine supply costs + cost of delivery) × (1 + wastage)

For each year where there is a repeat campaign, the vaccination cost is calculated as:
Vaccination cost = (one time introduction costs) × (targeted age group population size) + (number of people vaccinated) × (number of doses) × (vaccine price + vaccine supply costs + cost of delivery) × (1 + wastage)

For each year where there is only routine delivery of the vaccine, the vaccination cost is calculated as:
Vaccination cost = (number of people vaccinated) × (number of doses) × (vaccine price + vaccine supply costs + cost of delivery) × (1 + wastage)

For the vaccination cost from the societal perspective, the patient time cost of vaccination was added as a multiplier to the number of doses, and therefore included in the equation along with vaccine price, vaccine supply costs, and the cost of delivery.

#### 5.4 Cost-effectiveness analysis and willingness-to-pay thresholds

We calculated the incremental cost effectiveness ratio as the ratio between the incremental benefit, in DALYs averted, and the incremental cost, in USD, for each run across vaccination and baseline scenario. Both costs and benefits were discounted to 2025 (when vaccination began) at 3% per year, per guidelines.[48] We measured cost-effectiveness by 2050 against three India specific cost thresholds: 1x gross domestic product (GDP) per-capita (US$1,927.71),[47] and two country-level opportunity cost thresholds defined by Ochalek et al [the upper (US$363), and lower (US$264) bounds].[49]

#### 5.5 Total costs from the health-system and societal perspectives

The following costs are included in the health-system perspective:

- Vaccine costs: One-time vaccine introduction costs, recurring vaccine delivery costs, vaccine price per dose, and supply costs
- Cost of testing and diagnosis for drug-susceptible and drug-resistant cases
- Cost of treatment for drug-susceptible and drug-resistant cases

In addition to the costs from the health-system perspective, costs from the societal perspective include:

- Vaccine costs: Patient time cost for vaccination
- Non-medical patient costs (including transportation) for drug-susceptible and drug-resistant cases
- Indirect patient costs for drug-susceptible and drug-resistant cases

**Table S5.1 Tuberculosis testing, diagnostic, and vaccination related cost inputs**

| Unit Cost | Estimate | Lower Bound | Upper Bound | Sources |
|---|---|---|---|---|
| Unit cost of testing/diagnosis for DS cases per person | $22.45 | $18.37 | $26.53 | 50 |
| Unit cost of testing/diagnosis for DR cases per person | $24.36 | $5.04 | $117.81 | 51 |
| Unit cost of treatment for DS cases per person | $317.00 | $254.00 | $374.00 | 52 |
| Unit cost of treatment for DR cases per person | $3,891.00 | $3,382.00 | $4,401.00 | 53 |
| Non-medical patient cost per DS-TB disease episode (including transportation) per person | $51.25 | $22.12 | $76.94 | 54,55 |
| Indirect patient cost per DS-TB disease episode (time spent on treatment and transport * wage) per person | $117.01 | $24.04 | $460.24 | 55,56 |
| Non-medical patient cost per DR-TB disease episode (including transportation) per person | $143.49 | $61.95 | $215.42 | 54,55 |
| Indirect patient cost per DR TB disease episode (time spent on treatment and transport * wage) per person | $327.63 | $67.30 | $1,288.66 | 55,56 |
| Recurrent vaccine delivery cost per person per dose | $2.50 | $1.00 | $5.00 | 44 |
| One-time vaccine introduction costs per targeted person | $2.40 | $1.20 | $4.80 | 44 |
| Vaccine supply costs per person per dose | $0.11 | $0.06 | $0.22 | 45 |
| Cost of vaccination time per person per dose | $0.94 | $0.13 | $1.52 | 46,47 |

#### 6. Health impact outcomes

The following measures were calculated for each vaccine scenario as the median and 95% uncertainty range

- Percent incidence rate reduction in 2050 for each vaccine scenario compared to the estimated value in 2050 by *No-New-Vaccine* baseline
- Percent mortality rate reduction in 2050 for each vaccine scenario compared to the estimated value in 2050 by *No-New-Vaccine* baseline
- Cumulative cases averted for each vaccine scenario between vaccine introduction (either 2025 or 2030) and 2050 compared to the cumulative number of cases estimated by the *No-New-Vaccine* baseline between the corresponding years
- Cumulative deaths averted for each vaccine scenario between vaccine introduction (either 2025 or 2030) and 2050 compared to the cumulative number of cases estimated by the *No-New-Vaccine* baseline between the corresponding years
- Cumulative treatments averted for each vaccine scenario between vaccine introduction (either 2025 or 2030) and 2050 compared to the cumulative number of cases estimated by the *No-New-Vaccine* baseline between the corresponding years

**SUPPLEMENTARY RESULTS**

#### 7. No-new-vaccine baselines

##### 7.1 *Status Quo* no-new-vaccine baseline calibration

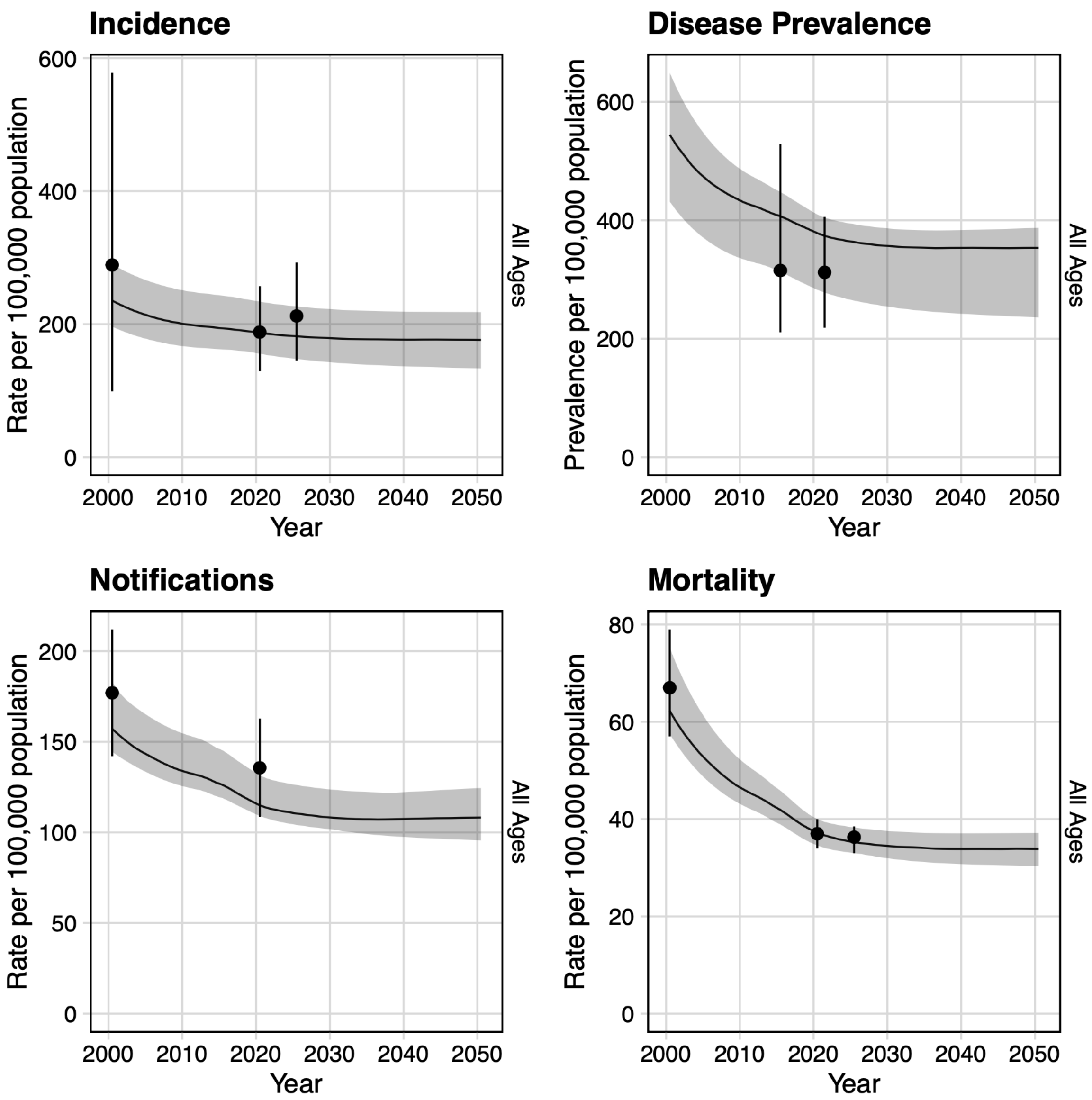


**Figure S7.1 Tuberculosis incidence, disease prevalence, case notification and mortality rate trends from 2000–2050 for all ages**

*The black trend line indicates the median modelled output with 95% uncertainty in shaded grey. The black dot and vertical line is the calibration target from Table S3.1.*

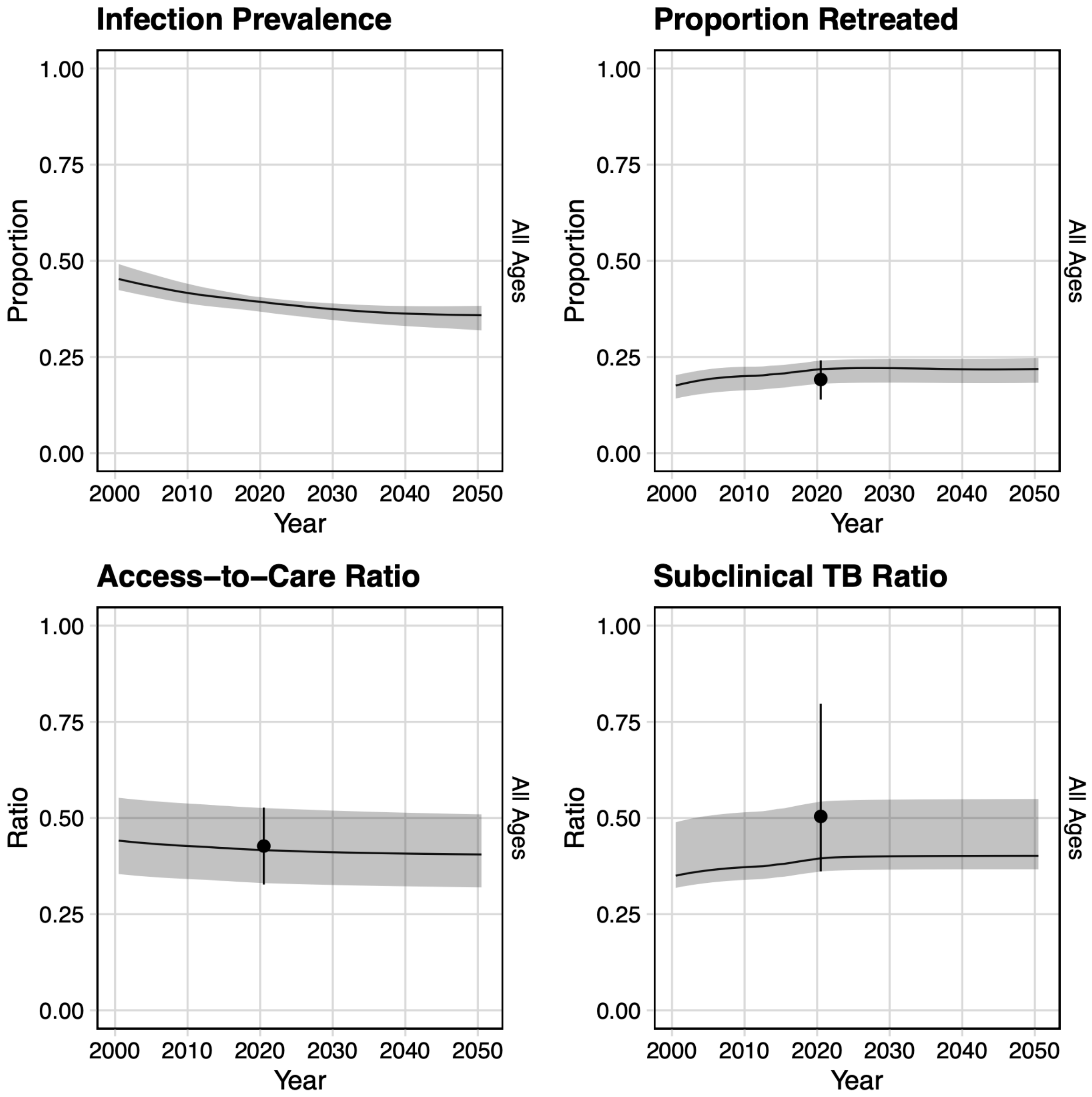


**Figure S7.2 Tuberculosis infection prevalence, proportion retreated, access-to-care ratio and ratio of subclinical tuberculosis to total tuberculosis trends from 2000–2050 for all ages**

*The black trend line indicates the median modelled output with 95% uncertainty in shaded grey. The black dot and vertical line is the calibration target from Table S3.1.*

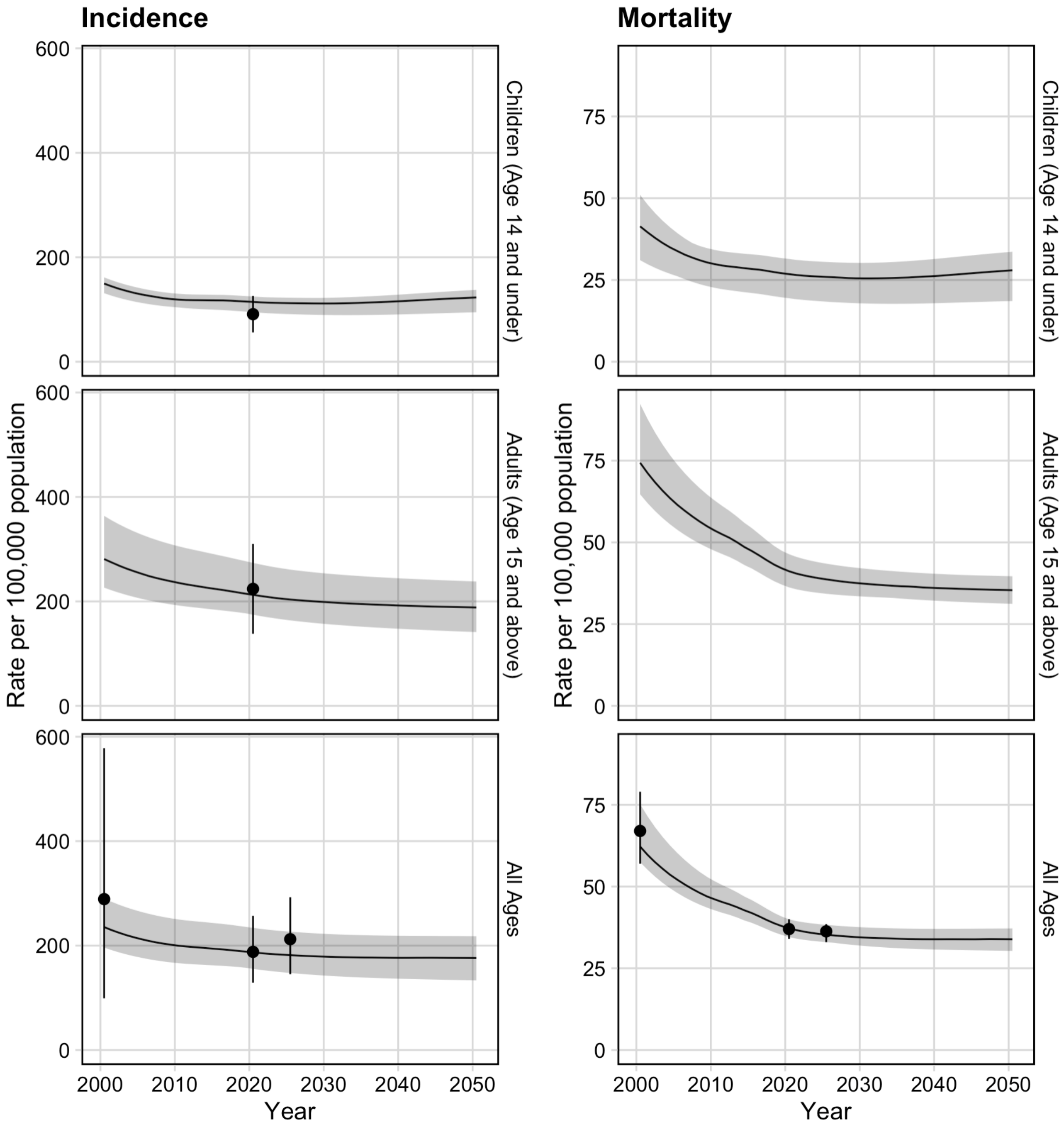


**Figure S7.3 Tuberculosis incidence and mortality rate trends from 2000–2050 by age group**

*The black trend line indicates the median modelled output with 95% uncertainty in shaded grey. The black dot and vertical line is the calibration target from Table S3.1.*

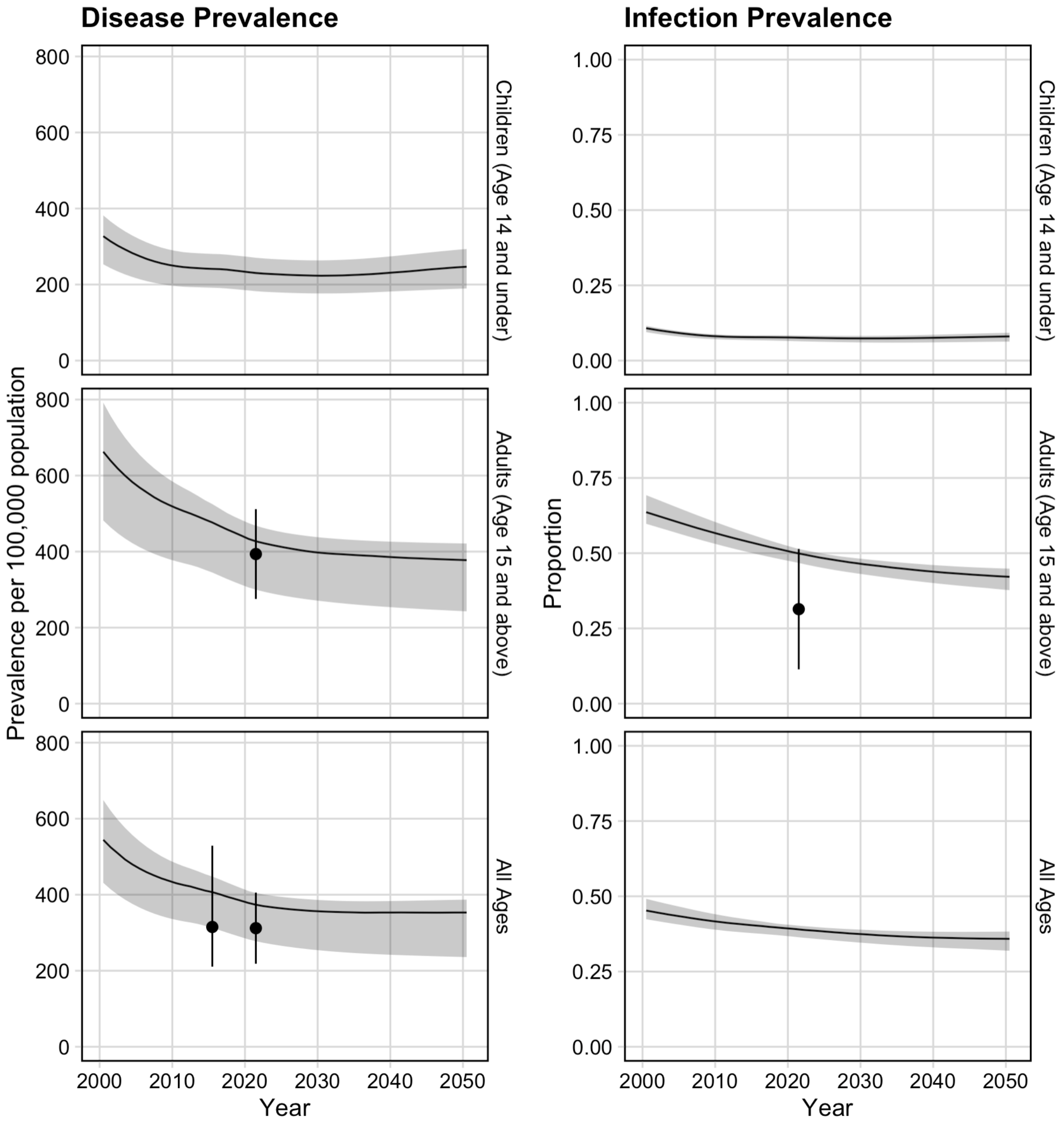


**Figure S7.4 Tuberculosis disease and infection prevalence trends from 2000–2050 by age group**

*The black trend line indicates the median modelled output with 95% uncertainty in shaded grey. The black dot and vertical line is the calibration target from Table S3.1.*

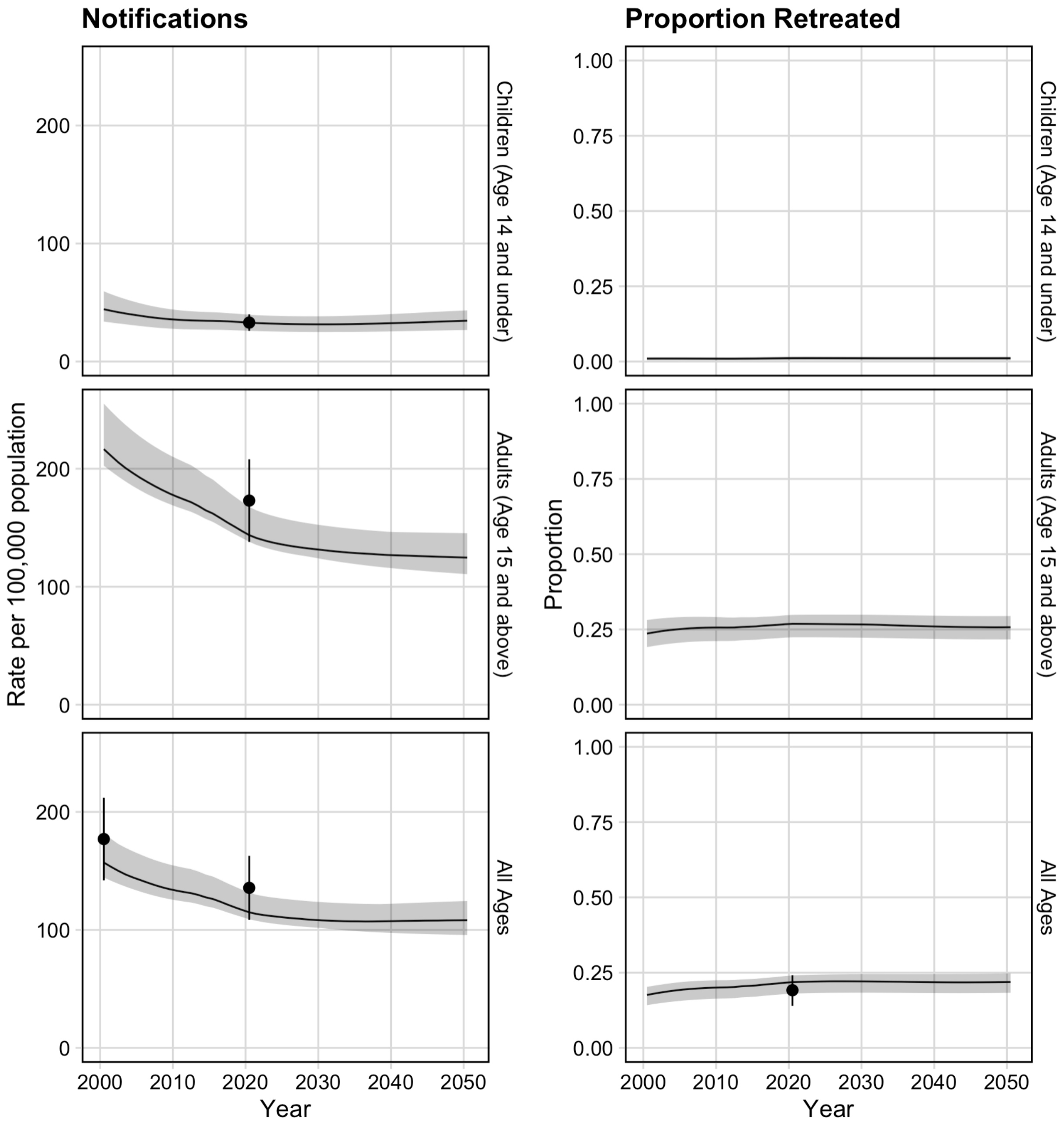


**Figure S7.5 Tuberculosis case notification and proportion retreated trends from 2000–2050 by age group**

*The black trend line indicates the median modelled output with 95% uncertainty in shaded grey. The black dot and vertical line is the calibration target from Table S3.1.*

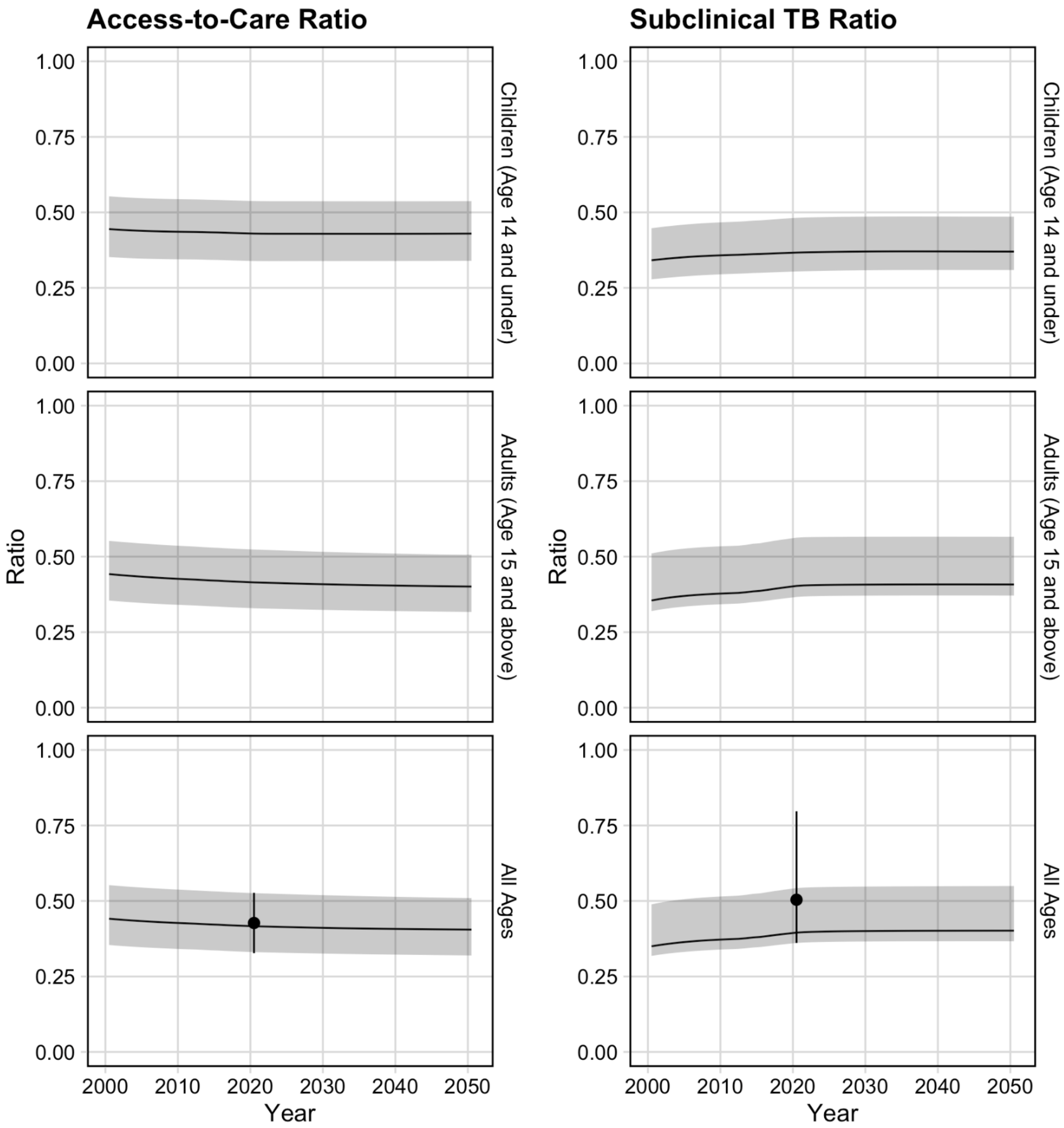


**Figure S7.6** **Access-to-care ratio and the ratio of subclinical tuberculosis to all active tuberculosis trends from 2000–2050 by age group**

*The black trend line indicates the median modelled output with 95% uncertainty in shaded grey. The black dot and vertical line is the calibration target from Table S3.1.*

#### 7.2 Posteriors distributions for model parameters

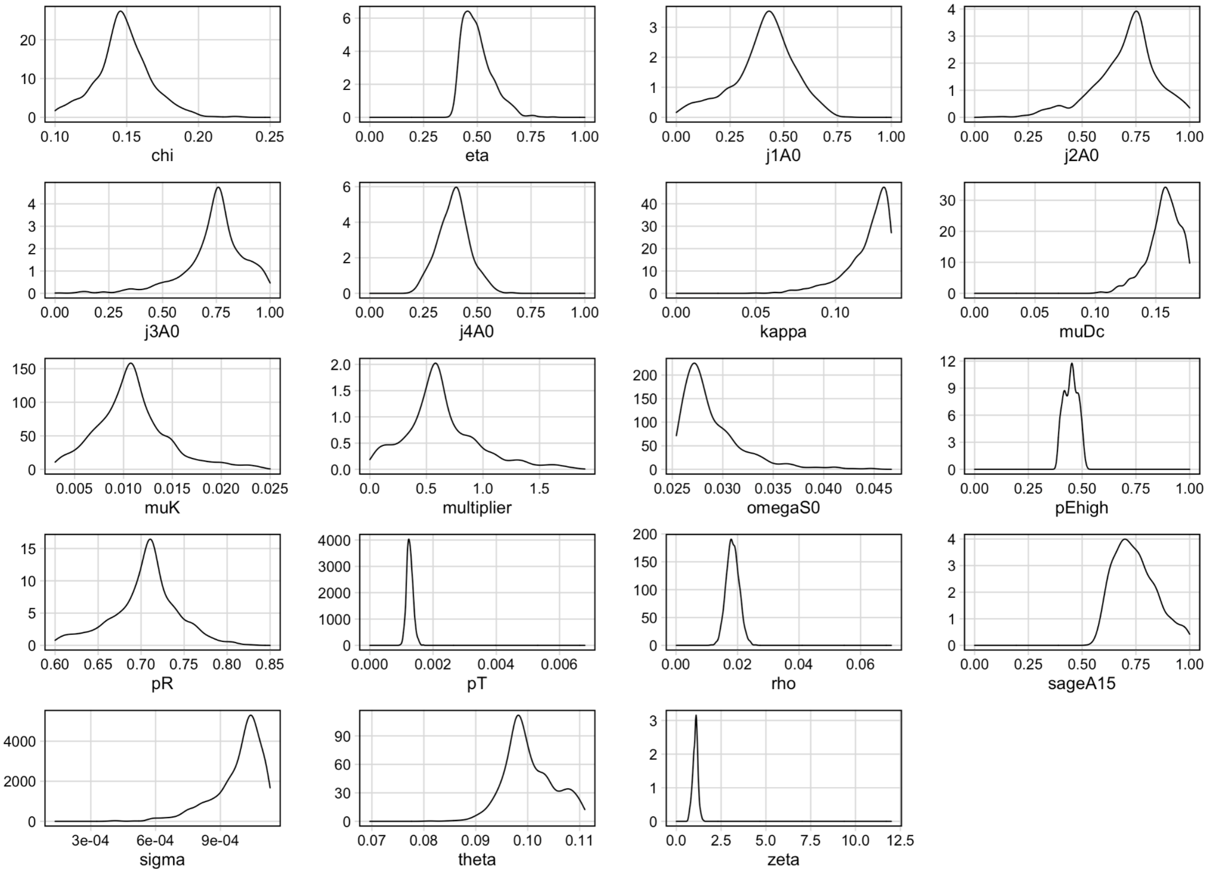


**Figure S7.7 Posterior distributions for the 1000 parameter sets of the 19 parameters varied during calibration**

*Definitions: chi = rate of natural cure, eta = rate of treatment initiation, j1A0 = age multiplier for rate of fast progression (theta), j2A0 = age multiplier for rate of reactivation (sigma), j3A0 = age multiplier for rate of relapse (rho), j4A0 = age multiplier for rate of treatment initiation, kappa = on-treatment mortality fraction, muDc = rate of clinical disease mortality, muK = rate of background mortality for increased mortality rate from the Recovered compartment, multiplier = the multiplier to see the initial distribution of the population into the natural history compartments, omegaS0 = rate of progression between Latent-Slow and Latent-Zero, pEhigh = multiplier for high access-to-care relative to low access-to-care, pR = protection from reinfection for those in the Latency or Recovered compartments, pT = rate of transmission, rho = rate of relapse, sageA15 = age multiplier for mortality rates, sigma = rate of reactivation, theta = rate of fast progression following infection, zeta = rate of progression from subclinical to clinical disease compartments.*

##### 7.3 *Strengthened Current Interventions* no-new-vaccine baseline calibration

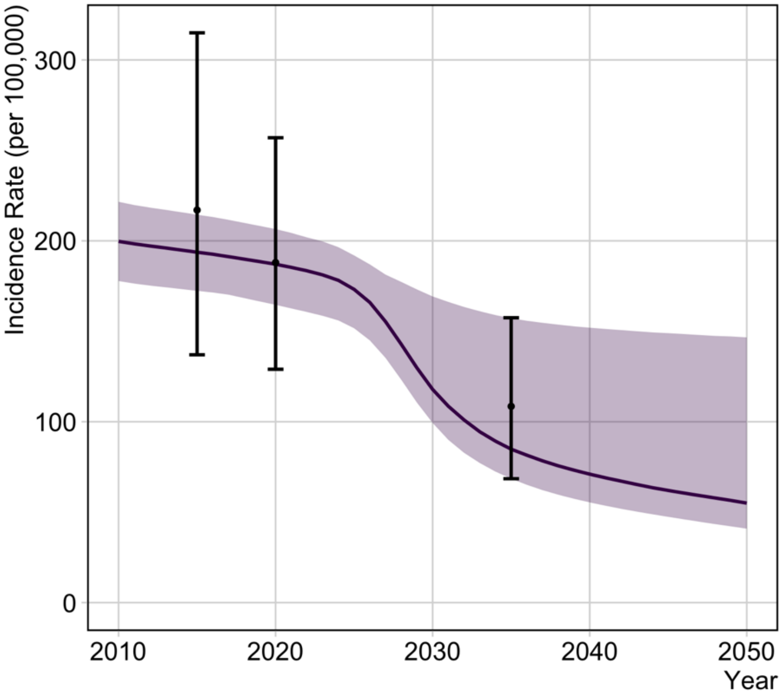


**Figure S7.8 Tuberculosis incidence rate for the calibrated *Strengthened Current Interventions* no-new-vaccine baseline**

*The black trend line indicates the median modelled output with 95% uncertainty in shaded grey. The black dot and vertical lines are the targets as described below.*

The targets indicated on the plot are (from left to right) the tuberculosis incidence rate per 100,000 population per year in the year:

- 2015 (not calibrated, but used to calculate the 2035 target):
- 2020 (the *Status Quo* baseline calibration target)
- 2035 (the *Strengthened Current Interventions* baseline calibration target—a 50% reduction compared to 2015 target)

### 8. Health impact results

#### 8.1 M72/AS01E scenarios

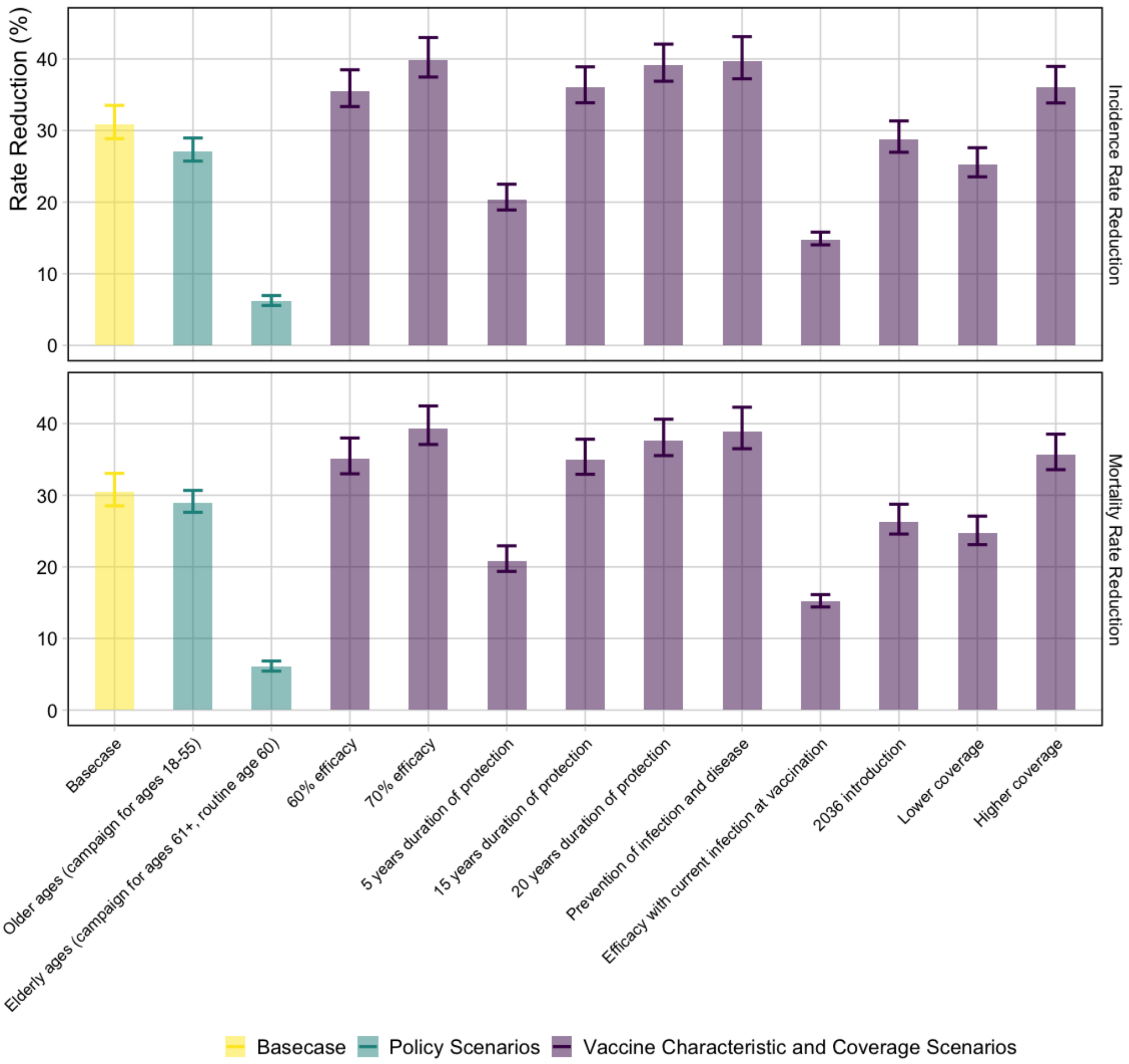


**Figure S8.1** **Incidence and mortality rate reductions in 2050 for the M72/AS01E scenarios (*Status Quo* baseline)**

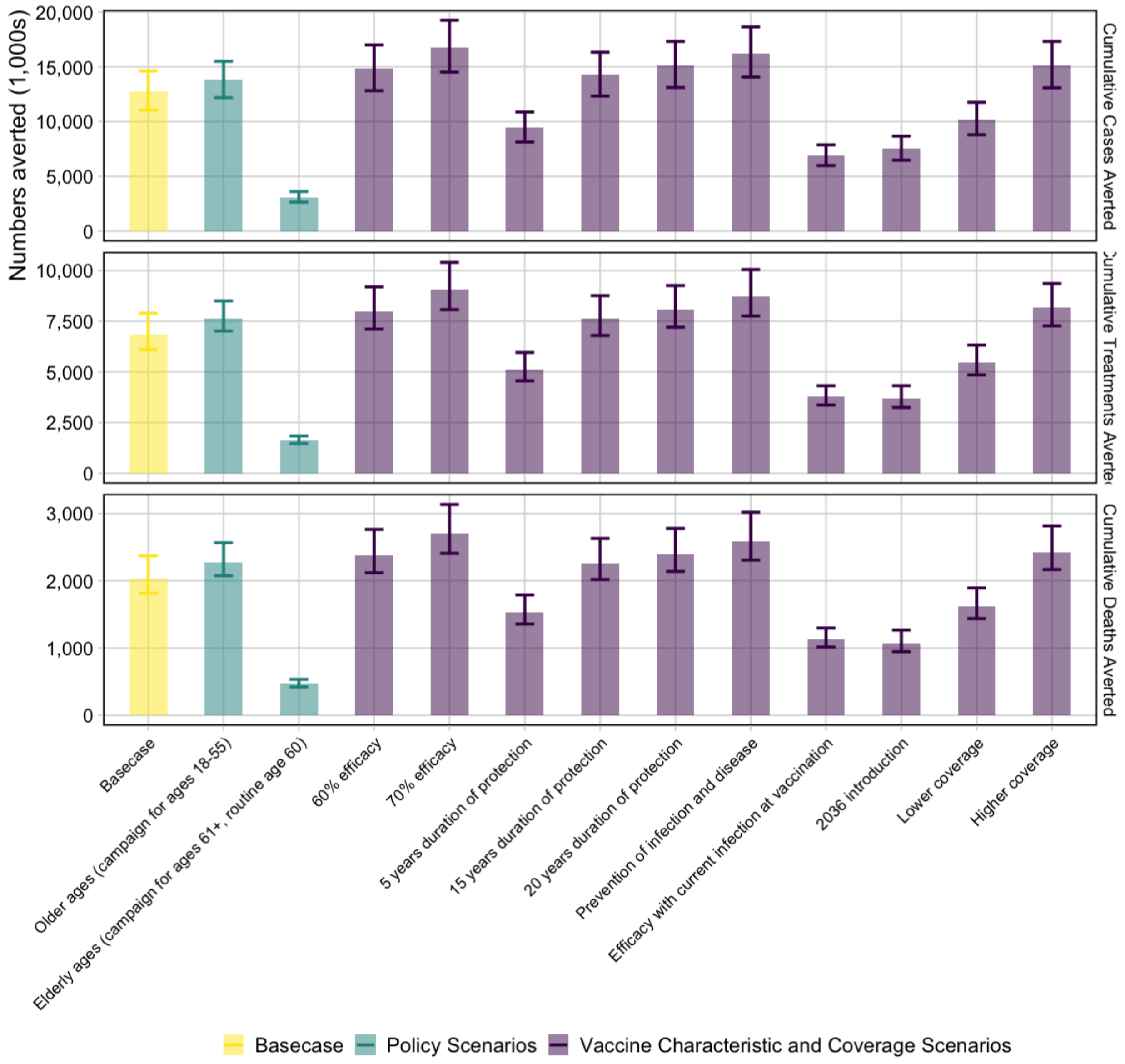


**Figure S8.2 Cumulative tuberculosis cases, treatments, and deaths averted between 2030 and 2050 for the M72/AS01E scenarios (*Status Quo* baseline)**

*The horizontal line is the median value of the Basecase for each vaccine, and the vertical line separates vaccine profile and delivery scenarios.*

#### Table S8.1 Health impact results for the M72/AS01$_E$ scenarios

| Scenario | IRR in 2050 (%) | MRR in 2050 (%) | Cumulative cases averted 2030–2050 | Cumulative treatments averted 2030–2050 | Cumulative deaths averted 2030–2050 |
|---|---|---|---|---|---|
| Basecase (*Status Quo* baseline) | 30.9% (28.9–33.5) | 30.4% (28.5–33.1) | 12.7m (11.0–14.6) | 6.9m (6.1–7.9) | 2.0m (1.8–2.4) |
| **Policy Scenarios (*Status Quo* baseline)** | | | | | |
| Older ages (campaign for ages 18-55) | 27.1% (25.7–29.0) | 28.9% (27.6–30.7) | 13.8m (12.2–15.5) | 7.6m (7.0–8.5) | 2.3m (2.1–2.6) |
| Elderly ages (campaign for ages 61+, routine age 60) | 6.2% (5.6–7.0) | 6.1% (5.4–6.9) | 3.1m (2.6–3.6) | 1.6m (1.5–1.8) | 0.5m (0.4–0.5) |
| **Vaccine Characteristic and Coverage Scenarios (*Status Quo* baseline)** | | | | | |
| 60% efficacy | 35.6% (33.3–38.5) | 35.1% (33.0–38.0) | 14.8m (12.8–17.0) | 8.0m (7.1–9.2) | 2.4m (2.1–2.8) |
| 70% efficacy | 39.8% (37.5–43.0) | 39.3% (37.1–42.5) | 16.8m (14.5–19.3) | 9.1m (8.1–10.4) | 2.7m (2.4–3.1) |
| 5 years protection | 20.4% (18.9–22.5) | 20.9% (19.4–22.9) | 9.4m (8.1–10.9) | 5.1m (4.6–6.0) | 1.5m (1.4–1.8) |
| 15 years protection | 36.0% (33.9–38.9) | 35.0% (32.9–37.8) | 14.3m (12.3–16.3) | 7.6m (6.8–8.8) | 2.3m (2.0–2.6) |
| 20 years protection | 39.1% (36.9–42.1) | 37.7% (35.5–40.6) | 15.2m (13.1–17.3) | 8.1m (7.2–9.3) | 2.4m (2.1–2.8) |
| Prevention of infection and disease | 39.7% (37.2–43.1) | 39.0% (36.5–42.3) | 16.2m (14.1–18.7) | 8.7m (7.8–10.0) | 2.6m (2.3–3.0) |
| Efficacious with current infection at vaccination | 14.8% (14.0–15.8) | 15.2% (14.4–16.1) | 6.9m (6.0–7.9) | 3.8m (3.4–4.3) | 1.1m (1.0–1.3) |
| 2036 introduction | 28.8% (27.0–31.3) | 26.3% (24.6–28.7) | 7.5m (6.5–8.7) | 3.7m (3.2–4.3) | 1.1m (0.9–1.3) |
| Lower coverage | 25.3% (23.5–27.6) | 24.8% (23.1–27.1) | 10.2m (8.8–11.8) | 5.5m (4.8–6.3) | 1.6m (1.4–1.9) |
| Higher coverage | 36.0% (33.9–39.0) | 35.7% (33.6–38.5) | 15.1m (13.1–17.3) | 8.2m (7.3–9.4) | 2.4m (2.2–2.8) |
| ***Strengthened Current Interventions* no-new-vaccine baseline** | | | | | |
| Basecase | 16.1% (4.8–30.5) | 17.1% (5.7–30.1) | 3.0m (1.1–11.3) | 1.7m (0.68–6.2) | 0.51 (0.19–1.9) |

*Abbreviations: IRR = incidence rate reduction, MRR = mortality rate reduction.*

#### 8.2 BCG-revaccination scenarios

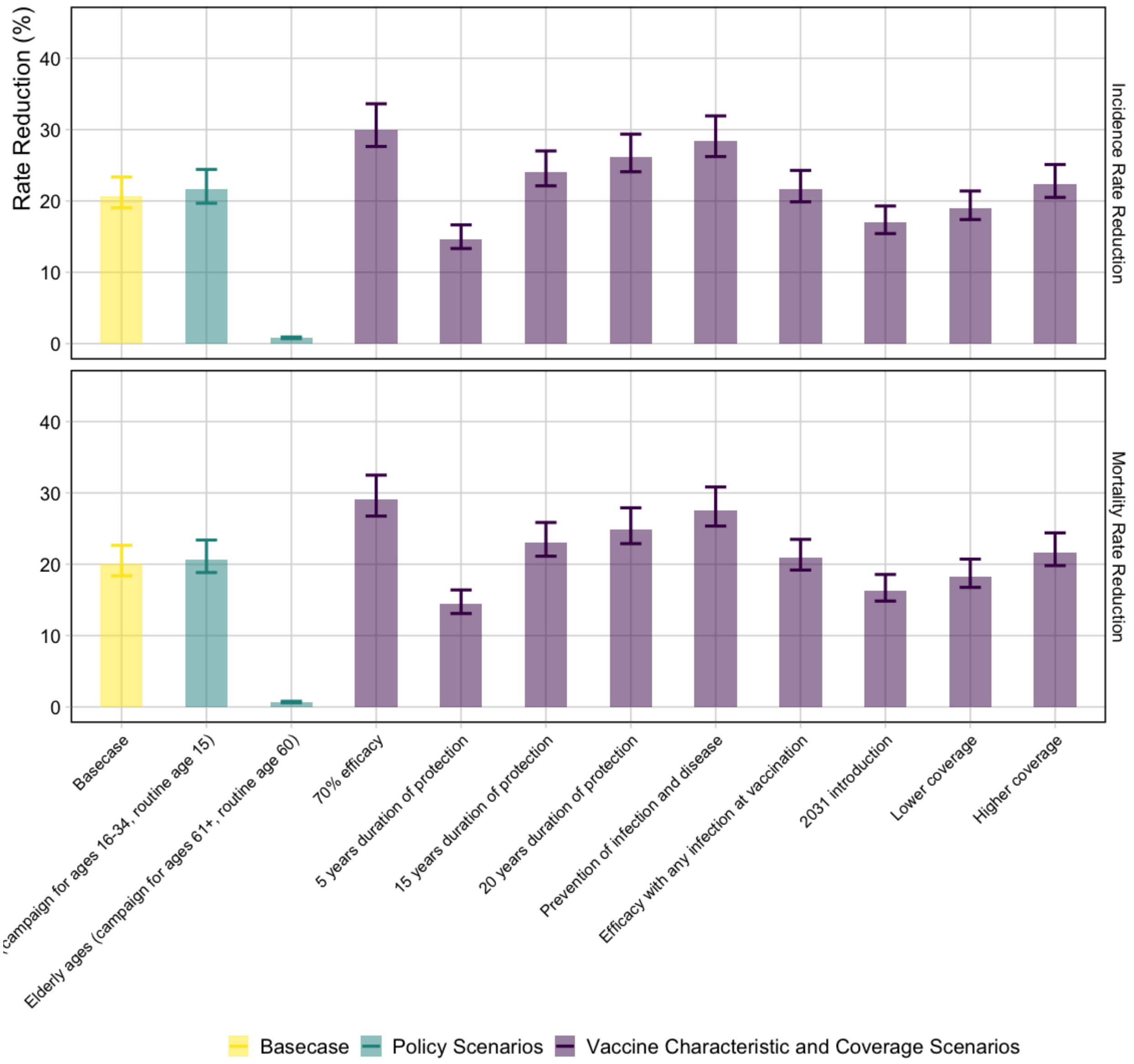


**Figure S8.3** **Incidence and mortality rate reductions in 2050 for the BCG-revaccination scenarios (*Status Quo* baseline)**

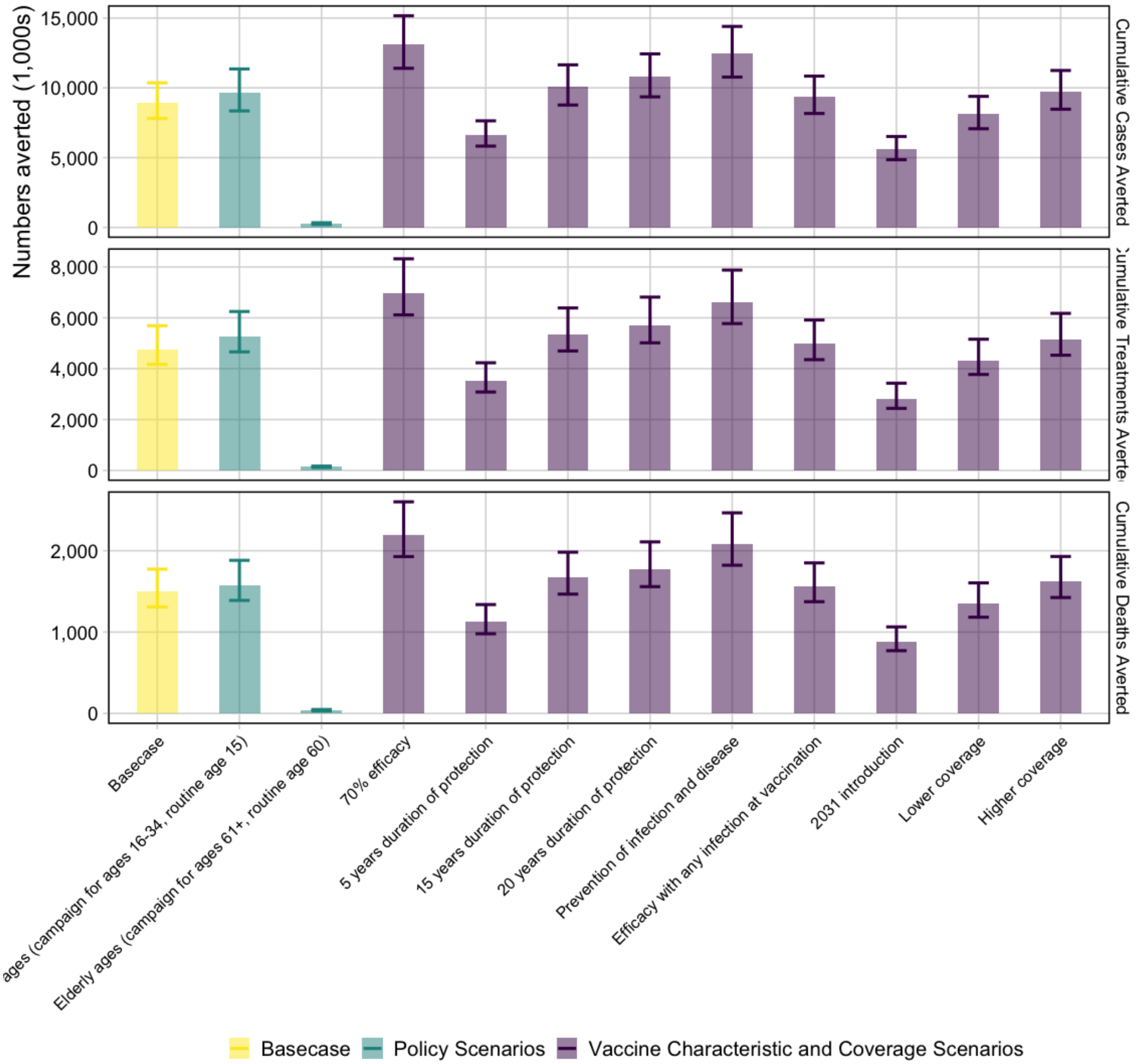


**Figure S8.4 Cumulative tuberculosis cases, treatments, and deaths averted between 2025 and 2050 for the BCG-revaccination scenarios (*Status Quo* baseline)**

*The horizontal line is the median value of the Basecase for each vaccine, and the vertical line separates vaccine profile and delivery scenarios.*

#### Table S8.2 Health impact results for the BCG-revaccination scenarios

| Scenario | IRR in 2050 (%) | MRR in 2050 (%) | Cumulative cases averted 2025–2050 | Cumulative treatments averted 2025–2050 | Cumulative deaths averted 2025–2050 |
|---|---|---|---|---|---|
| Basecase (*Status Quo* baseline) | 20.7% (19.0–23.4) | 20.0% (18.4–22.6) | 9.0m (7.8–10.4) | 4.8m (4.2–5.7) | 1.5m (1.3–1.8) |
| **Policy Scenarios (*Status Quo* baseline)** | | | | | |
| Older ages (campaign for ages 16-34, routine age 15) | 21.6% (19.7–24.4) | 20.7% (18.8–23.4) | 9.7m (8.3–11.4) | 5.3m (4.7–6.2) | 1.6m (1.4–1.9) |
| Elderly ages (campaign for ages 61+, routine age 60) | 0.8% (0.7–0.9) | 0.7% (0.6–0.8) | 0.3m (0.2–0.3) | 0.1m (0.1–0.2) | 0.04m (0.03–0.05) |
| **Vaccine Characteristic and Coverage Scenarios (*Status Quo* baseline)** | | | | | |
| 70% Efficacy | 30.0% (27.6–33.6) | 29.0% (26.7–32.5) | 13.2m (11.4–15.2) | 7.0m (6.1–8.3) | 2.2m (1.9–2.6) |
| 5 years protection | 14.6% (13.3–16.6) | 14.4% (13.1–16.4) | 6.6m (5.8–7.6) | 3.5m (3.1–4.2) | 1.1m (1.0–1.3) |
| 15 years protection | 24.0% (22.1–27.0) | 23.0% (21.1–25.9) | 10.1m (8.8–11.6) | 5.3m (4.7–6.4) | 1.7m (1.5–2.0) |
| 20 years protection | 26.1% (24.1–29.4) | 24.9% (22.9–27.9) | 10.8m (9.4–12.4) | 5.7m (5.0–6.8) | 1.8m (1.6–2.1) |
| Prevention of infection and disease | 28.4% (26.2–31.9) | 27.5% (25.3–30.8) | 12.4m (10.8–14.4) | 6.6m (5.8–7.9) | 2.1m (1.8–2.5) |
| Efficacious with any infection status at vaccination | 21.6% (19.9–24.3) | 20.9% (19.2–23.5) | 9.4m (8.2–10.8) | 5.0m (4.4–5.9) | 1.6m (1.4–1.9) |
| 2031 introduction | 17.0% (15.4–19.3) | 16.3% (14.8–18.6) | 5.6m (4.9–6.5) | 2.8m (2.4–3.4) | 0.9m (0.8–1.1) |
| Lower coverage | 19.0% (17.4–21.4) | 18.3% (16.8–20.7) | 8.1m (7.1–9.4) | 4.3m (3.8–5.2) | 1.4m (1.2–1.6) |
| Higher coverage | 22.3% (20.5–25.1) | 21.6% (19.8–24.4) | 9.7m (8.5–11.2) | 5.2m (4.5–6.2) | 1.6m (1.4–1.9) |
| ***Strengthened Current Interventions* no-new-vaccine baseline** | | | | | |
| Basecase | 8.9% (1.0–20.8) | 8.9% (1.0–20.1) | 1.9m (0.42–8.0) | 1.1m (0.25–4.4) | 0.34m (0.08–1.4) |

*Abbreviations: IRR = incidence rate reduction, MRR = mortality rate reduction.*

#### 9. Economic results

##### 9.1 M72/AS01E scenarios

**Table S9.1 Cost-effectiveness analysis for M72/AS01E *Policy Scenarios***

| Scenario | Total costs (USD, 1000s) | Total DALYs (1000s) | Total DALYs averted (1000s) | Incremental cost (USD, 1000s) | Incremental DALYs averted (1000s) | Cost (USD) per DALY averted |
|---|---|---|---|---|---|---|
| No-new-vaccine | 14,262,475 | 3,991,720 | – | 14,262,475 | – | – |
| Elderly ages (campaign for ages 61+, routine age 60) | 17,523,764 | 3,986,463 | 5,256.71 | – | – | *Weakly dominated* |
| Basecase (campaign for ages 16–34, routine age 15) | 19,596,068 | 3,954,863 | 36,856.95 | 5,333,593 | 36,856.95 | $144.71[+] |
| Older ages (campaign for ages 18–55) | 21,456,380 | 3,953,202 | 38,517.91 | 1,860,312 | 1,660.96 | $1,120.02[#] |

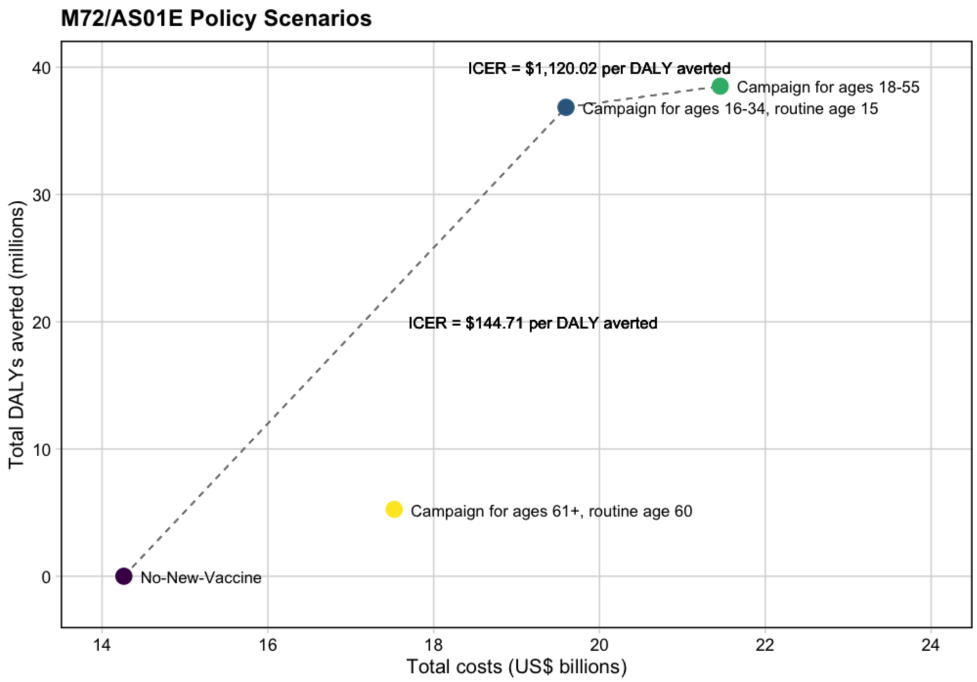


**Figure S9.1 Efficiency frontiers for M72/AS01E *Policy Scenarios***

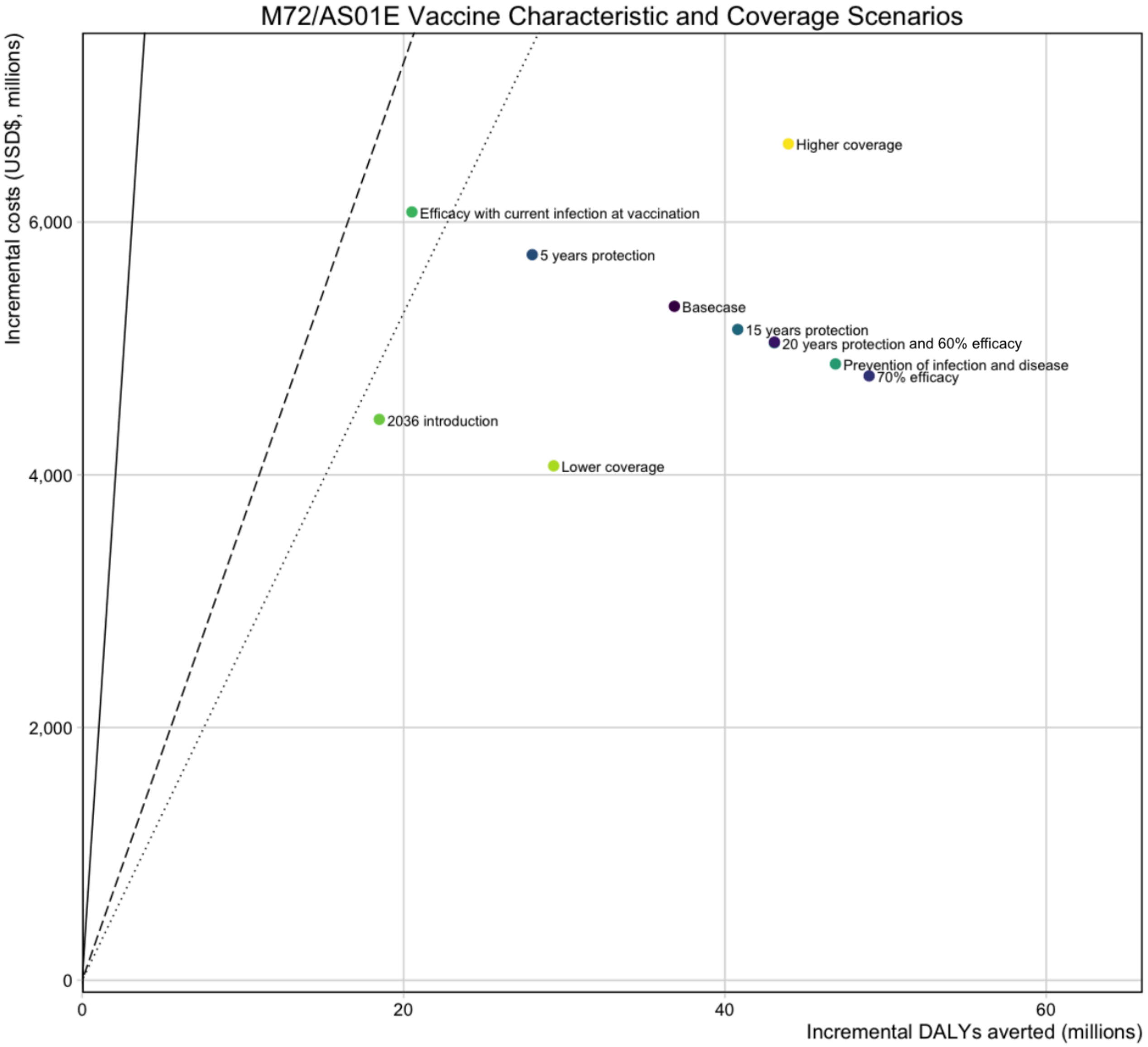


**Figure S9.2 Comparison of ICERs for M72/AS01$_E$ *Vaccine Characteristic and Coverage* Scenarios**

*Abbreviations: DALYs = disability-adjusted life years, USD$ = United States Dollars.*

*Points are the mean incremental costs and mean incremental DALYs averted for each scenario compared to the costs and DALYs from the no-new-vaccine baseline. The solid line represents 1x GDP, the dashed line represents the country-level upper bound, and the dotted line represents the country-level lower bound. The 20 years protection and 60% efficacy scenarios for the M72/AS01$_E$ vaccine overlap and appear as one single point on the figure.*

*The Basecase M72/AS01E scenario assumes a 50% efficacy POD vaccine efficacious with any infection status at the time of vaccination, with 10 years duration of protection reaching 80% coverage for 15-year-olds and 70% coverage for those aged 16–34. Each M72/AS01E scenario is delivered routinely to those aged 15 and as a campaign for those aged 16–34. The scenarios on the figure are labelled with the difference in product characteristics for that scenario compared to the Basecase.*

**Table S9.2** **Incremental DALYs averted, incremental costs averted, and ICERs from the health-system and societal perspectives for the M72/AS01E *Vaccine Characteristic and Coverage Scenarios* compared to the no-new-vaccine baseline**

| Scenario | Incremental DALYs averted between 2025–2050 (millions) | Health-system perspective | | Societal perspective | |
|---|---|---|---|---|---|
| | | Incremental costs between 2025–2050 ($, millions) | ICERs ($/DALY averted) | Incremental costs between 2025–2050 ($, millions) | ICERs ($/DALY averted) |
| Basecase | 36.9 (32.5, 42.9) | 5 334 (3 036, 8 573) | 145 (82, 236) | 5 948 (3 242, 9 272) | 161 (84, 259) |
| **Vaccine Characteristic and Coverage Scenarios** | | | | | |
| 60% efficacy | 43.1 (38.0, 50.1) | 5 050 (2 752, 8 311) | 117 (63, 196) | 5 552 (2 807, 8 976) | 129 (64, 213) |
| 70% efficacy | 49.0 (43.2, 56.9) | 4 783 (2 483, 8 036) | 98 (50, 167) | 5 177 (2 336, 8 617) | 106 (47, 180) |
| 5 years protection | 28.0 (24.6, 32.7) | 5 742 (3 445, 9 046) | 205 (122, 326) | 6 519 (3 874, 9 853) | 233 (132, 361) |
| 15 years protection | 40.8 (36.0, 47.4) | 5 151 (2 852, 8 391) | 126 (69, 209) | 5 692 (2 993, 9 077) | 140 (71, 228) |
| 20 years protection | 43.1 (38.0, 50.0) | 5 045 (2 745, 8 305) | 117 (63, 196) | 5 544 (2 798, 8 970) | 129 (64, 212) |
| Prevention of infection and disease | 46.9 (41.3, 54.5) | 4 878 (2 595, 8 119) | 104 (54, 176) | 5 310 (2 506, 8 744) | 113 (52, 191) |
| Efficacy with current infection at vaccination | 20.5 (18.2, 23.5) | 6 080 (3 794, 9 429) | 296 (184, 457) | 6 992 (4 402, 10 256) | 341 (211, 515) |
| 2036 introduction | 18.5 (16.2, 21.6) | 4 439 (2 710, 6 930) | 240 (145, 378) | 5 070 (3 102, 7 553) | 274 (162, 419) |
| Lower coverage | 29.3 (25.8, 34.2) | 4 071 (2 302, 6 555) | 139 (77, 228) | 4 525 (2 442, 7 087) | 154 (79, 248) |
| Higher coverage | 43.9 (38.8, 51.0) | 6 620 (3 790, 10 626) | 151 (85, 245) | 7 403 (4 145, 11 508) | 168 (88, 268) |

*Abbreviations: DALYs = disability-adjusted life years, ICERs = incremental cost-effectiveness ratio, US$ = United States Dollar. Values in cells are the mean and 95% uncertainty ranges.*

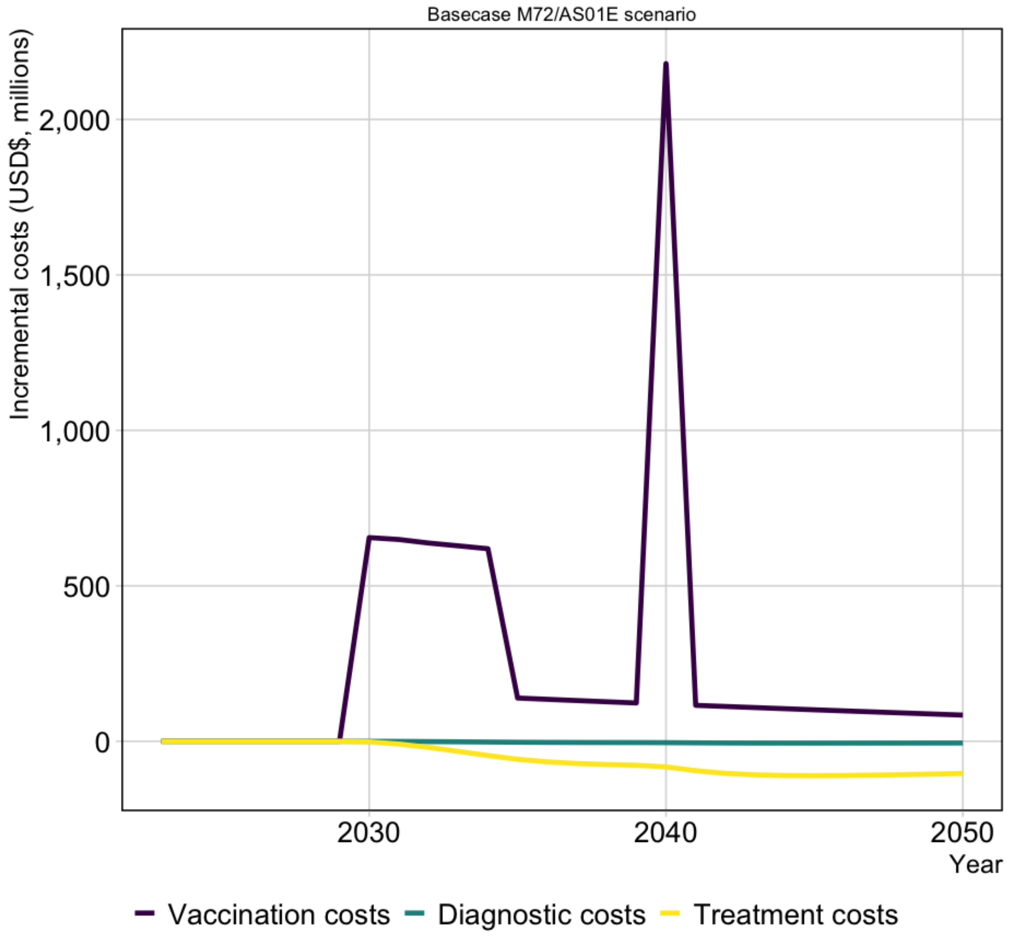


**Figure S9.3 Basecase M72/AS01$_E$ scenario incremental discounted costs (USD$, millions) by year**

*Abbreviations: DS-TB = drug-susceptible tuberculosis, RR-TB = rifampicin-resistant tuberculosis, USD$ = United States dollars.*

**Table S9.3 Total vaccination costs, and incremental diagnostic, treatment, and net costs between 2025–2050 for the M72/AS01E scenarios from the health-system perspective**

| Scenario | Vaccination costs (US$, millions) | DS-TB diagnostic costs (US$, millions) | RR-TB diagnostic costs (US$, millions) | DS-TB treatment costs (US$, millions) | RR-TB treatment costs (US$, millions) | Incremental cost (US$, millions) |
|---|---|---|---|---|---|---|
| Basecase | 7 021<br>(4 758, 10 301) | -84<br>(-103, -67) | -3<br>(-10, -0.01) | -1 183<br>(-1 467, -941) | -418<br>(-500, -352) | 5 334<br>(3 036, 8 573) |
| **Policy Scenarios** | | | | | | |
| Older ages (campaign for ages 18–55) | 9 097<br>(6 158, 13 365) | -94<br>(-114, -77) | -2.8<br>(-12, -0.01) | -1 334<br>(-1 610, -1 072) | -471<br>(-551, -402) | 7 194<br>(4 220, 11 502) |
| Elderly ages (campaign for ages 61+, routine age 60) | 3 663<br>(2 482, 5 362) | -20<br>(-24, -16) | -0.6<br>(-3, -0.003) | -282<br>(-348, -227) | -99<br>(-119, -84) | 3 261<br>(2 068, 4 962) |
| **Vaccine Characteristic and Coverage Scenarios** | | | | | | |
| 60% efficacy | 7 022<br>(4 758, 10 303) | -98<br>(-121, -78) | -3<br>(-12, -0.02) | -1 382<br>(-1 715, -1 099) | -488<br>(-584, -411) | 5 050<br>(2 752, 8 311) |
| 70% efficacy | 7 023<br>(4 759, 10 304) | -111<br>(-137, -89) | -3<br>(-14, -0.02) | -1 571<br>(-1 945, -1 249) | -555<br>(-664, -467) | 4 783<br>(2 483, 8 036) |
| 5 years protection | 7 020<br>(4 757, 10 300) | -63<br>(-79, -51) | -2<br>(-8, -0.01) | -896<br>(-1 115, -712) | -316<br>(-379, -266) | 5 742<br>(3 445, 9 046) |
| 15 years protection | 7 021<br>(4 758, 10 302) | -93<br>(-115, -74) | -3<br>(-12, -0.02) | -1 312<br>(-1 625, -1 043) | -463<br>(-554, -390) | 5 151<br>(2 852, 8 391) |
| 20 years protection | 7 022<br>(4 758, 10 302) | -98<br>(-121, -79) | -3<br>(-12, -0.02) | -1 386<br>(-1 717, -1 102) | -489<br>(-585, -413) | 5 045<br>(2 745, 8 305) |
| Prevention of infection and disease | 7 022<br>(4 759, 10 303) | -107<br>(-131, -85) | -3<br>(-13, -0.02) | -1 504<br>(-1 867, -1 196) | -531<br>(-635, -448) | 4 878<br>(2 595, 8 119) |
| Efficacy with current infection at vaccination | 7 019<br>(4 756, 10 298) | -47<br>(-57, -38) | -1<br>(-6, -0.01) | -658<br>(-806, -528) | -233<br>(-276, -195) | 6 080<br>(3 794, 9 429) |
| 2036 introduction | 5 284<br>(3 583, 7 749) | -42<br>(-52, -34) | -1<br>(-5, -0.01) | -593<br>(-739, -470) | -209<br>(-253, -174) | 4 439<br>(2 710, 6 930) |
| Lower coverage | 5 413<br>(3 670, 7 929) | -67<br>(-82, -53) | -2<br>(-8, -0.01) | -941<br>(-1 171, -748) | -332<br>(-399, -279) | 4 071<br>(2 302, 6 555) |
| Higher coverage | 8 633<br>(5 844, 12 679) | -100<br>(-123, -80) | -3<br>(-12, -0.02) | -1 412<br>(-1 747, -1 123) | -499<br>(-596, -420) | 6 620<br>(3 790, 10 626) |

*Abbreviations: DS-TB = drug-susceptible tuberculosis, RR-TB = rifampicin resistant tuberculosis, US$ = United States Dollars. Values in cells are the mean and 95% uncertainty ranges.*

**Table S9.4 Total vaccination costs, and incremental diagnostic, treatment, and net costs between 2025–2050 for the M72/AS01E scenarios from the societal perspective**

| Scenario | Vaccination costs (US$, millions) | Diagnostic costs (DS + RR-TB) (US$, millions) | Treatment costs (DS + RR-TB) (US$, millions) | Non-medical costs (US$, millions) | Indirect costs (US$, millions) | Incremental cost (US$, millions) |
|---|---|---|---|---|---|---|
| Basecase | 8307 (5720, 11621) | -86 (-107, -69) | -1601 (-1957, -1303) | -209 (-349, -112) | -463 (-1559, -22) | 5 948 (3 242, 9 272) |
| **Policy Scenarios** | | | | | | |
| Older ages (campaign for ages 18–55) | 10767 (7414, 15070) | -97 (-119, -79) | -1805 (-2151, -1493) | -236 (-388, -125) | -521 (-1715, -24) | 8 108 (4 663, 12 431) |
| Elderly ages (campaign for ages 61+, routine age 60) | 4327 (2969, 6025) | -21 (-25, -17) | -381 (-462, -314) | -50 (-82, -26) | -110 (-359, -5) | 3 766 (2 403, 5 492) |
| **Vaccine Characteristic and Coverage Scenarios** | | | | | | |
| 60% efficacy | 8308 (5721, 11623) | -101 (-125, -81) | -1871 (-2285, -1521) | -244 (-408, -130) | -541 (-1820, -25) | 5 552 (2 807, 8 976) |
| 70% efficacy | 8309 (5721, 11624) | -115 (-142, -92) | -2125 (-2594, -1728) | -278 (-462, -148) | -614 (-2066, -29) | 5 177 (2 336, 8 617) |
| 5 years protection | 8306 (5719, 11619) | -65 (-81, -52) | -1213 (-1486, -986) | -158 (-265, -85) | -351 (-1185, -16) | 6 519 (3 874, 9 853) |
| 15 years protection | 8308 (5720, 11622) | -96 (-119, -77) | -1775 (-2169, -1444) | -232 (-387, -124) | -513 (-1726, -24) | 5 692 (2 993, 9 077) |
| 20 years protection | 8308 (5721, 11622) | -101 (-125, -81) | -1876 (-2292, -1526) | -245 (-408, -131) | -542 (-1822, -25) | 5 544 (2 798, 8 970) |
| Prevention of infection and disease | 8309 (5721, 11624) | -110 (-136, -88) | -2035 (-2480, -1657) | -266 (-445, -142) | -588 (-1984, -28) | 5 310 (2 506, 8 744) |
| Efficacy with current infection at vaccination | 8305 (5718, 11617) | -48 (-60, -39) | -891 (-1078, -728) | -116 (-194, -62) | -257 (-855, -12) | 6 992 (4 402, 10 256) |
| 2036 introduction | 6252 (4304, 8743) | -43 (-54, -34) | -802 (-990, -646) | -105 (-175, -56) | -232 (-783, -11) | 5 070 (3 102, 7 553) |
| Lower coverage | 6401 (4406, 8946) | -69 (-85, -55) | -1273 (-1561, -1034) | -166 (-278, -89) | -368 (-1243, -17) | 4 525 (2 442, 7 087) |
| Higher coverage | 10218 (7037, 14300) | -103 (-128, -83) | -1910 (-2330, -1554) | -250 (-416, -133) | -552 (-1857, -26) | 7 403 (4 145, 11 508) |

*Abbreviations: DS-TB = drug-susceptible tuberculosis, RR-TB = rifampicin resistant tuberculosis, US$ = United States Dollars. Values in cells are the mean and 95% uncertainty ranges.*

#### 9.2 BCG-revaccination scenarios

**Table S9.5 Cost-effectiveness analysis for BCG-revaccination *Policy Scenarios***

| Scenario | Total costs (USD, 1000s) | Total DALYs (1000s) | Total DALYs averted (1000s) | Incremental cost (USD, 1000s) | Incremental DALYs averted (1000s) | Cost (USD) per DALY averted |
|---|---|---|---|---|---|---|
| No-new-vaccine | 14,262,475 | 3,991,720 | – | 14,262,475 | – | – |
| Basecase (campaign for ages 11–18, routine age 10) | 14,918,037 | 3,962,629 | 29,091.22 | 655,526 | 29,091.22 | \$22.53[+] |
| Older ages (campaign for ages 16–34, routine age 15) | 15,819,567 | 3,961,671 | 30,049.06 | 901,530 | 957.84 | \$941.21[#] |
| Elderly ages (campaign for ages 61+, routine age 60) | 15,922,705 | 3,991,270 | 449.83 | – | – | *Strongly dominated* |

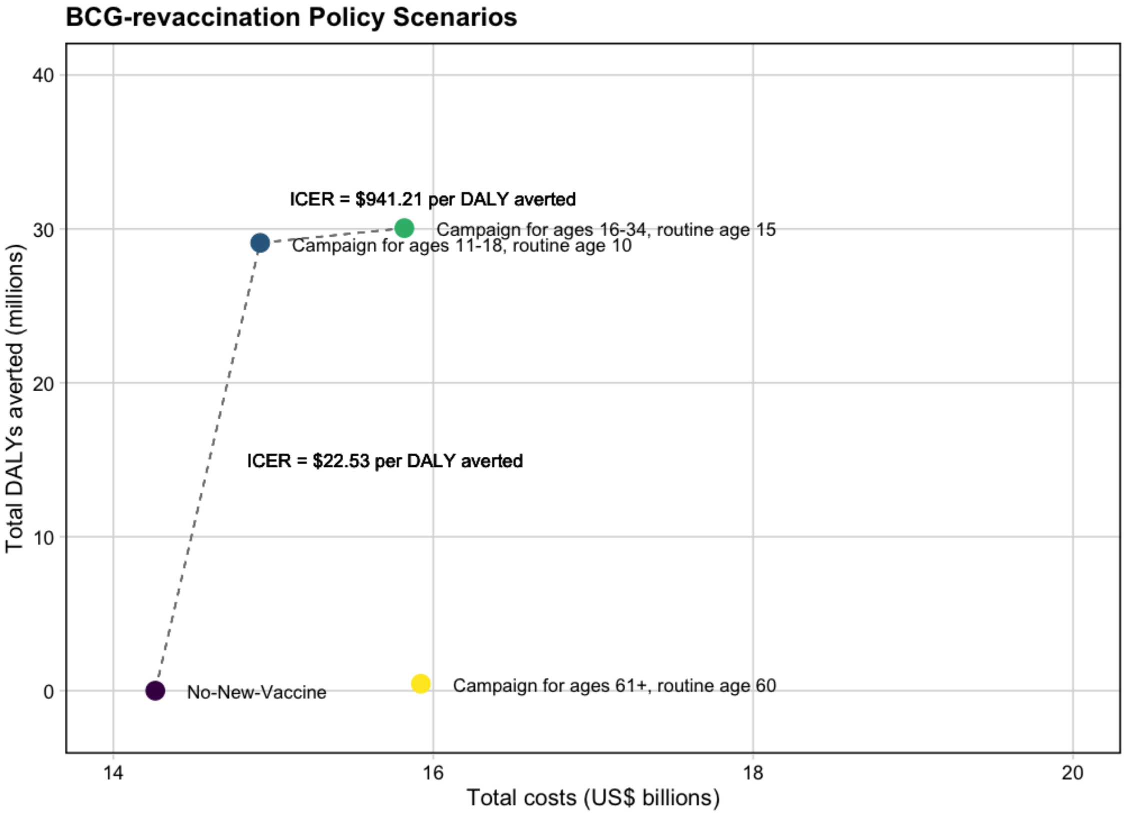


**Figure S9.4 Efficiency frontiers for BCG-revaccination *Policy Scenarios***

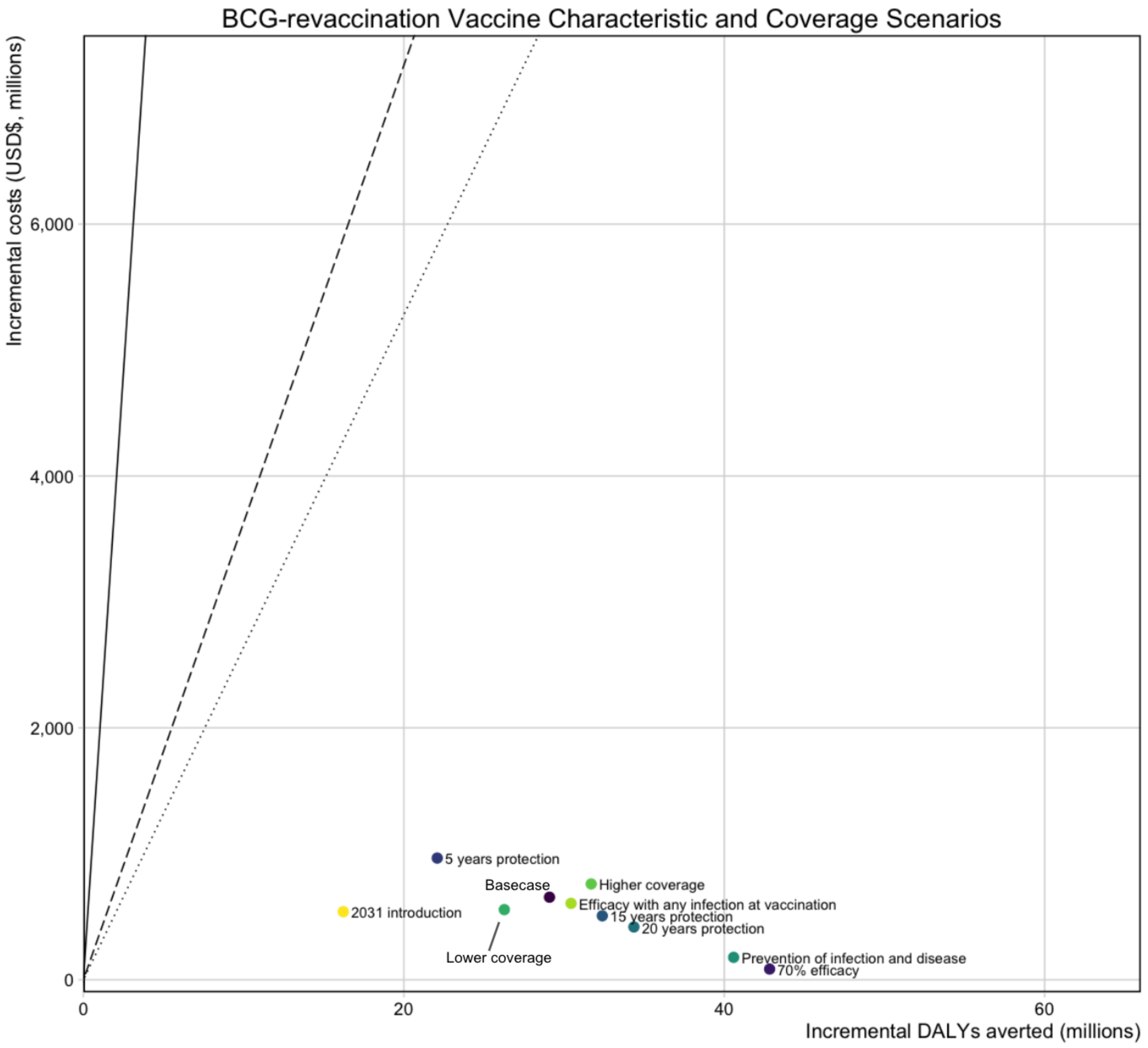


**Figure S9.5 Comparison of ICERs for BCG-revaccination *Vaccine Characteristic and Coverage* Scenarios**

*Abbreviations: DALYs = disability-adjusted life years, USD$ = United States Dollars.*

*Points are the mean incremental costs and mean incremental DALYs averted for each scenario compared to the costs and DALYs from the no-new-vaccine baseline. The solid line represents 1x GDP, the dashed line represents the country-level upper bound, and the dotted line represents the country-level lower bound.*

*The Basecase BCG-revaccination scenario assumes a 45% efficacy POI vaccine efficacious with no current infection at the time of vaccination, with 10 years duration of protection and reaching 80% coverage. Each BCG-revaccination scenario is delivered routinely to those aged 10 and as a campaign for those aged 11–18. The scenarios on the figure are labelled with the difference in product characteristics for that scenario compared to the Basecase.*

**Table S9.6** **Incremental DALYs averted, incremental costs averted, and ICERs from the health-system and societal perspectives for BCG-revaccination *Vaccine Characteristic and Coverage Scenarios* compared to the no-new-vaccine baseline**

| Scenario | Incremental DALYs averted between 2025–2050 (millions) | Health-system perspective | | Societal perspective | |
|---|---|---|---|---|---|
| | | Incremental costs between 2025–2050 ($, millions) | ICERs ($/DALY averted) | Incremental costs between 2025–2050 ($, millions) | ICERs ($/DALY averted) |
| Basecase | 29.1<br>(25.1, 34.6) | 656<br>(-442, 2 170) | 23<br>(cost-saving, 78) | 765<br>(-658, 2 405) | 26<br>(-22, 86) |
| **Vaccine Characteristic and Coverage Scenarios** | | | | | |
| 70% Efficacy | 42.8<br>(37, 51) | 85<br>(-1 058, 1 642) | 2<br>(cost-saving, 39) | -33<br>(-1 759, 1 735) | cost-saving<br>(cost-saving, 42) |
| 5 years protection | 22.1<br>(19, 26.3) | 966<br>(-108, 2 465) | 44<br>(cost-saving, 117) | 1 199<br>(-116, 2 778) | 54<br>(cost-saving, 131) |
| 15 years protection | 32.4<br>(27.9, 38.6) | 508<br>(-605, 2 031) | 16<br>(cost-saving, 65) | 559<br>(-954, 2 232) | 17<br>(cost-saving, 71) |
| 20 years protection | 34.4<br>(29.7, 40.9) | 419<br>(-704, 1 947) | 12<br>(cost-saving, 59) | 435<br>(-1113, 2 135) | 13<br>(cost-saving, 64) |
| Prevention of infection and disease | 40.6<br>(35, 48.4) | 178<br>(-958, 1 727) | 4<br>(cost-saving, 44) | 97<br>(-1556, 1 852) | 2<br>(cost-saving, 47) |
| Efficacy with any infection status at vaccination | 30.4<br>(26.4, 36.1) | 607<br>(-494, 2 143) | 20<br>(cost-saving, 73) | 696<br>(-756, 2 360) | 23<br>(cost-saving, 81) |
| 2031 introduction | 16.2<br>(13.9, 19.3) | 541<br>(-162, 1 509) | 33<br>(cost-saving, 98) | 658<br>(-231, 1 700) | 41<br>(cost-saving, 110) |
| Lower coverage | 26.3<br>(22.6, 31.3) | 557<br>(-414, 1 889) | 21<br>(cost-saving, 75) | 640<br>(-618, 2 085) | 24<br>(cost-saving, 83) |
| Higher coverage | 31.7<br>(27.3, 37.7) | 761<br>(-461, 2 457) | 24<br>(cost-saving, 82) | 899<br>(-668, 2 736) | 28<br>(cost-saving, 89) |

*Abbreviations: DALYs = disability-adjusted life years, ICERs = incremental cost-effectiveness ratio, US$ = United States Dollar. Values in cells are the mean and 95% uncertainty ranges.*

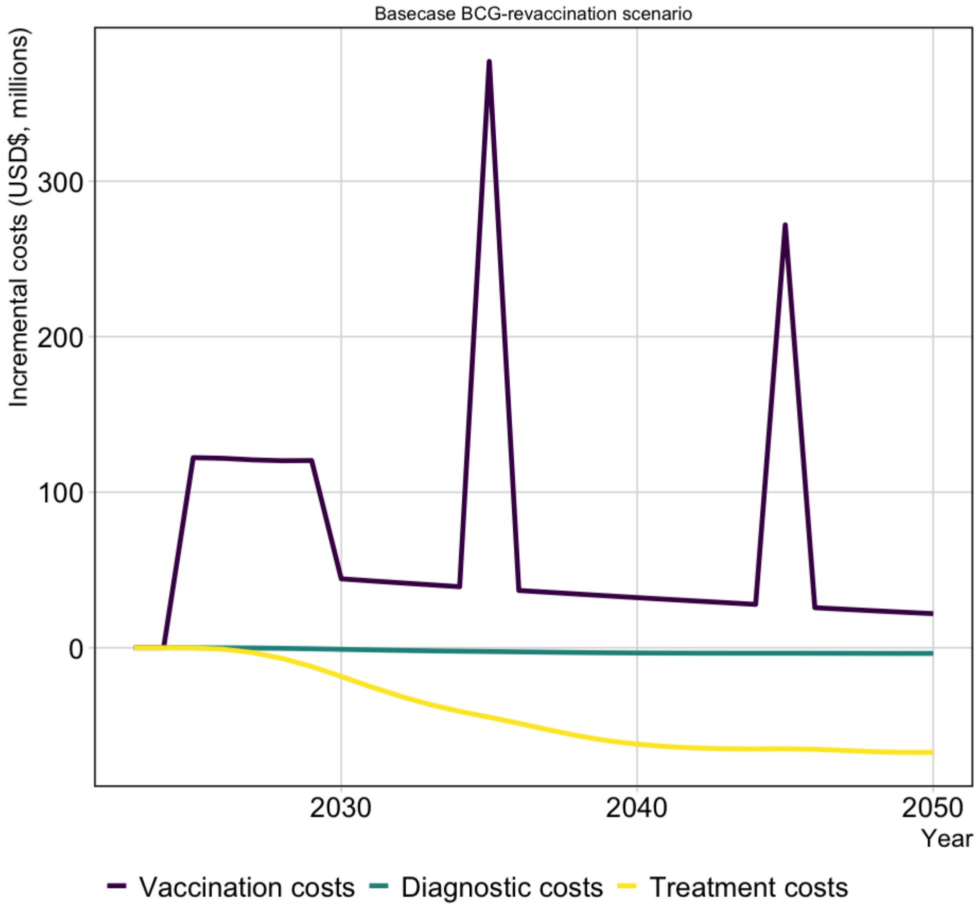


**Figure S9.6 Basecase BCG-revaccination scenario incremental discounted costs (US$, millions) by year**

*Abbreviations: USD$ = United States dollars.*

**Table S9.7 Total vaccination costs, and incremental diagnostic, treatment, and net costs between 2025–2050 for the BCG-revaccination scenarios from the health-system perspective**

| Scenario | Vaccination costs (US$, millions) | DS-TB diagnostic costs (US$, millions) | RR-TB diagnostic costs (US$, millions) | DS-TB treatment costs (US$, millions) | RR-TB treatment costs (US$, millions) | Incremental cost (US$, millions) |
|---|---|---|---|---|---|---|
| Basecase | 1 873 (804, 3 416) | -60 (-77, -48) | -2 (-8, -0.01) | -854 (-1 088, -674) | -301 (-368, -251) | 656 (-442, 2 170) |
| **Policy Scenarios** | | | | | | |
| Older ages (campaign for ages 16-34, routine age 15) | 2 930 (1 234, 5 368) | -68 (-86, -54) | -2 (-8, -0.01) | -962 (-1 219, -755) | -340 (-414, -282) | 1 557 (-142, 3 944) |
| Elderly ages (campaign for ages 61+, routine age 60) | 1 695 (730, 3 080) | -2 (-2, -1) | -0.1 (-0.2, 0) | -24 (-31, -19) | -9 (-11, -7) | 1 660 (695, 3 042) |
| **Vaccine Characteristic and Coverage Scenarios** | | | | | | |
| 70% efficacy | 1 877 (805, 3 423) | -89 (-113, -71) | -3 (-11, -0.02) | -1 257 (-1 600, -994) | -444 (-540, -367) | 85 (-1 058, 1 642) |
| 5 years protection | 1 876 (805, 3 420) | -45 (-57, -36) | -1 (-6, -0.01) | -638 (-811, -505) | -225 (-276, -187) | 966 (-108, 2 465) |
| 15 years protection | 1 872 (803, 3 413) | -68 (-86, -54) | -2 (-8, -0.01) | -956 (-1 219, -754) | -338 (-411, -280) | 508 (-605, 2 031) |
| 20 years protection | 1 871 (803, 3 412) | -72 (-91, -57) | -2 (-9, -0.01) | -1 018 (-1 296, -802) | -359 (-438, -298) | 419 (-704, 1 947) |
| Prevention of infection and disease | 1 874 (804, 3 417) | -84 (-107, -67) | -3 (-10, -0.01) | -1 189 (-1 517, -938) | -420 (-512, -348) | 178 (-958, 1 727) |
| Efficacy with any infection at vaccination | 1 881 (807, 3 428) | -63 (-80, -50) | -2 (-8, -0.01) | -893 (-1 141, -705) | -315 (-384, -262) | 607 (-494, 2 143) |
| 2031 introduction | 1 213 (520, 2 212) | -33 (-43, -26) | -1 (-4, -0.01) | -471 (-605, -368) | -166 (-205, -137) | 541 (-162, 1 509) |
| Lower coverage | 1 657 (714, 3 010) | -55 (-69, -43) | -2 (-7, -0.01) | -772 (-983, -609) | -272 (-333, -226) | 557 (-414, 1 889) |
| Higher coverage | 2 086 (892, 3 810) | -66 (-83, -52) | -2.0 (-8, -0.01) | -929 (-1 184, -735) | -328 (-400, -273) | 761 (-461, 2 457) |

*Abbreviations: DS-TB = drug-susceptible tuberculosis, RR-TB = rifampicin resistant tuberculosis, US$ = United States Dollars. Values in cells are the mean and 95% uncertainty ranges.*

**Table S9.8** **Total vaccination costs, and incremental diagnostic, treatment, and net costs between 2025–2050 for the BCG-revaccination scenarios from the societal perspective**

| Scenario | Vaccination costs (US$, millions) | Diagnostic costs (DS + RR-TB) (US$, millions) | Treatment costs (DS + RR-TB) (US$, millions) | Non-medical costs (US$, millions) | Indirect costs (US$, millions) | Incremental cost (US$, millions) |
|---|---|---|---|---|---|---|
| Basecase | 2467 (1238, 4002) | -62 (-78, -49) | -1155 (-1451, -929) | -151 (-251, -80) | -334 (-1134, -16) | 765 (-658, 2 405) |
| **Policy Scenarios** | | | | | | |
| Older ages (campaign for ages 16-34, routine age 15) | 3881 (1933, 6307) | -70 (-88, -55) | -1302 (-1624, -1049) | -170 (-286, -90) | -377 (-1282, -17) | 1 962 (-61, 4 453) |
| Elderly ages (campaign for ages 61+, routine age 60) | 2229 (1121, 3610) | -2 (-2, -1) | -33 (-42, -26) | -4 (-7, -2) | -10 (-31, 0) | 2 181 (1 074, 3 563) |
| **Vaccine Characteristic and Coverage Scenarios** | | | | | | |
| 70% efficacy | 2473 (1240, 4013) | -92 (-115, -72) | -1701 (-2137, -1374) | -222 (-368, -117) | -492 (-1672, -23) | -33 (-1 759, 1 735) |
| 5 years protection | 2471 (1239, 4009) | -47 (-59, -36) | -863 (-1080, -692) | -113 (-188, -60) | -250 (-846, -12) | 1 199 (-116, 2 778) |
| 15 years protection | 2466 (1237, 3999) | -70 (-88, -55) | -1294 (-1629, -1039) | -169 (-281, -89) | -374 (-1270, -18) | 559 (-954, 2 232) |
| 20 years protection | 2464 (1237, 3998) | -74 (-93, -58) | -1377 (-1730, -1106) | -180 (-299, -95) | -398 (-1353, -19) | 435 (-1113, 2 135) |
| Prevention of infection and disease | 2468 (1238, 4004) | -87 (-109, -68) | -1609 (-2022, -1296) | -210 (-348, -111) | -465 (-1582, -22) | 97 (-1556, 1 852) |
| Efficacy with any infection at vaccination | 2478 (1243, 4020) | -65 (-82, -51) | -1209 (-1513, -974) | -158 (-264, -83) | -350 (-1191, -16) | 696 (-756, 2 360) |
| 2031 introduction | 1598 (801, 2593) | -34 (-44, -27) | -638 (-802, -509) | -83 (-140, -44) | -185 (-627, -9) | 658 (-231, 1 700) |
| Lower coverage | 2179 (1096, 3526) | -56 (-71, -44) | -1044 (-1311, -839) | -136 (-227, -72) | -302 (-1024, -14) | 640 (-618, 2 085) |
| Higher coverage | 2753 (1377, 4467) | -68 (-85, -53) | -1257 (-1580, -1013) | -164 (-273, -87) | -364 (-1234, -17) | 899 (-668, 2 736) |

*Abbreviations: DS-TB = drug-susceptible tuberculosis, RR-TB = rifampicin resistant tuberculosis, US$ = United States Dollars. Values in cells are the mean and 95% uncertainty ranges.*

#### 9.3 Economic results with the *Strengthened Current interventions* no-new-vaccine baseline

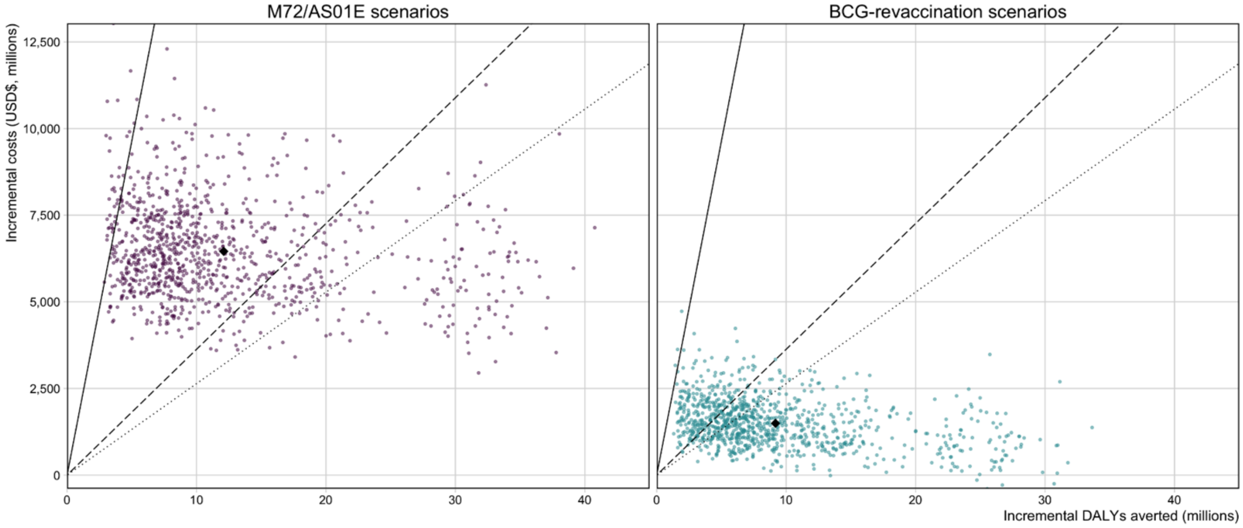


**Figure S9.7 Cost-effectiveness planes for the M72/AS01$_E$ and BCG-revaccination Basecase scenarios with the Strengthened Current interventions non-new-vaccine baseline**

*Abbreviations: USD$ = United States dollars The Black triangle is the mean estimate for the scenario, and each individual parameter set is represented as a single dot.*

*The solid line represents 1x GDP, the dashed line represents the country-level upper bound, and the dotted line represents the country-level lower bound.*

44. Gavi The Vaccine Alliance. GAVI Alliance Vaccine Introduction Grant and Operational Support for Campaigns Policy. Version No. 1.0. Geneva: Gavi, The Vaccine Alliance. 2 April 2013. www.gavi.org (accessed July 12, 2018).
